# Longitudinal assessment of ROPRO as an early indicator of overall survival in oncology clinical trials: a retrospective analysis

**DOI:** 10.1101/2022.10.11.22280399

**Authors:** H. Loureiro, T. M. Kolben, A. Kiermaier, D. Rüttinger, N. Ahmidi, T. Becker, A. Bauer-Mehren

**Author notes:** These authors contributed equally (T. Becker, A. Bauer-Mehren). **Address for Correspondence:** Dr. Anna Bauer-Mehren, Pharmaceutical Research and Early Development informatics (pREDi), Roche Diagnostics GmbH, PXID.2246, Nonnenwald 2, 82377 Penzberg / Germany, Mail. **Contribution info:** HL wrote the codes, performed the analysis and wrote the draft of the publication. TB contributed to the conceptualization of this study, supported the data preparation and contributed significantly to the manuscript. ABM, AK, DR, and TK contributed to the conceptualization of this study, supervised the study project with respect to clinical application and contributed to the manuscript. NA supervised ML aspects of the project, edited the manuscript significantly.

## Abstract

**Background:** The gold standard to evaluate treatment efficacy in oncology clinical trials is Overall Survival (OS). Its utility, however, is limited by the need for long trial duration and large sample sizes. Thus methods such as Progression-Free Survival (PFS) are applied to obtain early OS estimates across clinical trial phases, particularly to decide on further development of new molecular entities. Especially for cancer-immunotherapy, these established methods may be less suitable. Therefore, alternative approaches to obtain early OS estimates are required. In this work, we present a first evaluation of a new method, ΔRisk. ΔRisk uses the ROPRO, a state-of-the-art pan-cancer OS prognostic score, or DeepROPRO to predict OS benefit by measuring the patient’s improvement since baseline.

**Patients and methods:** We modeled the ΔRisk using Joint Models and tested whether a significant ΔRisk decrease correlated with OS improvement. We studied this hypothesis by comparing classical OS analysis against ΔRisk in a retrospective analysis of 12 real-world data emulated clinical trials, and 3 additional recent phase III immunotherapy clinical trials.

**Results:** Our new ΔRisk method correlated with the final OS readout in 14 out of 15 clinical trials. The ΔRisk, however, identified the treatment benefit up to seven months earlier than the OS log-rank test. Additionally, in two immunotherapy trials where PFS would have failed as an early OS estimate, the ΔRisk correctly predicted the treatment benefit.

**Conclusions:** We introduced a new method, ΔRisk, and demonstrated its correlation with OS. In retrospective analysis, ΔRisk is able to identify OS benefit earlier than standard methodology, and we show examples of lung cancer trials, where it maintains its predictive relevance whereas PFS does not correlate with OS. ΔRisk may prove useful for early decision support resulting in reduced need of resources. We also show the potential of ΔRisk as a candidate to define surrogate endpoints. To this purpose, more methodological work and further investigation of treatment-specific performance will be done in the future.

## Introduction

In oncology clinical trials, the gold-standard measure of efficacy is overall survival (OS). OS measures the time from randomization to the death of the patients (time-to-event) (Food and Drug Administration 2018). To get a reliable estimate of OS, a high number of patients and a long follow-up might be required (Mushti, Mulkey, and Sridhara 2018). These requirements constrain the estimation of OS across clinical trial phases: (1) in early clinical trial phases (phase I and II) where both the number of patients and follow-up time are limited, as well as (2) in interim analyses of late phases (phase III), where the follow-up time might still be a limiting factor. In order to obtain an early OS prediction, new endpoints allowing an early estimate of OS are warranted, particularly as existing surrogate endpoints may not always correlate with OS.

The objective response rate (ORR) and progression-free survival (PFS) are two of the most commonly used OS surrogate endpoints in solid tumors (Savina et al. 2018; Fiteni, Westeel, and Bonnetain 2017). Both of these imaging-based endpoints have shown a good correlation with OS in later clinical trial phases (phase III) of regular cytotoxic chemotherapies (Mauguen et al. 2013; Savina et al. 2018). However, it is reported that both PFS and ORR show a weaker association with OS in new treatment types, such as immunotherapy (Mushti, Mulkey, and Sridhara 2018; Ye et al. 2020). In cancer immunotherapy, the tumor flare reaction (TFR, also called pseudoprogression), a side effect of checkpoint inhibitor therapy in solid tumors, may be misinterpreted as cancer progression (leading to an inflation of the number of progressions) (Taleb B 2019). While TFR may lead to an underestimation of the treatment effect in cancer immunotherapy, the opposite has also been observed in some trials (Tan et al. 2017).

Apart from imaging-based endpoints, oncology prognostic scores, such as ROPRO (Becker et al. 2020) or DeepROPRO (Loureiro et al. 2021), that were recently introduced by our group, correlate with the mortality risk. Both models combine a set of 27 parameters describing the host (demographics, vitals, and blood-test parameters), the lifestyle (BMI and smoking status) as well as the tumor characteristics, all of which are associated with cancer survival. To date, the ROPRO and DeepROPRO prognostic scores have mainly been used for patient enrichment strategies in early oncology clinical trials based on baseline inclusion/exclusion criteria. In this analysis, we demonstrate their utility in assessing the patient’s mortality risk evolution over time. Relevant ROPRO/DeepROPRO parameters are routinely measured in clinical trials making this methodology more translational. We employ the time-wise risk values in a new method, ΔRisk, which measures the patient-level deviation of these scores from the start of treatment. We hypothesize that the ΔRisk represents the actual improvement/deterioration of the patient’s condition/fitness over time. Therefore, in a clinical trial setting, we expect that the treatment arm whose patients have the highest ΔRisk improvement (lowest deterioration) should be the arm with the highest actual OS benefit (Figure 1).

**Figure 1.**
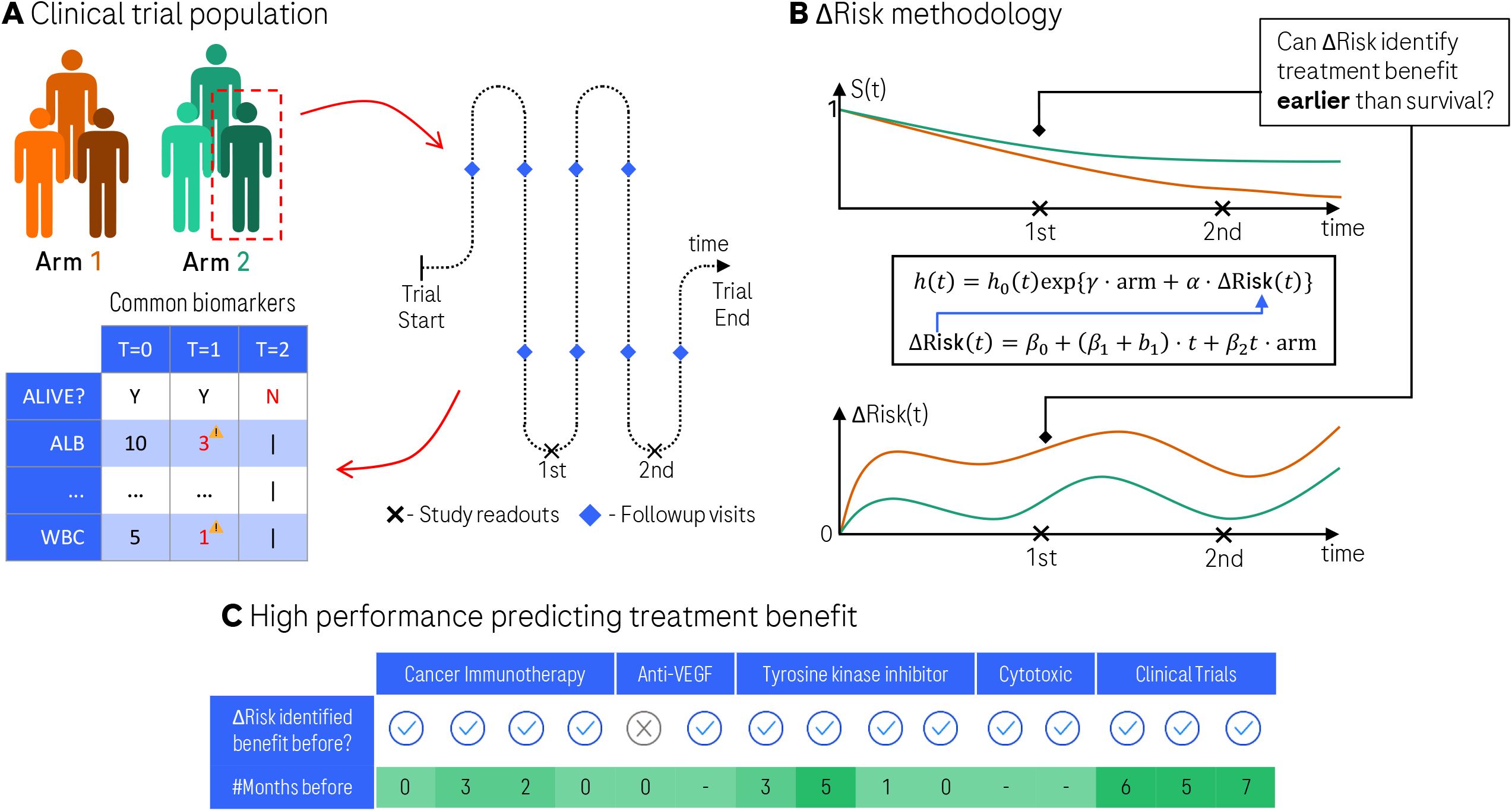
[Filename: “Visual abstract.pdf”]. The ΔRisk framework jointly models the risk variation and survival to identify early treatment benefit. (**A**) In a clinical trial, each patient has regular follow-up visits where several biomarker values are obtained. (**B**) At study readouts, previously obtained data is analyzed. At the readouts both the survival and risk values can be computed, yielding preliminary study results. The ΔRisk can be used at these points to obtain an indicator of treatment benefit. (**C**) The ΔRisk identified the treatment benefit in 14 out of 15 trials (including clinical trials emulated with real-world data).

In this first study of ΔRisk, we performed retrospective analyses to investigate its applicability as a R&D decision-support tool. We approached this by sequentially analyzing: (1) whether our ΔRisk scores (ΔROPRO and ΔDeepROPRO) correlate with OS, and if so, (2) contrast the performance of ΔROPRO/ΔDeepROPRO and PFS as early OS estimates. To study and validate these hypotheses, we gathered a broad set of recent oncology clinical trials. First, we emulated 12 recent clinical trials (covering a wide range of medication types) using data from a large real-world database to test the applicability of ΔRisk in early- and late-phase clinical trials. In addition, we validated our methods in 3 late-phase immunotherapy clinical trials conducted by Roche. In this work, we investigated the suitability of ΔRisk as a R&D decision-support tool to inform decision making in clinical trials. A formal validation of ΔRisk as a surrogate endpoint for OS will require access to more clinical data and thorough analysis following the Prentice methodology (Prentice 1989; Alonso et al. 2016; Burzykowski, Molenberghs, and Buyse 2005). We plan to investigate this in future work. We also discuss how the ΔRisk framework could be used as a R&D decision support tool in clinical trials to obtain an earlier OS estimate.

## Methods

### ΔRisk framework

The ΔRisk measures the evolution of mortality risk from the start of treatment. We derived the mortality risk values of the patients with ROPRO (Becker et al. 2020) and DeepROPRO (Deep learning-based model) (Loureiro et al. 2021). Both ROPRO and DeepROPRO are correlated with the mortality risk so that higher risk values are associated with shorter times-to-event (Becker et al. 2020; Loureiro et al. 2021). We call the ΔRisk values ΔROPRO and ΔDeepROPRO, when employing the ROPRO and DeepROPRO, respectively.

To appropriately use the ΔRisk values to compare and measure treatment effects, we developed the ΔRisk framework. This framework consists of three components: (1) a visualization tool, (2) a modeling approach, and (3) a statistical test.

First, the visualization tool uses the LOESS smoother (Cleveland and Devlin 1988) to represent the ΔRisk curves over time for both the treatment and control arms of a given clinical trial (refer to Figure 2 for three examples). We should note that uneven right censoring in the follow-up might induce artificial bias in the ΔRisk curves (more details in the Supplementary information). Therefore, direct interpretation of the ΔRisk curves should be done carefully to avoid misinterpretation of the real effect.

**Figure 2.**
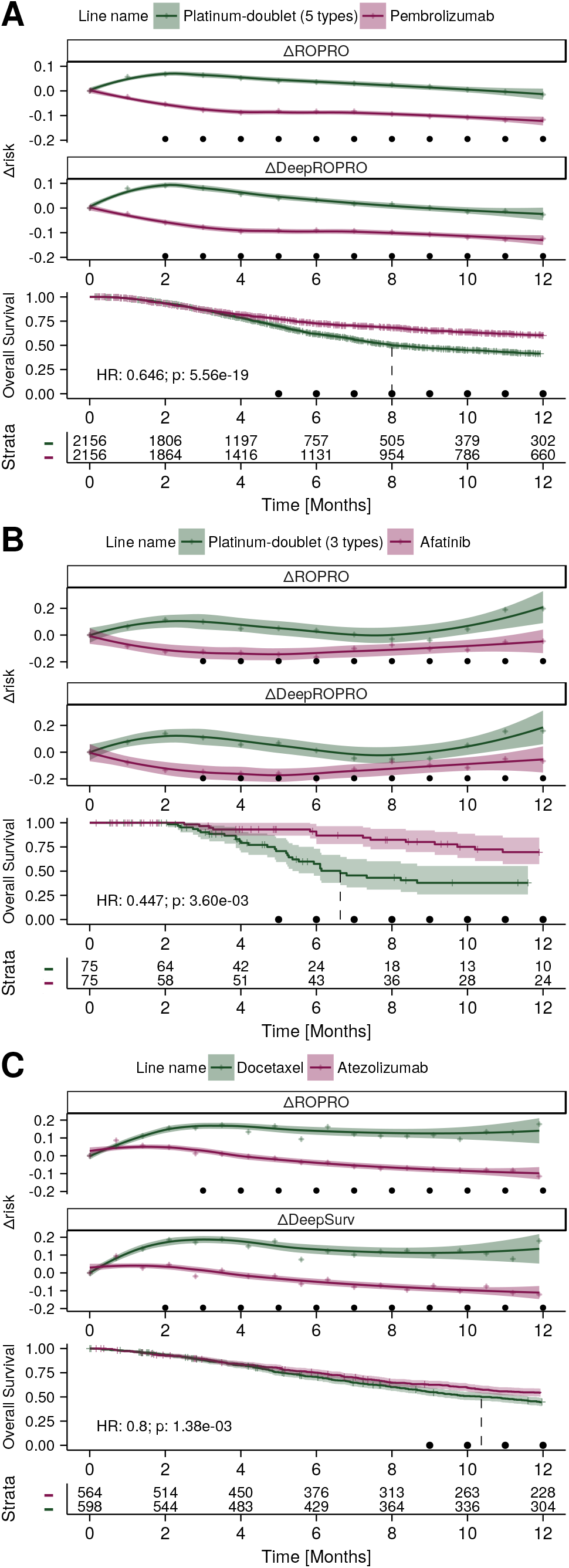
[Filename: “ΔRisk examples plot”]: Superimposed ΔRisk and survival curves of three distinct clinical trials. The clinical trials KEYNOTE-024 (higher caliper), LUX-Lung 3+6 (lower caliper), and OAK are displayed in the panels A, B, and C, respectively. In each panel, the first two plots are the ΔROPRO and ΔDeepROPRO for both arms of treatment. Followed by the OS Kaplan Meier curves, and the risk table. The Hazard Ratio (HR) is referent to the effect of the second medication (in purple, Pembrolizumab, Afatinib, and Atezolizumab, respectively). In the ΔROPRO and ΔDeepROPRO plots the filled line and confidence intervals were determined with LOESS. While the “+” symbols represent the mean of the ΔROPRO and ΔDeepROPRO, respectively. The round dots in the ΔRisk and OS curves represent the times at which the arms of treatment were significantly different according to the likelihood ratio and log-rank tests, respectively.

Second, we modeled the ΔRisk using the Joint Models for Longitudinal and Survival Data (in short JM) (Rizopoulos 2012) to overcome the negative bias induced by the uneven censoring. JM couples the ΔRisk (the longitudinal variable) with the survival information, measuring the impact that the ΔRisk trend has on survival, and hence eliminating the possible bias (Rizopoulos 2012).

Third, we created a statistical test based on the likelihood ratio to accurately measure the difference between the treatment arms. The statistical test measures the combined difference on the ΔRisk trend (model *β*_2_ parameter), and on early mortality (model γparameter) between the treatment arms. In other words, at a given time-point, we combined the information on ΔRisk trend and the OS information available until that time-point into a joint test. In the Supplementary information, we demonstrate that both parameters improve the model in their own right and that overall best performance is achieved when both parameters are combined.

### ΔRisk analysis

In this first analysis of the ΔRisk framework, we explored the following hypotheses: (Hypothesis 1) the treatment arm with the highest ΔRisk trend also showed the highest mortality rate (in essence, ΔRisk values are concordant with OS), (Hypothesis 2) the ΔRisk statistical test detects the treatment benefit earlier (or at the same time) than standard OS analysis, (Hypothesis 3) for immunotherapies the ΔRisk statistical test detects the treatment benefit before PFS, and, lastly, (Hypothesis 4) the early ΔRisk coefficients are predictive of OS at late cut-off points (Sargent et al. 2011).

To study these hypotheses, for each individual clinical trial we modeled the ΔRisk values with JM at every time-point starting from 2 to 24 months. At each time-point, we censored all future patient information, including ΔRisk values and event-times. For the first hypothesis, we compared the OS results of each trial with its JM coefficients. We confirmed this hypothesis if the treatment arm with the highest mortality rate also had the highest ΔRisk trend, or if there was no difference in ΔRisk and OS between the treatment arms. For the second hypothesis (only applicable when one treatment arm had OS benefit), we contrast the time-points at which the ΔRisk and log-rank (Peto and Peto 1972) tests identified a significant difference between the arms. We confirmed the second hypothesis if the ΔRisk statistical test identified a significant difference between the arms before or at the same time as the log-rank test. For the third hypothesis, we compared the ΔRisk results from the second hypothesis with PFS. We considered that the third hypothesis was confirmed if the ΔRisk identified the treatment benefit before PFS. Finally, for the fourth hypothesis, we followed the “simple” or trial-level surrogacy analysis from Sargent et al. (Sargent et al. 2011). Specifically, we performed a linear regression between the ΔRisk trend (β_2_) and mortality (γ) values from early time-points (we considered 3, 6, 9 and 12 months), and OS at later cut-offs (at 2, 3 and 4 years). From the linear regressions, we calculated the coefficients of determination (*R*^2^). We confirmed hypothesis four if there were high *R*^2^(we considered a *R*^2^ *≥0*.*75* as high correlation) between the early ΔRisk and the final OS. In this introductory work of the ΔRisk our focus was not to perform a complete surrogacy analysis as introduced by Prentice (Prentice 1989).

To study and validate these four hypotheses, we gathered 15 recent oncology clinical trials: 12 trials were emulated using a large real-world dataset, and 3 immunotherapy clinical trials conducted by Roche. Our analysis with the real-world dataset allowed us to test the ΔRisk framework in datasets similar to early- and late-phase clinical trials. These analyses would be impossible otherwise, due to lack of data. The 3 additional immunotherapy trials further validate the late-phase results.

### Clinical trials: emulation with real-world data (RWD)

To validate our ΔRisk framework on a broad range of medications, we emulated previous clinical trials with real-world data (RWD). First, we gathered a comprehensive list of clinical trials from ClinicalTrials.gov. Next, we emulated these clinical trials using the de-identified electronic health record (EHR)-derived Flatiron Health (FH) database. The FH database is a longitudinal database, comprising de-identified patient-level structured and unstructured data, curated via technology-enabled abstraction (Ma et al. 2020; Birnbaum et al. 2020). From FH, we extracted de-identified information collected between January 2011 and December 2020 from approximately 280 US cancer clinics (∼800 sites of care) about the first-line treatment of 34,061 patients diagnosed with advanced non-small-cell lung cancer (aNSCLC). Institutional review board approval of the study protocol was obtained prior to study conduct, and included a waiver of informed consent.

We focused our search on phase III studies in aNSCLC since: (1) aNSCLC is one of the most common types of cancer, and (2) phase III trials usually report OS results. We extracted a list of 184 clinical trials from the ClinicalTrials.gov website (accessed on Oct 5, 2021). From the initial list, we excluded 96 clinical trials based on the trial design (refer to the Supplementary Figure 1 for detailed criteria). Next, we excluded 75 additional clinical trials for which fewer than 200 patients were available for any treatment arm on the selected FH population. The final list of 12 potentially reproducible clinical trials is included in Supplementary Table 1 (following the analysis in Yang et al. (Yang et al. 2015) we incorporated LUX-Lung 3 and LUX-Lung 6 together, lowering the number of trials by one).

**Table 1:**
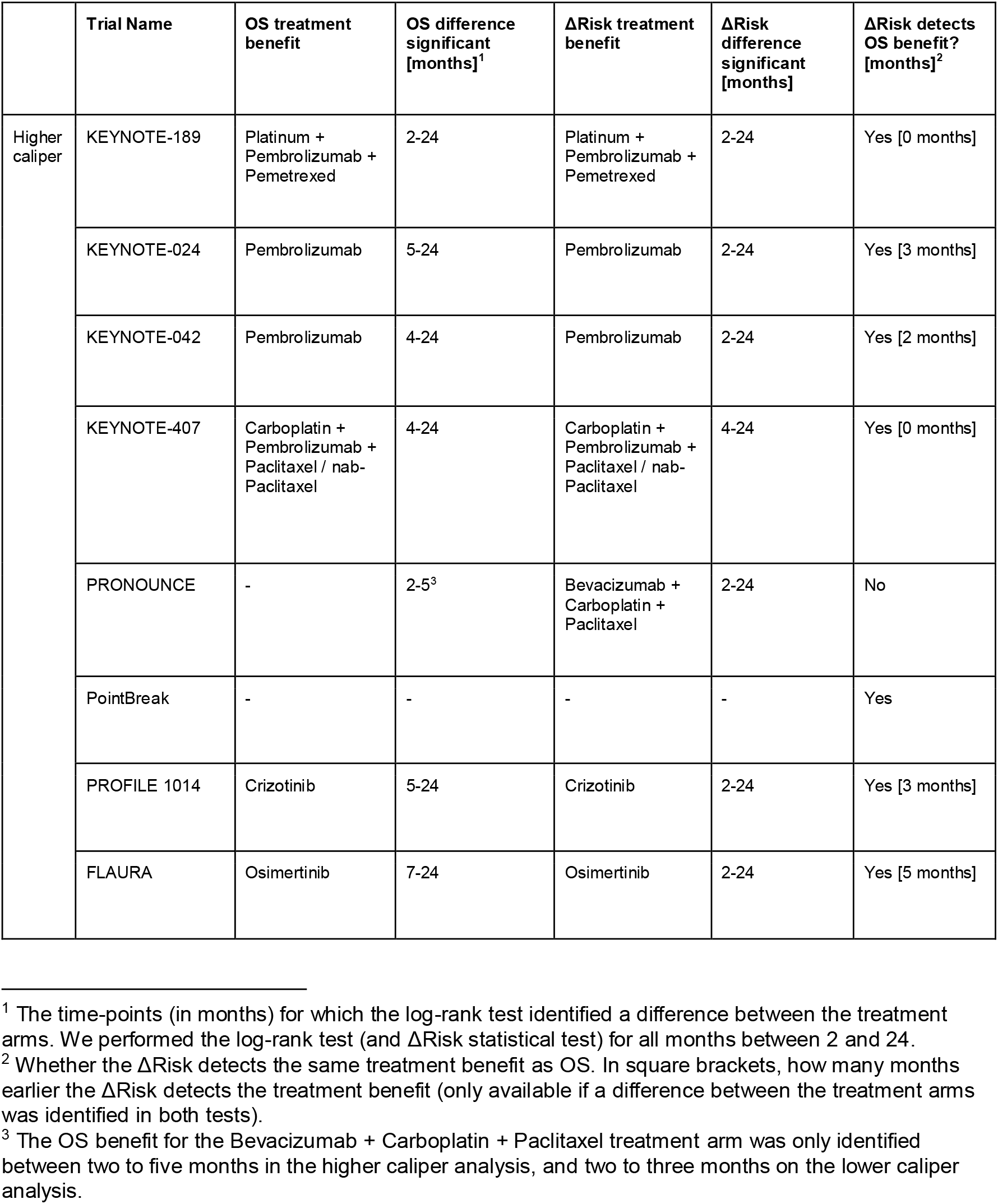
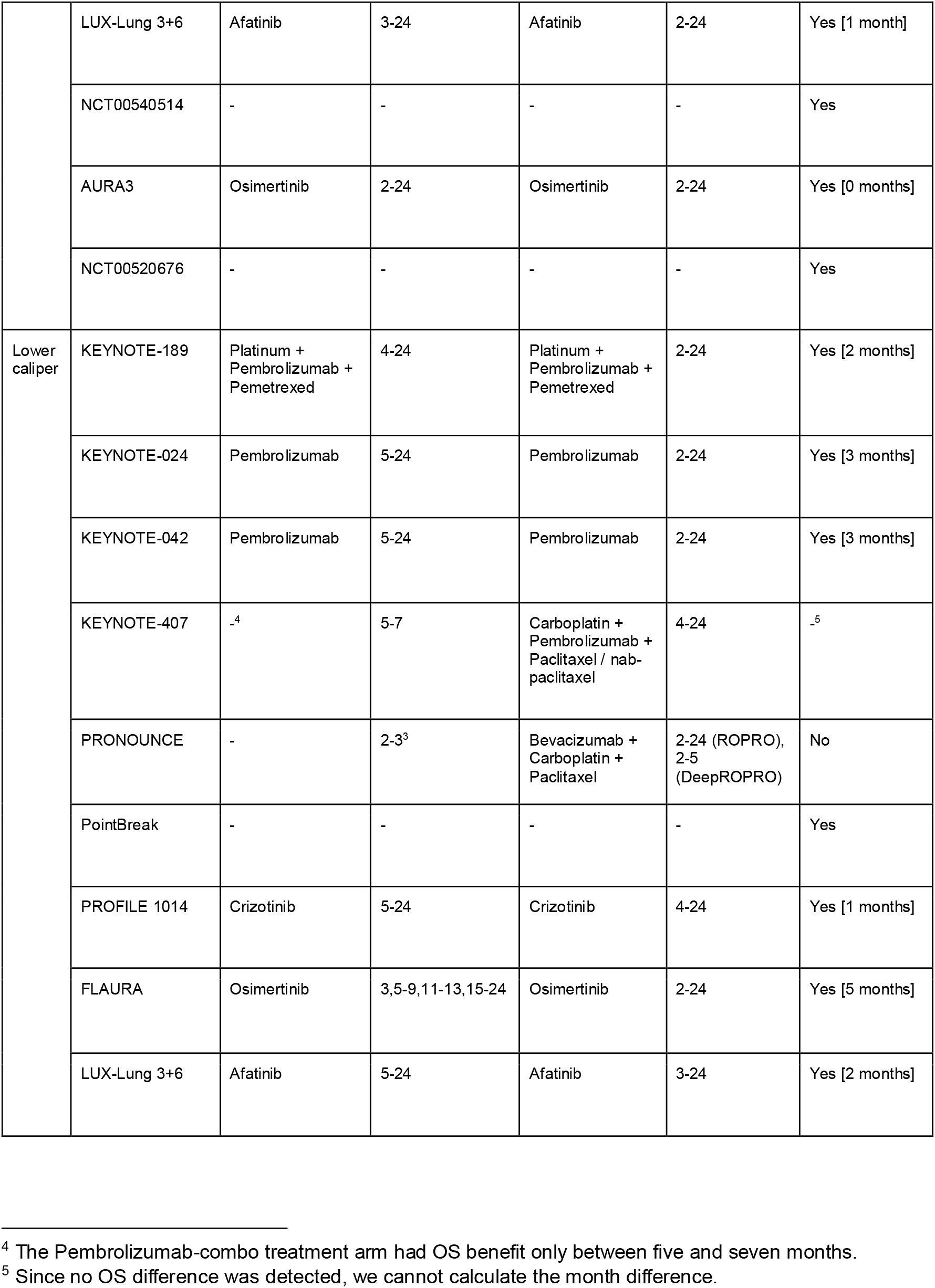
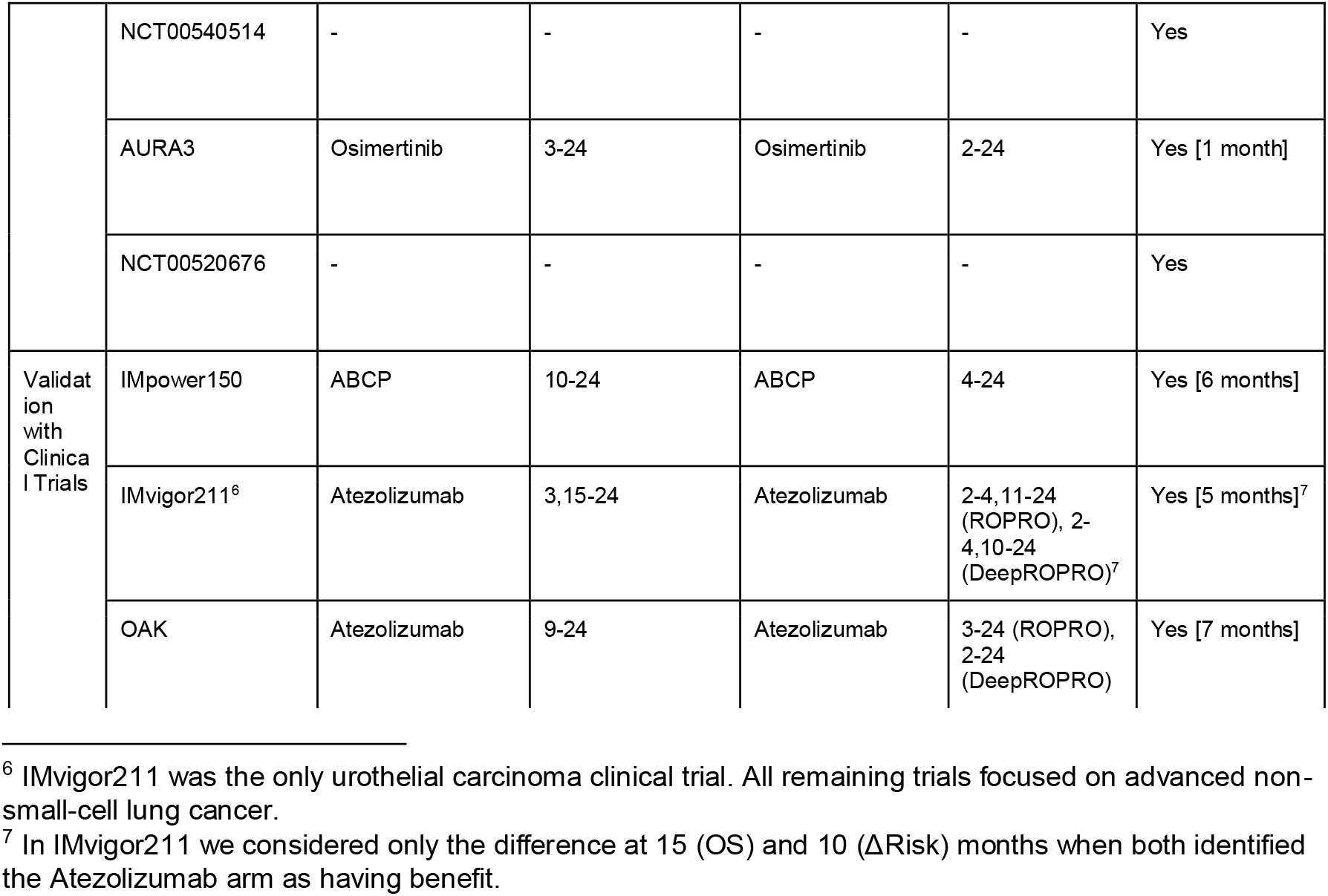
Treatment arms with overall survival (OS) and ΔRisk benefit for the higher caliper, lower caliper and validation with clinical trial analyses.

We followed a similar approach to Liu et al. (Liu et al. 2021) to emulate these 12 clinical trials. For each clinical trial, we selected a cohort of patients from FH that took the same medications as in the original clinical trial. To reduce confounding, we applied propensity score matching on the baseline (before treatment) ROPRO value to generate two treatment arms (treatment and control). For each clinical trial, we produced two distinct emulated datasets: one with a higher, and one with a lower number of matched patients (higher and lower caliper values, respectively). These two datasets mimic clinical trials at later (phase III, more patients) and earlier phases (phase I/II, less patients), allowing us to validate and measure the sensitivity of the ΔRisk framework on a broad range of cohort sizes. In some emulated clinical trials, the treatment arms had more patients than in the original trial. This was a side-effect of the large number of patients in FH for these medications. Any down-sampling attempt to match the patient numbers to the clinical trial would have made the analysis more complex, as we would have to resample many datasets. Our approach with the higher and lower caliper datasets for each emulated clinical trial mitigates this effect. Supplementary information contains more information on the trial emulation.

To verify that our emulated clinical trials are concordant with the original clinical trials, we compared their OS hazard ratio (HR) confidence intervals. If the confidence intervals intersected, we kept the emulated trial for our analysis.

### Clinical trials: additional immunotherapy studies

We included three additional datasets derived from recent phase III Roche clinical trials to further validate our ΔRisk framework, and to analyze both ΔRisk and PFS as early OS predictors. IMpower150 (Socinski et al. 2018; 2021), and OAK (Rittmeyer et al. 2017) focused on aNSCLC, while IMvigor211 (Powles et al. 2018; van der Heijden et al. 2021), focused on metastatic urothelial carcinoma. We analyzed the data from the first cut-off of IMvigor211 (Powles et al. 2018), since it was the latest data available to us at the time of this analysis. From the IMpower150, we selected the Bevacizumab+Carboplatin+Paclitaxel (BCP) (389 patients) and Atezolizumab+BCP (ABCP) (387 patients) arms because these arms were focused on the original publication (Socinski et al. 2018; 2021). From OAK and IMvigor211, respectively, we analyzed the full dataset composed of 598 and 454 patients who received Atezolizumab, and 564 and 429 patients who received chemotherapy (Docetaxel in OAK, and either Docetaxel, Vinflunine or Paclitaxel in IMvigor211).

This analysis does not include early-phase clinical trials because we did not have access to any clinical trial data with a control group. Instead, to analyze the performance of the ΔRisk framework in settings with a lower number of patients (similar to an early-phase clinical trial), we used the lower caliper datasets described above.

## Results

### Evaluation and sensitivity of the emulated trials

The datasets of the 12 RWD-emulated clinical trials cover a wide range of patient numbers (from 60 to 2156 patients, the emulated datasets include less patients because of the matching step) per treatment arm (see Supplementary Tables 2-25 for the baseline characteristics). The OS results of the emulated and original clinical trials were consistent in the majority of trials (Supplementary Figure 2). The only differences in OS between emulated and original clinical trials were: (1) a significantly higher OS benefit in the emulated versus original clinical trials for the Afatinib and Pembrolizumab treatments of the LUX-Lung 3+6 (higher caliper), and KEYNOTE-042 (lower caliper) clinical trials, respectively. And, (2) loss of OS statistical significance in the lower caliper dataset of KEYNOTE-407. This loss of significance was likely due to the lower number of patients (342 patients, ∼61% of the sample size of the original trial) when compared with the original clinical trial (559 patients) (Paz-Ares et al. 2018).

### ΔRisk evaluation on 12 emulated clinical trials

#### ΔRisk concordance with OS (Hypothesis 1)

In 11 out of 12 clinical trials, the ΔROPRO and ΔDeepROPRO trends were concordant with the original clinical trial’s OS for both caliper values (refer to Table 1 for a summary of the results). Out of the 12 clinical trials, 8 had a significant difference in OS. For all of these 8 clinical trials, the ΔROPRO/ΔDeepROPRO trends were associated with OS, i.e. the treatment arm with the higher ΔROPRO/ΔDeepROPRO trend had always the lower OS. For the remaining 4 clinical trials, the ΔROPRO/ΔDeepROPRO were concordant with OS in all clinical trials except one (PRONOUNCE). In PRONOUNCE, the Carboplatin+Pemetrexed treatment arm had significantly higher ΔROPRO and ΔDeepROPRO values, although there was no difference in OS between the arms in the original study (see Supplementary Figure 7). Complete results for each emulated clinical trial can be found in Supplementary Figures 3-26, and Supplementary Table 26-49.

#### ΔRisk identifies treatment difference before conventional methods (Hypothesis 2)

The ΔRisk statistical test strongly outperformed the log-rank test in predicting the difference between the treatment arms earlier (up to 5 months) for 5 out of 8 clinical trials in the higher caliper analysis (at the same time for the remaining 3 trials), and for 7 out of 7 clinical trials for the lower caliper analysis. KEYNOTE-407 was not considered in the lower caliper analysis because no significant difference in OS was detected. Strikingly, the ΔRisk statistical test identified the difference between the treatment arms before or at the same time as the OS log-rank test for all emulated clinical trials. Furthermore, on average, the ΔRisk statistical test identified the difference between the treatment arms 1.75, and 1.86 months before the OS log-rank test for the higher and lower calipers, respectively (Table 1 contains a summary of the results).

### ΔRisk evaluation in 3 additional immunotherapy studies

#### ΔRisk concordance with OS (Hypothesis 1)

To validate our findings from the RWD analysis, we analyzed the ΔRisk performance in three Roche immunotherapy clinical trials (baseline characteristics of each cohort in the Supplementary Tables 50-52). The ΔROPRO/ΔDeepROPRO were also concordant with OS in all three Roche clinical trials (see Table 1 for a summary of the results). Figure 2C, the Supplementary Figures 27-29, and the model coefficients in Supplementary Tables 53-55 show an improvement in both OS (higher survival) and ΔROPRO/ΔDeepROPRO (lower mortality risk) for the treatment arms containing Atezolizumab in all clinical trials.

#### ΔRisk identifies treatment difference before conventional methods (Hypothesis 2)

In addition, the ΔRisk statistical test detected the difference between the treatment arms before the OS log-rank test in all cases (IMpower150: 6 months before; OAK: 7 months before). For IMvigor211 (Supplementary Figures 28), there was a crossover in the OS curves between 4 and 8 months. The ΔROPRO and ΔDeepROPRO detected the crossover and, interestingly, at 6 months (two months before any visual difference in OS) there was already a clear ΔROPRO/ΔDeepROPRO improvement for the Atezolizumab treatment arm. After the crossover, both the log-rank and ΔROPRO/ΔDeepROPRO (up to 5 months before the log-rank test) tests identified an improvement in the Atezolizumab treatment arm. In summary, for these three clinical trials, the ΔROPRO/ΔDeepROPRO detected the treatment benefit, on average 5.67 (ROPRO) and 6 (DeepROPRO) months before the OS log-rank test.

#### ΔRisk identifies treatment difference before PFS (Hypothesis 3)

In contrast to the RWD analysis (for which progression data was not available), we could evaluate the performance of PFS as an early estimator of OS in these three Roche clinical trials. Interestingly, the early PFS was only concordant with OS in the IMpower150 clinical trial. The ABCP treatment arm had higher PFS and OS (significant difference from 5 and 10 months onward, respectively). Inversely, for both IMvigor211 and OAK, the early PFS was opposite to the OS. The Atezolizumab treatment arms had lower PFS in the earlier months of the clinical trial (IMvigor211: significant difference 3-6 months, OAK: significant difference 2-4 months), i.e, there were more progression events in the Atezolizumab treatment arms than in the control arm. Still, the Atezolizumab treatment arms showed the highest OS benefit. Therefore, the increase in progression events (which, at least in part, could possibly be explained by TFR) was not translated into an increase in mortality. Strikingly, in these cases, the early PFS would have failed as an early surrogate endpoint, while the ΔROPRO/ΔDeepROPRO would have correctly identified the survival difference between the treatment arms.

### ΔRisk correlation with final OS hazard ratio (Hypothesis 4)

As a last analysis of the ΔROPRO/ΔDeepROPRO performance, we explored the correlation between the early JM coefficients and OS at late cut-offs. Remarkably, the JM coefficients at 3 months were already highly correlated with two year OS (ΔROPRO *R*^2^ values [confidence interval], *β*_2_ and *γ*: 0.91 [0.73, 0.98], only *β*_2_: 0.91 [0.68, 0.98]). Even at the last time-point considered (4 years), the correlation with the 3 month JM coefficients was still high (ΔROPRO *R*^2^ values [confidence interval], *β*_2_ and *γ*: 0.87 [0.66, 0.97], only *β*_2_: 0.84 [0.56, 0.96]). Therefore, the early JM coefficients were highly correlated with OS measurements ∼2 and ∼4 years in the future. The high correlation demonstrates a high trial-level “simple surrogacy” (the full correlation results are available in Supplementary Tables 56-58, and Supplementary Figures 33-34).

## Discussion

We introduce ΔRisk, as a potential R&D decision-support tool built on top of routinely collected clinical parameters, and of the ROPRO and DeepROPRO prognostic scores. Additionally, we present a statistical test leveraging the joint model framework to identify differences in ΔRisk between treatment arms, which can be used to detect early treatment benefit in oncology clinical trials. Our results show that the ΔRisk trend (for ROPRO and DeepROPRO) was comprehensively concordant with the final OS, in 11 out of 12 RWD-emulated clinical trials, and in all three recent phase III cancer immunotherapy clinical trials from Roche. The ΔRisk statistical test detected the difference between the treatment arms, on average, ∼2 (RWD-emulated trials) and ∼6 months (real clinical trials) before the log-rank test for OS. Additionally, the early ΔRisk coefficients (from as early as 3 months) were predictive of OS at 4 years.

Strikingly, there were two cases where early PFS would have failed as an early OS estimator but our ΔROPRO/ΔDeepROPRO correctly identified the difference between the treatment arms. Thus, we conclude that the ΔRisk framework has potential to be a R&D decision-support tool.

The ΔRisk framework showed a strong performance even when the sample size was relatively low (lower caliper analysis, some datasets had only ∼100 patients). First, the ΔRisk consistently predicted the treatment arms with OS benefit in the lower caliper analysis for all emulated clinical trials except one (PRONOUNCE). Second, the ΔRisk statistical test had only a small performance loss when the cohort sizes were reduced (from higher to lower caliper).

Specifically, for the majority of clinical trials (6 out of 8), the ΔRisk statistical test detected the treatment benefit at the same time across higher and lower caliper analyses (compared with 3 out of 8 for the standard log-rank test), suggesting that ΔRisk outperforms standard methods to detect OS benefit early. Finally, the KEYNOTE-407 analysis further illustrated the power of ΔRisk in earlier clinical trial phases. An OS benefit for the Pembrolizumab-combo treatment arm was detected in the original trial (559 patients) (Paz-Ares et al. 2018), and the higher caliper analysis (1174 patients), but not in the lower caliper analysis (342 patients, closer to a phase II in cohort size). In contrast, the ΔRisk consistently identified an improvement in the Pembrolizumab-combo treatment arm across caliper values, indicating that ΔRisk could identify medication benefit even in smaller cohort sizes and shorter observation time. All these data point out that ΔRisk may be particularly useful for decision making in early clinical trials where the number of patients is low and decisions need to be taken early on.

The only clinical trial for which the ΔRisk was not concordant with OS was our emulated PRONOUNCE trial, originally a randomized phase III trial investigating Carboplatin+Pemetrexed (Arm A) in advanced nonsquamous NSCLC patients. Arm A revealed the highest ΔRisk trend, although none of the treatment arms showed OS benefit in the original trial (Zinner et al. 2015). We hypothesize that this difference results from the higher number of grade 3-4 anemia and thrombocytopenia adverse events detected on Arm A in the original (Zinner et al. 2015) and our RWD-emulated clinical trials (Supplementary Figures 30-31). Since ROPRO and DeepROPRO use hemoglobin and platelet levels, their abnormal values could increase the ΔRisk of the patients, thus influencing the safety profile of the drugs but not translating into more deaths.

Hence, in cases where the treatment strongly influences blood parameters of the ROPRO/DeepROPRO models, the ΔRisk needs to be analyzed carefully. Still, higher adverse event cases did not always lead to incorrect ΔROPRO/ΔDeepROPRO predictions. For instance, the Pembrolizumab and Carboplatin+nab-Paclitaxel treatment arms of the KEYNOTE-189 and NCT00540514 clinical trials, respectively, had a significantly higher number of grade 3-4 anemia cases (also influencing ROPRO blood parameters). Still, in these two emulated clinical trials the ΔRisk statistical test correctly identified the observed OS benefit, independently of the higher adverse event numbers.

### ΔRisk in cancer immunotherapy

We analyzed 7 cancer immunotherapy clinical trials (4 with Pembrolizumab emulated with RWD, and 3 with Atezolizumab). The ΔROPRO/ΔDeepROPRO consistently identified the treatment arm with OS benefit in all cases. Additionally, the ΔRisk statistical test identified the difference between the arms earlier than the log-rank test [higher caliper: 1.25; lower caliper: 2.67; Atezolizumab analysis: 5.67 (ROPRO) and 6 (DeepROPRO) months before]. In addition, our analysis confirmed the lack of correlation between PFS and OS, possibly due to tumor flare reaction (Mushti, Mulkey, and Sridhara 2018; Ye et al. 2020), in two clinical trials (IMvigor211 and OAK). Only in the IMpower150 trial, was PFS associated with OS. Hence, in two trials, PFS would have failed to predict the treatment benefit. Unlike PFS, our ΔRisk framework correctly identified the treatment arms with final OS benefit, and did so before the OS difference could be detected. We hypothesize that the routine measurements of many parameters of the ROPRO allow us to measure the patients’ performance status more comprehensively, thus giving a better reflection of OS benefit earlier on. The possible lower performance of PFS, and the strong performance of the ΔRisk in cancer immunotherapy provides evidence that the ΔRisk could be a valuable R&D decision-support tool in cancer immunotherapy.

### Potential future applications of the ΔRisk

Based on our analysis we see a value of applying the ΔRisk in early (RWD lower caliper) as well as late phase trials (RWD higher caliper and Roche trials). In both settings, the ΔRisk analysis could provide an estimate of the OS benefit early on. The RWD lower caliper analysis illustrated how the ΔRisk could be useful in early clinical trial phases (phase I/II). Specifically, the ΔRisk estimate could serve as an initial indicator of the potential OS benefit in subsequent confirmatory trials. The initial ΔRisk modeling could be performed whenever enough data is available. In the absence of a control arm, a suitable control arm could be emulated by matching patients from RWD or historical clinical trials. Still, further analyses are required in clinical trial data to validate this hypothesis. Additionally, as exemplified by the RWD higher caliper and Roche trials, in later phases (e.g., phase III), the ΔRisk modeling could be performed at the first interim analysis, to inform a futility analysis of the treatment that could further guide the clinical trial team. In summary, the ΔRisk could serve as a valuable additional R&D support tool.

Another interesting but yet to be confirmed application may be to analyze how individual patients are responding to medications. The ROPRO longitudinal analysis may potentially help physicians in taking treatment decisions. However, more validation work would be needed to test ΔROPRO/ΔDeepROPRO on a patient level.

### Limitations

First, we focused on aNSCLC, one of the most common types of cancer and the largest cohort in Flatiron Health. Therefore, further analyses are required to validate the framework in other cancer indications. Nevertheless, from our experience with the baseline pan-cancer ROPRO and our analysis in urothelial carcinoma (IMvigor211), we believe ΔROPRO will perform equally well in most indications. Second, the majority of our analyses were conducted using RWD because it gave us the flexibility to test ΔRisk in a broad range of medication types and number of patients. Given the differences between RWD and clinical trial data (e.g., higher missingness), the results from RWD might not be directly translatable to clinical trials. To account for this, we validated our RWD results in three clinical trials, still, more validation will be needed, especially in early clinical trial phases. Third, we could not analyze PFS in the RWD-emulated clinical trials and thus our results on PFS are only based on the three clinical trials.

Fourth, our analysis is biased towards trials testing efficacious medication as enough patients could only be found in the Flatiron Health database for these medications. We plan to further investigate the ΔRisk framework in clinical trials of drugs that failed to show efficacy.

Further, as this is a first analysis of the ΔRisk framework, we did not prove its surrogacy to OS according to the guidelines introduced by Prentice (Prentice 1989). There are several reasons why we decided not to attempt a formal surrogate endpoint validation at this point. First, we felt that a more informal exploration as presented here is needed to trigger respective interest and discussion, before it is possible to enter into a formal validation process. Second, for a formal validation a broader range of clinical studies has to be investigated and further institutions, both from academia and pharmaceutical industry, have to be involved. We started with a decent data basis that allowed the emulation of a series of hallmark studies and added further relevant in-house studies from Roche. Still, further sources are indispensable to achieve proof of generalizability. Finally, some statistical methodology work needs to be done to establish a basis for using joint models within the established framework for surrogate endpoint validation (Alonso et al. 2016; Burzykowski, Molenberghs, and Buyse 2005). In our analyses here, we already show study-level association between the surrogate and final OS (Supplementary Tables 58-63). Study-level association can be computed easily also when two parameters per study are estimated, as we do as one analysis alternative in the joint model. Evaluation of patient-level association, however, is more complex. In contrast to typical surrogate validation where two different time-to-event categories (e.g. PFS and OS) are associated, the estimates we get from the joint model are of a different type than the classic OS estimates. Therefore, we plan to do respective methodological work in the future. In this context, it will be of particular relevance to investigate potential interaction between mode of action of a drug and parameters of the ROPRO. Such direct interaction seems to be likely for chemotherapies, as also the PRONOUNCE trial suggests. As a consequence, ROPRO would not only measure overall patient fitness, but would be confounded by treatment burden. For cancer immunotherapy or targeted therapies we would expect such interplay to be less important or even negligible. In any case, it will be necessary to investigate the sensitivity of our approach to effects intrinsic to different treatment classes. As we are expecting to see more trials which compare different cancer immunotherapy treatments against each other, there should be a growing range of applicability of our framework in the future. Lastly, we tested the ΔRisk with ROPRO and DeepROPRO, which are composed mainly of vital and blood-test parameters. There are other, independent, prognostic biomarkers that were not included in these models (cancer-specific biomarkers, circulating tumor DNA [ctDNA], C-reactive protein [CRP], patient reported outcomes [PROs], cell surface markers, etc.). These can be investigated in their own right, using the ΔRisk framework we exemplified here, or be combined into a joined score to further increase the performance.

## Conclusion

Trustworthy estimates of Overall Survival (OS) are essential for high quality decision-making in clinical trials. Our results show that the ΔRisk framework (with ROPRO and DeepROPRO) was able to predict treatment benefit for the majority of clinical trials studied. Importantly, the ΔRisk could identify the difference between the clinical trial arms significantly earlier than OS readout, and thus could aid early clinical trial decisions. In addition, our analysis suggests that ΔROPRO/ΔDeepROPRO is less susceptible to tumor flare reaction (TFR) compared to early PFS and thus can identify treatment benefits in cases where PFS fails as an early surrogate endpoint.

The results in this initial analysis of the ΔRisk framework imply that it could be used as an additional R&D decision-support tool in advanced non-small-cell cancer (aNSCLC) clinical trials, and potentially beyond as suggested by our urothelial carcinoma (IMvigor211) analysis. Additional analyses on OS correlation, on other cancer indications and even other diseases should be made to further validate the ΔRisk framework.

## Data Availability

The data presented is owned by either Flatiron Health Inc or F. Hoffmann-la Roche LTD. Access to Flatiron Health may be made available upon request, and are subject to a license agreement with Flatiron Health. Requests to access these datasets should be directed to.

## Acknowledgements

The authors thank Carlos Talavera-López (Institute for Computational Health, Helmholtz Munich), Fabian Schmich (Roche Innovation Center Munich) and Bruno Gomes (Roche Innovation Center Basel) for their valuable input.

HL is supported by the Helmholtz Association under the joint research school “Munich School for Data Science - MUDS”.

