## Supplementary Figures and Tables for "Longitudinal assessment of ROPRO as an early indicator of overall survival in oncology clinical trials: a retrospective analysis": Population characteristics.docx

### 1. Population characteristics

The tables below contain the baseline characteristics of the patients in each of the real-world data emulated clinical trials. Note that the emulated datasets include less patients than the original Flatiron Health cohorts because of the propensity score matching. In the matching step only some patients are kept, hence the lower numbers.

#### 1.1. Higher caliper population characteristics

Supplementary Table 2: Baseline values for the emulated KEYNOTE-189 clinical trial.

|  | Platinum + Pembrolizumab + Pemetrexed | Platinum + Pemetrexed |
| --- | --- | --- |
| Number of Patients | 1691 | 1691 |
| ROPRO (mean (SD)) | 0.17 (0.36) | 0.17 (0.36) |
| Age at baseline [years] (mean (SD)) | 67.59 (9.46) | 66.89 (9.26) |
| Gender = Male (mean (SD)) | 0.54 (0.50) | 0.52 (0.50) |
| History of Smoking = Yes (mean (SD)) | 0.90 (0.30) | 0.91 (0.28) |
| Number of metastatic sites (mean (SD)) | 0.65 (0.90) | 0.24 (0.57) |
| Hemoglobin [mass/volume] in blood (mean (SD)) | 11.82 (1.68) | 11.76 (1.72) |
| Urea nitrogen [mass/volume] in serum or plasma (mean (SD)) | 15.99 (5.61) | 16.35 (5.67) |
| Platelets [#/volume] in blood (mean (SD)) | 249.27 (83.01) | 241.32 (83.16) |
| Calcium [mass/volume] in serum or plasma (mean (SD)) | 9.17 (0.48) | 9.16 (0.48) |
| Glucose [mass/volume] in serum or plasma (mean (SD)) | 118.02 (24.23) | 118.09 (25.57) |
| Lymphocytes/100 leukocytes in blood (mean (SD)) | 17.97 (8.24) | 17.90 (8.65) |
| Alkaline phosphatase [enzymatic activity/volume] in serum or plasma (mean (SD)) | 101.79 (32.58) | 100.21 (31.13) |
| Protein [mass/volume] in serum or plasma (mean (SD)) | 66.86 (6.02) | 66.71 (5.87) |
| Alanine aminotransferase [enzymatic activity/volume] in serum or plasma (mean (SD)) | 23.83 (10.96) | 24.03 (11.12) |
| Albumin [mass/volume] in serum or plasma (mean (SD)) | 36.99 (4.17) | 36.75 (4.25) |
| Bilirubin.total [mass/volume] in serum or plasma (mean (SD)) | 0.42 (0.17) | 0.42 (0.18) |
| Chloride [moles/volume] in serum or plasma (mean (SD)) | 100.80 (3.20) | 100.73 (2.96) |
| Monocytes [#/volume] in blood (mean (SD)) | 0.59 (0.22) | 0.56 (0.22) |
| Eosinophils/100 leukocytes in blood (mean (SD)) | 1.71 (1.12) | 1.70 (0.97) |
| Lactate dehydrogenase [enzymatic activity/volume] in serum or plasma (mean (SD)) | 210.55 (51.20) | 209.03 (49.49) |
| Heart rate (mean (SD)) | 87.50 (14.19) | 86.33 (13.72) |
| Systolic blood pressure (mean (SD)) | 125.45 (15.36) | 124.63 (14.96) |
| Oxygen saturation in arterial blood by pulse oximetry (mean (SD)) | 96.90 (1.13) | 96.92 (0.97) |
| ECOG (mean (SD)) | 0.82 (0.60) | 0.91 (0.54) |
| NLR (mean (SD)) | 3.73 (1.56) | 3.84 (1.58) |
| BMI (mean (SD)) | 26.03 (5.16) | 25.92 (5.23) |
| Tumor stage at baseline (mean (SD)) | 3.72 (0.78) | 3.57 (0.86) |
| AST/ALT ratio [%] (mean (SD)) | 1.09 (0.39) | 1.08 (0.38) |
| Histology = Non-squamous cell carcinoma (%) | 1691 (100.0) | 1691 (100.0) |

Supplementary Table 3: Baseline values for the emulated KEYNOTE-024 clinical trial.

|  | Pembrolizumab | Platinum-doublet (5 types) |
| --- | --- | --- |
| Number of Patients | 2156 | 2156 |
| ROPRO (mean (SD)) | 0.20 (0.41) | 0.20 (0.41) |
| Age at baseline [years] (mean (SD)) | 71.90 (9.24) | 67.57 (9.15) |
| Gender = Male (mean (SD)) | 0.49 (0.50) | 0.58 (0.49) |
| History of Smoking = Yes (mean (SD)) | 0.93 (0.26) | 0.93 (0.25) |
| Number of metastatic sites (mean (SD)) | 0.54 (0.83) | 0.19 (0.55) |
| Hemoglobin [mass/volume] in blood (mean (SD)) | 12.15 (1.76) | 11.54 (1.66) |
| Urea nitrogen [mass/volume] in serum or plasma (mean (SD)) | 16.61 (6.10) | 16.35 (5.90) |
| Platelets [#/volume] in blood (mean (SD)) | 265.40 (80.53) | 242.50 (80.36) |
| Calcium [mass/volume] in serum or plasma (mean (SD)) | 9.24 (0.49) | 9.14 (0.49) |
| Glucose [mass/volume] in serum or plasma (mean (SD)) | 115.49 (25.08) | 116.65 (25.63) |
| Lymphocytes/100 leukocytes in blood (mean (SD)) | 17.39 (7.16) | 17.46 (8.46) |
| Alkaline phosphatase [enzymatic activity/volume] in serum or plasma (mean (SD)) | 94.42 (29.75) | 94.59 (30.64) |
| Protein [mass/volume] in serum or plasma (mean (SD)) | 67.70 (6.18) | 66.70 (6.01) |
| Alanine aminotransferase [enzymatic activity/volume] in serum or plasma (mean (SD)) | 19.01 (10.10) | 22.42 (10.80) |
| Albumin [mass/volume] in serum or plasma (mean (SD)) | 36.75 (4.41) | 36.39 (4.34) |
| Bilirubin.total [mass/volume] in serum or plasma (mean (SD)) | 0.45 (0.18) | 0.43 (0.18) |
| Chloride [moles/volume] in serum or plasma (mean (SD)) | 100.86 (3.34) | 100.81 (3.11) |
| Monocytes [#/volume] in blood (mean (SD)) | 0.63 (0.20) | 0.51 (0.22) |
| Eosinophils/100 leukocytes in blood (mean (SD)) | 2.24 (1.24) | 1.78 (1.00) |
| Lactate dehydrogenase [enzymatic activity/volume] in serum or plasma (mean (SD)) | 202.88 (45.42) | 206.10 (51.92) |
| Heart rate (mean (SD)) | 84.75 (14.14) | 87.23 (13.91) |
| Systolic blood pressure (mean (SD)) | 125.75 (16.01) | 122.56 (14.66) |
| Oxygen saturation in arterial blood by pulse oximetry (mean (SD)) | 96.77 (1.11) | 96.90 (0.98) |
| ECOG (mean (SD)) | 0.97 (0.63) | 0.93 (0.56) |
| NLR (mean (SD)) | 3.81 (1.47) | 3.81 (1.51) |
| BMI (mean (SD)) | 25.69 (5.35) | 25.96 (5.50) |
| Tumor stage at baseline (mean (SD)) | 3.38 (1.04) | 3.45 (0.88) |
| AST/ALT ratio [%] (mean (SD)) | 1.19 (0.42) | 1.12 (0.38) |
| Histology = Squamous cell carcinoma (%) | 626 (29.0) | 820 (38.0) |

Supplementary Table 4: Baseline values for the emulated KEYNOTE-042 clinical trial.

|  | Pembrolizumab | Platinum-doublet (2 types) |
| --- | --- | --- |
| Number of Patients | 2064 | 2064 |
| ROPRO (mean (SD)) | 0.20 (0.40) | 0.20 (0.40) |
| Age at baseline [years] (mean (SD)) | 71.98 (9.23) | 67.62 (9.10) |
| Gender = Male (mean (SD)) | 0.50 (0.50) | 0.57 (0.50) |
| History of Smoking = Yes (mean (SD)) | 0.93 (0.25) | 0.93 (0.25) |
| Number of metastatic sites (mean (SD)) | 0.53 (0.83) | 0.20 (0.55) |
| Hemoglobin [mass/volume] in blood (mean (SD)) | 12.16 (1.75) | 11.56 (1.66) |
| Urea nitrogen [mass/volume] in serum or plasma (mean (SD)) | 16.58 (6.09) | 16.40 (5.89) |
| Platelets [#/volume] in blood (mean (SD)) | 265.13 (80.29) | 244.05 (80.23) |
| Calcium [mass/volume] in serum or plasma (mean (SD)) | 9.24 (0.49) | 9.14 (0.49) |
| Glucose [mass/volume] in serum or plasma (mean (SD)) | 115.46 (25.02) | 116.40 (25.60) |
| Lymphocytes/100 leukocytes in blood (mean (SD)) | 17.36 (7.07) | 17.09 (8.19) |
| Alkaline phosphatase [enzymatic activity/volume] in serum or plasma (mean (SD)) | 94.22 (29.66) | 93.68 (30.52) |
| Protein [mass/volume] in serum or plasma (mean (SD)) | 67.70 (6.17) | 66.58 (6.03) |
| Alanine aminotransferase [enzymatic activity/volume] in serum or plasma (mean (SD)) | 19.00 (10.09) | 22.23 (10.82) |
| Albumin [mass/volume] in serum or plasma (mean (SD)) | 36.74 (4.38) | 36.39 (4.33) |
| Bilirubin.total [mass/volume] in serum or plasma (mean (SD)) | 0.45 (0.18) | 0.43 (0.18) |
| Chloride [moles/volume] in serum or plasma (mean (SD)) | 100.88 (3.34) | 100.89 (3.11) |
| Monocytes [#/volume] in blood (mean (SD)) | 0.63 (0.20) | 0.51 (0.22) |
| Eosinophils/100 leukocytes in blood (mean (SD)) | 2.25 (1.24) | 1.80 (1.01) |
| Lactate dehydrogenase [enzymatic activity/volume] in serum or plasma (mean (SD)) | 203.05 (45.38) | 206.01 (51.65) |
| Heart rate (mean (SD)) | 84.76 (14.16) | 87.50 (13.86) |
| Systolic blood pressure (mean (SD)) | 125.76 (15.97) | 122.41 (14.84) |
| Oxygen saturation in arterial blood by pulse oximetry (mean (SD)) | 96.77 (1.11) | 96.91 (0.97) |
| ECOG (mean (SD)) | 0.97 (0.62) | 0.94 (0.56) |
| NLR (mean (SD)) | 3.81 (1.47) | 3.81 (1.51) |
| BMI (mean (SD)) | 25.72 (5.33) | 25.93 (5.53) |
| Tumor stage at baseline (mean (SD)) | 3.40 (1.03) | 3.45 (0.88) |
| AST/ALT ratio [%] (mean (SD)) | 1.20 (0.42) | 1.12 (0.38) |
| Histology = Squamous cell carcinoma (%) | 601 (29.1) | 693 (33.6) |

Supplementary Table 5: Baseline values for the emulated KEYNOTE-407 clinical trial.

|  | Carboplatin + Paclitaxel | Carboplatin + Paclitaxel+Pembrolizumab |
| --- | --- | --- |
| Number of Patients | 587 | 587 |
| ROPRO (mean (SD)) | 0.23 (0.34) | 0.23 (0.34) |
| Age at baseline [years] (mean (SD)) | 69.21 (8.14) | 69.12 (8.81) |
| Gender = Male (mean (SD)) | 0.64 (0.48) | 0.66 (0.47) |
| History of Smoking = Yes (mean (SD)) | 0.96 (0.18) | 0.97 (0.18) |
| Number of metastatic sites (mean (SD)) | 0.14 (0.45) | 0.51 (0.84) |
| Hemoglobin [mass/volume] in blood (mean (SD)) | 11.35 (1.62) | 11.33 (1.67) |
| Urea nitrogen [mass/volume] in serum or plasma (mean (SD)) | 15.97 (5.76) | 15.82 (5.54) |
| Platelets [#/volume] in blood (mean (SD)) | 245.12 (79.71) | 244.90 (76.55) |
| Calcium [mass/volume] in serum or plasma (mean (SD)) | 9.14 (0.45) | 9.17 (0.44) |
| Glucose [mass/volume] in serum or plasma (mean (SD)) | 118.02 (26.43) | 119.50 (26.45) |
| Lymphocytes/100 leukocytes in blood (mean (SD)) | 17.97 (8.57) | 18.72 (7.86) |
| Alkaline phosphatase [enzymatic activity/volume] in serum or plasma (mean (SD)) | 89.42 (28.02) | 97.37 (31.01) |
| Protein [mass/volume] in serum or plasma (mean (SD)) | 66.83 (6.05) | 67.55 (6.09) |
| Alanine aminotransferase [enzymatic activity/volume] in serum or plasma (mean (SD)) | 19.64 (9.90) | 19.93 (10.03) |
| Albumin [mass/volume] in serum or plasma (mean (SD)) | 36.14 (4.17) | 36.36 (4.08) |
| Bilirubin.total [mass/volume] in serum or plasma (mean (SD)) | 0.44 (0.18) | 0.41 (0.16) |
| Chloride [moles/volume] in serum or plasma (mean (SD)) | 100.75 (3.24) | 100.97 (3.32) |
| Monocytes [#/volume] in blood (mean (SD)) | 0.49 (0.21) | 0.55 (0.21) |
| Eosinophils/100 leukocytes in blood (mean (SD)) | 1.83 (0.95) | 1.88 (1.07) |
| Lactate dehydrogenase [enzymatic activity/volume] in serum or plasma (mean (SD)) | 200.89 (52.41) | 199.68 (40.02) |
| Heart rate (mean (SD)) | 86.94 (13.77) | 87.32 (13.65) |
| Systolic blood pressure (mean (SD)) | 120.72 (14.20) | 124.44 (14.78) |
| Oxygen saturation in arterial blood by pulse oximetry (mean (SD)) | 96.77 (1.01) | 96.88 (1.20) |
| ECOG (mean (SD)) | 0.93 (0.58) | 0.88 (0.58) |
| NLR (mean (SD)) | 3.81 (1.53) | 3.72 (1.44) |
| BMI (mean (SD)) | 26.20 (5.58) | 26.14 (5.42) |
| Tumor stage at baseline (mean (SD)) | 3.37 (0.91) | 3.50 (0.90) |
| AST/ALT ratio [%] (mean (SD)) | 1.17 (0.37) | 1.18 (0.40) |
| Histology = Squamous cell carcinoma (%) | 587 (100.0) | 587 (100.0) |

Supplementary Table 6: Baseline values for the emulated PRONOUNCE clinical trial.

|  | Bevacizumab + Carboplatin+Paclitaxel | Carboplatin + Pemetrexed |
| --- | --- | --- |
| Number of Patients | 502 | 502 |
| ROPRO (mean (SD)) | 0.16 (0.33) | 0.16 (0.33) |
| Age at baseline [years] (mean (SD)) | 65.85 (9.07) | 67.19 (9.35) |
| Gender = Male (mean (SD)) | 0.57 (0.50) | 0.54 (0.50) |
| History of Smoking = Yes (mean (SD)) | 0.91 (0.29) | 0.90 (0.29) |
| Number of metastatic sites (mean (SD)) | 0.18 (0.56) | 0.25 (0.67) |
| Hemoglobin [mass/volume] in blood (mean (SD)) | 12.03 (1.70) | 11.79 (1.65) |
| Urea nitrogen [mass/volume] in serum or plasma (mean (SD)) | 16.00 (5.76) | 16.29 (5.70) |
| Platelets [#/volume] in blood (mean (SD)) | 242.82 (75.51) | 243.99 (81.12) |
| Calcium [mass/volume] in serum or plasma (mean (SD)) | 9.08 (0.49) | 9.19 (0.48) |
| Glucose [mass/volume] in serum or plasma (mean (SD)) | 117.20 (25.48) | 116.56 (25.06) |
| Lymphocytes/100 leukocytes in blood (mean (SD)) | 18.01 (8.54) | 18.22 (8.35) |
| Alkaline phosphatase [enzymatic activity/volume] in serum or plasma (mean (SD)) | 101.82 (31.08) | 97.80 (31.33) |
| Protein [mass/volume] in serum or plasma (mean (SD)) | 66.67 (6.22) | 66.81 (5.68) |
| Alanine aminotransferase [enzymatic activity/volume] in serum or plasma (mean (SD)) | 22.03 (10.61) | 24.90 (11.29) |
| Albumin [mass/volume] in serum or plasma (mean (SD)) | 36.61 (4.18) | 36.77 (4.16) |
| Bilirubin.total [mass/volume] in serum or plasma (mean (SD)) | 0.43 (0.19) | 0.42 (0.18) |
| Chloride [moles/volume] in serum or plasma (mean (SD)) | 101.12 (2.99) | 100.74 (2.98) |
| Monocytes [#/volume] in blood (mean (SD)) | 0.55 (0.24) | 0.57 (0.22) |
| Eosinophils/100 leukocytes in blood (mean (SD)) | 1.69 (0.95) | 1.73 (1.01) |
| Lactate dehydrogenase [enzymatic activity/volume] in serum or plasma (mean (SD)) | 208.36 (51.02) | 205.62 (41.44) |
| Heart rate (mean (SD)) | 87.24 (13.99) | 87.01 (14.15) |
| Systolic blood pressure (mean (SD)) | 126.99 (14.50) | 125.55 (14.70) |
| Oxygen saturation in arterial blood by pulse oximetry (mean (SD)) | 96.90 (1.06) | 96.91 (0.89) |
| ECOG (mean (SD)) | 0.95 (0.47) | 0.91 (0.51) |
| NLR (mean (SD)) | 3.82 (1.58) | 3.76 (1.56) |
| BMI (mean (SD)) | 25.75 (4.95) | 26.04 (5.17) |
| Tumor stage at baseline (mean (SD)) | 3.73 (0.70) | 3.63 (0.81) |
| AST/ALT ratio [%] (mean (SD)) | 1.12 (0.39) | 1.07 (0.37) |
| Histology = Non-squamous cell carcinoma (%) | 502 (100.0) | 502 (100.0) |

Supplementary Table 7: Baseline values for the emulated PointBreak clinical trial.

|  | Bevacizumab + Carboplatin+Paclitaxel | Bevacizumab + Carboplatin+Pemetrexed |
| --- | --- | --- |
| Number of Patients | 296 | 296 |
| ROPRO (mean (SD)) | 0.11 (0.31) | 0.11 (0.31) |
| Age at baseline [years] (mean (SD)) | 65.76 (8.89) | 65.53 (9.12) |
| Gender = Male (mean (SD)) | 0.55 (0.50) | 0.52 (0.50) |
| History of Smoking = Yes (mean (SD)) | 0.93 (0.25) | 0.92 (0.27) |
| Number of metastatic sites (mean (SD)) | 0.17 (0.56) | 0.31 (0.74) |
| Hemoglobin [mass/volume] in blood (mean (SD)) | 12.15 (1.65) | 11.87 (1.69) |
| Urea nitrogen [mass/volume] in serum or plasma (mean (SD)) | 15.83 (5.78) | 15.58 (5.52) |
| Platelets [#/volume] in blood (mean (SD)) | 240.48 (77.99) | 234.29 (87.86) |
| Calcium [mass/volume] in serum or plasma (mean (SD)) | 9.11 (0.46) | 9.16 (0.47) |
| Glucose [mass/volume] in serum or plasma (mean (SD)) | 117.29 (26.66) | 117.67 (25.10) |
| Lymphocytes/100 leukocytes in blood (mean (SD)) | 18.19 (7.92) | 18.68 (8.21) |
| Alkaline phosphatase [enzymatic activity/volume] in serum or plasma (mean (SD)) | 100.23 (29.44) | 102.99 (33.60) |
| Protein [mass/volume] in serum or plasma (mean (SD)) | 66.95 (6.12) | 67.40 (6.36) |
| Alanine aminotransferase [enzymatic activity/volume] in serum or plasma (mean (SD)) | 21.92 (10.17) | 24.95 (11.24) |
| Albumin [mass/volume] in serum or plasma (mean (SD)) | 36.82 (3.95) | 37.21 (4.14) |
| Bilirubin.total [mass/volume] in serum or plasma (mean (SD)) | 0.42 (0.19) | 0.42 (0.18) |
| Chloride [moles/volume] in serum or plasma (mean (SD)) | 101.36 (2.94) | 100.70 (3.06) |
| Monocytes [#/volume] in blood (mean (SD)) | 0.55 (0.24) | 0.57 (0.22) |
| Eosinophils/100 leukocytes in blood (mean (SD)) | 1.78 (1.04) | 1.78 (0.84) |
| Lactate dehydrogenase [enzymatic activity/volume] in serum or plasma (mean (SD)) | 204.92 (51.86) | 207.89 (49.42) |
| Heart rate (mean (SD)) | 86.76 (14.17) | 86.74 (13.94) |
| Systolic blood pressure (mean (SD)) | 126.85 (14.91) | 126.48 (14.50) |
| Oxygen saturation in arterial blood by pulse oximetry (mean (SD)) | 96.88 (1.03) | 96.85 (0.90) |
| ECOG (mean (SD)) | 0.89 (0.51) | 0.87 (0.53) |
| NLR (mean (SD)) | 3.87 (1.65) | 3.72 (1.46) |
| BMI (mean (SD)) | 25.98 (4.95) | 25.74 (4.75) |
| Tumor stage at baseline (mean (SD)) | 3.68 (0.78) | 3.68 (0.78) |
| AST/ALT ratio [%] (mean (SD)) | 1.11 (0.39) | 1.04 (0.38) |
| Histology = Non-squamous cell carcinoma (%) | 296 (100.0) | 296 (100.0) |

Supplementary Table 8: Baseline values for the emulated PROFILE 1014 clinical trial.

|  | Carboplatin-Cisplatin + Pemetrexed | Crizotinib |
| --- | --- | --- |
| Number of Patients | 190 | 190 |
| ROPRO (mean (SD)) | 0.11 (0.38) | 0.11 (0.38) |
| Age at baseline [years] (mean (SD)) | 66.23 (10.21) | 66.76 (10.70) |
| Gender = Male (mean (SD)) | 0.54 (0.50) | 0.48 (0.50) |
| History of Smoking = Yes (mean (SD)) | 0.92 (0.26) | 0.58 (0.49) |
| Number of metastatic sites (mean (SD)) | 0.21 (0.57) | 0.35 (0.68) |
| Hemoglobin [mass/volume] in blood (mean (SD)) | 12.07 (1.68) | 12.19 (1.81) |
| Urea nitrogen [mass/volume] in serum or plasma (mean (SD)) | 16.37 (5.83) | 16.99 (5.79) |
| Platelets [#/volume] in blood (mean (SD)) | 241.09 (81.78) | 263.00 (80.21) |
| Calcium [mass/volume] in serum or plasma (mean (SD)) | 9.22 (0.50) | 8.87 (0.52) |
| Glucose [mass/volume] in serum or plasma (mean (SD)) | 113.65 (22.71) | 112.29 (23.80) |
| Lymphocytes/100 leukocytes in blood (mean (SD)) | 17.79 (8.54) | 17.89 (7.47) |
| Alkaline phosphatase [enzymatic activity/volume] in serum or plasma (mean (SD)) | 99.75 (31.03) | 103.74 (30.02) |
| Protein [mass/volume] in serum or plasma (mean (SD)) | 66.59 (5.90) | 63.73 (6.28) |
| Alanine aminotransferase [enzymatic activity/volume] in serum or plasma (mean (SD)) | 24.32 (10.41) | 30.54 (11.96) |
| Albumin [mass/volume] in serum or plasma (mean (SD)) | 36.99 (4.26) | 34.48 (4.79) |
| Bilirubin.total [mass/volume] in serum or plasma (mean (SD)) | 0.43 (0.18) | 0.40 (0.18) |
| Chloride [moles/volume] in serum or plasma (mean (SD)) | 100.91 (2.98) | 101.60 (3.10) |
| Monocytes [#/volume] in blood (mean (SD)) | 0.57 (0.21) | 0.64 (0.23) |
| Eosinophils/100 leukocytes in blood (mean (SD)) | 1.77 (1.02) | 2.31 (1.29) |
| Lactate dehydrogenase [enzymatic activity/volume] in serum or plasma (mean (SD)) | 206.73 (44.44) | 213.86 (59.58) |
| Heart rate (mean (SD)) | 83.29 (13.59) | 79.69 (15.25) |
| Systolic blood pressure (mean (SD)) | 124.95 (14.33) | 122.91 (13.95) |
| Oxygen saturation in arterial blood by pulse oximetry (mean (SD)) | 96.98 (0.85) | 96.89 (0.90) |
| ECOG (mean (SD)) | 0.89 (0.57) | 0.97 (0.59) |
| NLR (mean (SD)) | 3.82 (1.62) | 3.88 (1.62) |
| BMI (mean (SD)) | 26.18 (5.10) | 26.79 (5.47) |
| Tumor stage at baseline (mean (SD)) | 3.54 (0.88) | 3.71 (0.72) |
| AST/ALT ratio [%] (mean (SD)) | 1.05 (0.38) | 1.00 (0.30) |
| Histology = Non-squamous cell carcinoma (%) | 190 (100.0) | 190 (100.0) |

Supplementary Table 9: Baseline values for the emulated FLAURA clinical trial.

|  | Erlotinib-Gefitinib | Osimertinib |
| --- | --- | --- |
| Number of Patients | 363 | 363 |
| ROPRO (mean (SD)) | -0.04 (0.36) | -0.04 (0.36) |
| Age at baseline [years] (mean (SD)) | 69.16 (10.15) | 69.41 (10.66) |
| Gender = Male (mean (SD)) | 0.33 (0.47) | 0.32 (0.47) |
| History of Smoking = Yes (mean (SD)) | 0.55 (0.49) | 0.47 (0.50) |
| Number of metastatic sites (mean (SD)) | 0.23 (0.58) | 0.71 (0.98) |
| Hemoglobin [mass/volume] in blood (mean (SD)) | 12.57 (1.56) | 12.26 (1.62) |
| Urea nitrogen [mass/volume] in serum or plasma (mean (SD)) | 16.70 (5.67) | 16.53 (5.91) |
| Platelets [#/volume] in blood (mean (SD)) | 256.68 (71.42) | 215.28 (66.48) |
| Calcium [mass/volume] in serum or plasma (mean (SD)) | 9.28 (0.48) | 9.09 (0.49) |
| Glucose [mass/volume] in serum or plasma (mean (SD)) | 110.97 (23.75) | 111.20 (22.81) |
| Lymphocytes/100 leukocytes in blood (mean (SD)) | 19.58 (7.98) | 18.70 (7.52) |
| Alkaline phosphatase [enzymatic activity/volume] in serum or plasma (mean (SD)) | 94.38 (30.25) | 94.02 (33.56) |
| Protein [mass/volume] in serum or plasma (mean (SD)) | 67.15 (5.70) | 66.78 (5.83) |
| Alanine aminotransferase [enzymatic activity/volume] in serum or plasma (mean (SD)) | 21.45 (10.20) | 20.05 (10.10) |
| Albumin [mass/volume] in serum or plasma (mean (SD)) | 38.15 (3.73) | 37.81 (4.02) |
| Bilirubin.total [mass/volume] in serum or plasma (mean (SD)) | 0.56 (0.21) | 0.45 (0.18) |
| Chloride [moles/volume] in serum or plasma (mean (SD)) | 101.80 (2.99) | 101.58 (2.95) |
| Monocytes [#/volume] in blood (mean (SD)) | 0.57 (0.18) | 0.55 (0.18) |
| Eosinophils/100 leukocytes in blood (mean (SD)) | 2.22 (1.21) | 2.25 (1.39) |
| Lactate dehydrogenase [enzymatic activity/volume] in serum or plasma (mean (SD)) | 204.76 (47.04) | 203.44 (38.85) |
| Heart rate (mean (SD)) | 84.90 (13.51) | 81.54 (12.86) |
| Systolic blood pressure (mean (SD)) | 128.04 (16.18) | 128.05 (17.13) |
| Oxygen saturation in arterial blood by pulse oximetry (mean (SD)) | 96.98 (0.79) | 96.88 (1.06) |
| ECOG (mean (SD)) | 0.96 (0.53) | 0.86 (0.60) |
| NLR (mean (SD)) | 3.65 (1.40) | 3.67 (1.43) |
| BMI (mean (SD)) | 25.65 (5.23) | 25.80 (4.79) |
| Tumor stage at baseline (mean (SD)) | 3.47 (0.94) | 3.64 (0.84) |
| AST/ALT ratio [%] (mean (SD)) | 1.14 (0.35) | 1.15 (0.41) |
| Histology = Squamous cell carcinoma (%) | 27 (7.4) | 7 (1.9) |

Supplementary Table 10: Baseline values for the emulated LUX-Lung 3+6 clinical trial.

|  | Afatinib | Platinum-doublet (3 types) |
| --- | --- | --- |
| Number of Patients | 253 | 253 |
| ROPRO (mean (SD)) | 0.07 (0.36) | 0.07 (0.36) |
| Age at baseline [years] (mean (SD)) | 70.19 (10.20) | 66.74 (8.95) |
| Gender = Male (mean (SD)) | 0.34 (0.48) | 0.55 (0.50) |
| History of Smoking = Yes (mean (SD)) | 0.56 (0.50) | 0.94 (0.24) |
| Number of metastatic sites (mean (SD)) | 0.50 (0.83) | 0.19 (0.47) |
| Hemoglobin [mass/volume] in blood (mean (SD)) | 12.15 (1.48) | 11.88 (1.62) |
| Urea nitrogen [mass/volume] in serum or plasma (mean (SD)) | 17.15 (6.08) | 16.87 (5.92) |
| Platelets [#/volume] in blood (mean (SD)) | 262.74 (81.01) | 256.01 (79.33) |
| Calcium [mass/volume] in serum or plasma (mean (SD)) | 9.14 (0.49) | 9.20 (0.48) |
| Glucose [mass/volume] in serum or plasma (mean (SD)) | 110.66 (23.03) | 119.07 (24.41) |
| Lymphocytes/100 leukocytes in blood (mean (SD)) | 17.69 (6.69) | 17.93 (8.66) |
| Alkaline phosphatase [enzymatic activity/volume] in serum or plasma (mean (SD)) | 98.81 (32.38) | 97.80 (29.82) |
| Protein [mass/volume] in serum or plasma (mean (SD)) | 65.92 (5.50) | 67.35 (5.31) |
| Alanine aminotransferase [enzymatic activity/volume] in serum or plasma (mean (SD)) | 21.45 (10.63) | 23.93 (11.22) |
| Albumin [mass/volume] in serum or plasma (mean (SD)) | 36.95 (4.13) | 37.56 (4.19) |
| Bilirubin.total [mass/volume] in serum or plasma (mean (SD)) | 0.46 (0.18) | 0.39 (0.17) |
| Chloride [moles/volume] in serum or plasma (mean (SD)) | 101.69 (3.34) | 101.00 (2.93) |
| Monocytes [#/volume] in blood (mean (SD)) | 0.61 (0.19) | 0.56 (0.22) |
| Eosinophils/100 leukocytes in blood (mean (SD)) | 2.25 (1.19) | 1.82 (1.05) |
| Lactate dehydrogenase [enzymatic activity/volume] in serum or plasma (mean (SD)) | 206.34 (51.79) | 206.07 (49.36) |
| Heart rate (mean (SD)) | 82.48 (13.53) | 85.31 (13.52) |
| Systolic blood pressure (mean (SD)) | 125.40 (14.75) | 125.79 (14.79) |
| Oxygen saturation in arterial blood by pulse oximetry (mean (SD)) | 97.00 (0.93) | 96.97 (0.88) |
| ECOG (mean (SD)) | 0.96 (0.63) | 0.81 (0.58) |
| NLR (mean (SD)) | 3.79 (1.41) | 3.71 (1.36) |
| BMI (mean (SD)) | 25.50 (4.94) | 26.45 (5.28) |
| Tumor stage at baseline (mean (SD)) | 3.67 (0.76) | 3.53 (0.84) |
| AST/ALT ratio [%] (mean (SD)) | 1.17 (0.42) | 1.03 (0.38) |
| Histology = Squamous cell carcinoma (%) | 9 (3.6) | 18 (7.1) |

Supplementary Table 11: Baseline values for the emulated NCT00540514 clinical trial.

|  | Carboplatin + nab-Paclitaxel | Carboplatin + Paclitaxel |
| --- | --- | --- |
| Number of Patients | 1185 | 1185 |
| ROPRO (mean (SD)) | 0.21 (0.35) | 0.21 (0.35) |
| Age at baseline [years] (mean (SD)) | 69.54 (8.13) | 68.61 (8.55) |
| Gender = Male (mean (SD)) | 0.63 (0.48) | 0.59 (0.49) |
| History of Smoking = Yes (mean (SD)) | 0.95 (0.21) | 0.94 (0.25) |
| Number of metastatic sites (mean (SD)) | 0.20 (0.52) | 0.16 (0.46) |
| Hemoglobin [mass/volume] in blood (mean (SD)) | 11.25 (1.55) | 11.56 (1.60) |
| Urea nitrogen [mass/volume] in serum or plasma (mean (SD)) | 16.03 (5.95) | 16.46 (5.83) |
| Platelets [#/volume] in blood (mean (SD)) | 235.10 (74.87) | 243.77 (74.47) |
| Calcium [mass/volume] in serum or plasma (mean (SD)) | 9.10 (0.47) | 9.16 (0.46) |
| Glucose [mass/volume] in serum or plasma (mean (SD)) | 115.75 (24.16) | 116.57 (25.56) |
| Lymphocytes/100 leukocytes in blood (mean (SD)) | 20.72 (8.77) | 16.94 (8.08) |
| Alkaline phosphatase [enzymatic activity/volume] in serum or plasma (mean (SD)) | 91.24 (29.83) | 91.08 (29.08) |
| Protein [mass/volume] in serum or plasma (mean (SD)) | 66.10 (5.99) | 66.73 (5.89) |
| Alanine aminotransferase [enzymatic activity/volume] in serum or plasma (mean (SD)) | 20.48 (10.25) | 20.90 (10.13) |
| Albumin [mass/volume] in serum or plasma (mean (SD)) | 36.07 (3.94) | 36.47 (4.14) |
| Bilirubin.total [mass/volume] in serum or plasma (mean (SD)) | 0.43 (0.17) | 0.44 (0.18) |
| Chloride [moles/volume] in serum or plasma (mean (SD)) | 100.82 (2.96) | 100.85 (3.14) |
| Monocytes [#/volume] in blood (mean (SD)) | 0.49 (0.19) | 0.49 (0.21) |
| Eosinophils/100 leukocytes in blood (mean (SD)) | 1.89 (0.92) | 1.76 (0.91) |
| Lactate dehydrogenase [enzymatic activity/volume] in serum or plasma (mean (SD)) | 205.11 (53.73) | 203.01 (48.79) |
| Heart rate (mean (SD)) | 86.36 (13.51) | 87.48 (13.93) |
| Systolic blood pressure (mean (SD)) | 121.97 (14.67) | 122.00 (14.54) |
| Oxygen saturation in arterial blood by pulse oximetry (mean (SD)) | 96.82 (1.06) | 96.88 (1.00) |
| ECOG (mean (SD)) | 0.89 (0.58) | 0.96 (0.54) |
| NLR (mean (SD)) | 3.66 (1.42) | 3.86 (1.56) |
| BMI (mean (SD)) | 26.34 (5.45) | 25.84 (5.50) |
| Tumor stage at baseline (mean (SD)) | 3.49 (0.88) | 3.36 (0.91) |
| AST/ALT ratio [%] (mean (SD)) | 1.13 (0.38) | 1.14 (0.39) |
| Histology = Squamous cell carcinoma (%) | 915 (77.2) | 606 (51.1) |

Supplementary Table 12: Baseline values for the emulated AURA3 clinical trial.

|  | Osimertinib | Platinum + Pemetrexed |
| --- | --- | --- |
| Number of Patients | 475 | 475 |
| ROPRO (mean (SD)) | 0.12 (0.39) | 0.12 (0.39) |
| Age at baseline [years] (mean (SD)) | 71.99 (9.85) | 65.99 (9.03) |
| Gender = Male (mean (SD)) | 0.35 (0.48) | 0.49 (0.50) |
| History of Smoking = Yes (mean (SD)) | 0.52 (0.50) | 0.90 (0.30) |
| Number of metastatic sites (mean (SD)) | 0.73 (0.94) | 0.22 (0.57) |
| Hemoglobin [mass/volume] in blood (mean (SD)) | 12.11 (1.61) | 11.83 (1.62) |
| Urea nitrogen [mass/volume] in serum or plasma (mean (SD)) | 17.03 (6.08) | 16.47 (5.82) |
| Platelets [#/volume] in blood (mean (SD)) | 218.38 (68.97) | 241.06 (81.72) |
| Calcium [mass/volume] in serum or plasma (mean (SD)) | 9.09 (0.51) | 9.19 (0.46) |
| Glucose [mass/volume] in serum or plasma (mean (SD)) | 113.58 (22.88) | 116.41 (24.43) |
| Lymphocytes/100 leukocytes in blood (mean (SD)) | 18.25 (7.54) | 17.89 (8.79) |
| Alkaline phosphatase [enzymatic activity/volume] in serum or plasma (mean (SD)) | 94.40 (32.77) | 98.25 (31.60) |
| Protein [mass/volume] in serum or plasma (mean (SD)) | 66.42 (6.15) | 66.93 (5.62) |
| Alanine aminotransferase [enzymatic activity/volume] in serum or plasma (mean (SD)) | 18.82 (9.77) | 24.17 (11.34) |
| Albumin [mass/volume] in serum or plasma (mean (SD)) | 36.95 (3.94) | 37.10 (4.38) |
| Bilirubin.total [mass/volume] in serum or plasma (mean (SD)) | 0.45 (0.19) | 0.41 (0.18) |
| Chloride [moles/volume] in serum or plasma (mean (SD)) | 101.27 (3.19) | 100.79 (2.98) |
| Monocytes [#/volume] in blood (mean (SD)) | 0.56 (0.18) | 0.59 (0.22) |
| Eosinophils/100 leukocytes in blood (mean (SD)) | 2.23 (1.33) | 1.78 (0.98) |
| Lactate dehydrogenase [enzymatic activity/volume] in serum or plasma (mean (SD)) | 202.42 (38.34) | 205.75 (47.11) |
| Heart rate (mean (SD)) | 82.14 (13.44) | 86.48 (13.75) |
| Systolic blood pressure (mean (SD)) | 126.53 (16.47) | 124.49 (14.47) |
| Oxygen saturation in arterial blood by pulse oximetry (mean (SD)) | 96.81 (1.09) | 96.91 (0.95) |
| ECOG (mean (SD)) | 1.00 (0.60) | 0.87 (0.55) |
| NLR (mean (SD)) | 3.74 (1.42) | 3.72 (1.57) |
| BMI (mean (SD)) | 25.32 (5.29) | 26.16 (5.09) |
| Tumor stage at baseline (mean (SD)) | 3.68 (0.81) | 3.56 (0.85) |
| AST/ALT ratio [%] (mean (SD)) | 1.22 (0.42) | 1.06 (0.41) |
| Histology = Squamous cell carcinoma (%) | 11 (2.3) | 7 (1.5) |

Supplementary Table 13: Baseline values for the emulated NCT00520676 clinical trial.

|  | Carboplatin + Docetaxel | Carboplatin + Pemetrexed |
| --- | --- | --- |
| Number of Patients | 198 | 198 |
| ROPRO (mean (SD)) | 0.23 (0.33) | 0.23 (0.33) |
| Age at baseline [years] (mean (SD)) | 66.67 (8.45) | 68.08 (9.02) |
| Gender = Male (mean (SD)) | 0.54 (0.50) | 0.47 (0.50) |
| History of Smoking = Yes (mean (SD)) | 0.92 (0.26) | 0.91 (0.29) |
| Number of metastatic sites (mean (SD)) | 0.12 (0.45) | 0.25 (0.52) |
| Hemoglobin [mass/volume] in blood (mean (SD)) | 11.63 (1.66) | 11.59 (1.68) |
| Urea nitrogen [mass/volume] in serum or plasma (mean (SD)) | 16.66 (5.93) | 16.66 (6.18) |
| Platelets [#/volume] in blood (mean (SD)) | 247.58 (75.20) | 237.30 (86.23) |
| Calcium [mass/volume] in serum or plasma (mean (SD)) | 9.02 (0.48) | 9.07 (0.51) |
| Glucose [mass/volume] in serum or plasma (mean (SD)) | 117.94 (23.75) | 117.38 (23.88) |
| Lymphocytes/100 leukocytes in blood (mean (SD)) | 16.63 (6.99) | 17.01 (7.95) |
| Alkaline phosphatase [enzymatic activity/volume] in serum or plasma (mean (SD)) | 89.49 (30.10) | 101.17 (31.12) |
| Protein [mass/volume] in serum or plasma (mean (SD)) | 64.76 (5.58) | 65.91 (5.42) |
| Alanine aminotransferase [enzymatic activity/volume] in serum or plasma (mean (SD)) | 21.48 (10.99) | 24.64 (11.62) |
| Albumin [mass/volume] in serum or plasma (mean (SD)) | 35.81 (4.12) | 36.06 (4.14) |
| Bilirubin.total [mass/volume] in serum or plasma (mean (SD)) | 0.41 (0.18) | 0.42 (0.18) |
| Chloride [moles/volume] in serum or plasma (mean (SD)) | 100.54 (3.49) | 100.69 (3.14) |
| Monocytes [#/volume] in blood (mean (SD)) | 0.57 (0.22) | 0.58 (0.21) |
| Eosinophils/100 leukocytes in blood (mean (SD)) | 1.56 (0.92) | 1.72 (0.93) |
| Lactate dehydrogenase [enzymatic activity/volume] in serum or plasma (mean (SD)) | 200.56 (44.90) | 208.17 (45.96) |
| Heart rate (mean (SD)) | 89.44 (14.81) | 86.44 (13.27) |
| Systolic blood pressure (mean (SD)) | 121.47 (16.38) | 124.93 (15.05) |
| Oxygen saturation in arterial blood by pulse oximetry (mean (SD)) | 97.02 (1.13) | 96.85 (0.94) |
| ECOG (mean (SD)) | 0.90 (0.52) | 1.00 (0.55) |
| NLR (mean (SD)) | 3.65 (1.27) | 3.92 (1.55) |
| BMI (mean (SD)) | 25.64 (5.62) | 26.44 (5.19) |
| Tumor stage at baseline (mean (SD)) | 3.62 (0.81) | 3.55 (0.96) |
| AST/ALT ratio [%] (mean (SD)) | 1.05 (0.42) | 1.03 (0.36) |
| Histology = Non-squamous cell carcinoma (%) | 198 (100.0) | 198 (100.0) |

#### 1.2. Lower caliper population characteristics

Supplementary Table 14: Baseline values for the emulated KEYNOTE-189 clinical trial.

|  | Platinum + Pembrolizumab+Pemetrexed | Platinum + Pemetrexed |
| --- | --- | --- |
| Number of Patients | 472 | 472 |
| ROPRO (mean (SD)) | 0.18 (0.33) | 0.18 (0.33) |
| Age at baseline [years] (mean (SD)) | 67.89 (10.00) | 66.94 (9.62) |
| Gender = Male (mean (SD)) | 0.57 (0.50) | 0.49 (0.50) |
| History of Smoking = Yes (mean (SD)) | 0.88 (0.33) | 0.90 (0.29) |
| Number of metastatic sites (mean (SD)) | 0.69 (0.87) | 0.25 (0.58) |
| Hemoglobin [mass/volume] in blood (mean (SD)) | 11.83 (1.75) | 11.76 (1.70) |
| Urea nitrogen [mass/volume] in serum or plasma (mean (SD)) | 16.14 (5.82) | 16.28 (5.49) |
| Platelets [#/volume] in blood (mean (SD)) | 249.09 (85.40) | 240.58 (83.60) |
| Calcium [mass/volume] in serum or plasma (mean (SD)) | 9.18 (0.48) | 9.17 (0.49) |
| Glucose [mass/volume] in serum or plasma (mean (SD)) | 118.37 (24.76) | 118.98 (25.66) |
| Lymphocytes/100 leukocytes in blood (mean (SD)) | 17.89 (8.39) | 17.91 (8.59) |
| Alkaline phosphatase [enzymatic activity/volume] in serum or plasma (mean (SD)) | 102.01 (32.91) | 98.34 (29.90) |
| Protein [mass/volume] in serum or plasma (mean (SD)) | 67.23 (6.13) | 66.97 (5.87) |
| Alanine aminotransferase [enzymatic activity/volume] in serum or plasma (mean (SD)) | 23.90 (10.87) | 23.32 (10.94) |
| Albumin [mass/volume] in serum or plasma (mean (SD)) | 37.10 (4.22) | 36.84 (4.17) |
| Bilirubin.total [mass/volume] in serum or plasma (mean (SD)) | 0.42 (0.17) | 0.41 (0.18) |
| Chloride [moles/volume] in serum or plasma (mean (SD)) | 100.73 (3.27) | 100.66 (3.00) |
| Monocytes [#/volume] in blood (mean (SD)) | 0.59 (0.23) | 0.56 (0.22) |
| Eosinophils/100 leukocytes in blood (mean (SD)) | 1.68 (1.14) | 1.71 (0.94) |
| Lactate dehydrogenase [enzymatic activity/volume] in serum or plasma (mean (SD)) | 210.28 (52.68) | 210.50 (52.71) |
| Heart rate (mean (SD)) | 87.18 (13.68) | 86.94 (13.81) |
| Systolic blood pressure (mean (SD)) | 125.42 (14.93) | 124.46 (15.31) |
| Oxygen saturation in arterial blood by pulse oximetry (mean (SD)) | 96.83 (1.14) | 96.88 (1.02) |
| ECOG (mean (SD)) | 0.82 (0.59) | 0.94 (0.49) |
| NLR (mean (SD)) | 3.79 (1.64) | 3.87 (1.58) |
| BMI (mean (SD)) | 25.73 (5.14) | 25.79 (5.35) |
| Tumor stage at baseline (mean (SD)) | 3.73 (0.77) | 3.54 (0.90) |
| AST/ALT ratio [%] (mean (SD)) | 1.08 (0.38) | 1.10 (0.37) |
| Histology = Non-squamous cell carcinoma (%) | 472 (100.0) | 472 (100.0) |

Supplementary Table 15: Baseline values for the emulated KEYNOTE-024 clinical trial.

|  | Pembrolizumab | Platinum-doublet (5 types) |
| --- | --- | --- |
| Number of Patients | 969 | 969 |
| ROPRO (mean (SD)) | 0.18 (0.36) | 0.18 (0.36) |
| Age at baseline [years] (mean (SD)) | 71.67 (9.36) | 67.85 (9.35) |
| Gender = Male (mean (SD)) | 0.50 (0.50) | 0.58 (0.49) |
| History of Smoking = Yes (mean (SD)) | 0.93 (0.26) | 0.93 (0.24) |
| Number of metastatic sites (mean (SD)) | 0.55 (0.84) | 0.18 (0.50) |
| Hemoglobin [mass/volume] in blood (mean (SD)) | 12.18 (1.71) | 11.56 (1.65) |
| Urea nitrogen [mass/volume] in serum or plasma (mean (SD)) | 16.27 (5.88) | 16.36 (5.91) |
| Platelets [#/volume] in blood (mean (SD)) | 266.15 (81.60) | 243.39 (81.56) |
| Calcium [mass/volume] in serum or plasma (mean (SD)) | 9.23 (0.49) | 9.13 (0.47) |
| Glucose [mass/volume] in serum or plasma (mean (SD)) | 115.33 (24.24) | 116.37 (26.26) |
| Lymphocytes/100 leukocytes in blood (mean (SD)) | 17.68 (6.82) | 17.46 (8.30) |
| Alkaline phosphatase [enzymatic activity/volume] in serum or plasma (mean (SD)) | 95.17 (28.95) | 94.80 (31.12) |
| Protein [mass/volume] in serum or plasma (mean (SD)) | 67.86 (6.05) | 66.67 (5.90) |
| Alanine aminotransferase [enzymatic activity/volume] in serum or plasma (mean (SD)) | 19.17 (10.46) | 22.22 (10.78) |
| Albumin [mass/volume] in serum or plasma (mean (SD)) | 36.83 (4.27) | 36.58 (4.23) |
| Bilirubin.total [mass/volume] in serum or plasma (mean (SD)) | 0.45 (0.18) | 0.42 (0.18) |
| Chloride [moles/volume] in serum or plasma (mean (SD)) | 100.90 (3.32) | 100.90 (3.13) |
| Monocytes [#/volume] in blood (mean (SD)) | 0.63 (0.20) | 0.51 (0.22) |
| Eosinophils/100 leukocytes in blood (mean (SD)) | 2.28 (1.25) | 1.82 (1.02) |
| Lactate dehydrogenase [enzymatic activity/volume] in serum or plasma (mean (SD)) | 205.07 (49.05) | 207.34 (53.99) |
| Heart rate (mean (SD)) | 85.21 (14.12) | 87.10 (14.03) |
| Systolic blood pressure (mean (SD)) | 125.66 (16.17) | 123.41 (14.14) |
| Oxygen saturation in arterial blood by pulse oximetry (mean (SD)) | 96.78 (1.10) | 96.91 (0.95) |
| ECOG (mean (SD)) | 0.93 (0.62) | 0.94 (0.54) |
| NLR (mean (SD)) | 3.79 (1.42) | 3.83 (1.51) |
| BMI (mean (SD)) | 25.58 (5.45) | 26.26 (5.63) |
| Tumor stage at baseline (mean (SD)) | 3.43 (1.01) | 3.46 (0.87) |
| AST/ALT ratio [%] (mean (SD)) | 1.20 (0.42) | 1.13 (0.39) |
| Histology = Squamous cell carcinoma (%) | 272 (28.1) | 365 (37.7) |

Supplementary Table 16: Baseline values for the emulated KEYNOTE-042 clinical trial.

|  | Pembrolizumab | Platinum-doublet (2 types) |
| --- | --- | --- |
| Number of Patients | 860 | 860 |
| ROPRO (mean (SD)) | 0.18 (0.36) | 0.18 (0.36) |
| Age at baseline [years] (mean (SD)) | 71.52 (9.32) | 67.73 (9.44) |
| Gender = Male (mean (SD)) | 0.51 (0.50) | 0.57 (0.50) |
| History of Smoking = Yes (mean (SD)) | 0.93 (0.25) | 0.93 (0.25) |
| Number of metastatic sites (mean (SD)) | 0.54 (0.84) | 0.18 (0.49) |
| Hemoglobin [mass/volume] in blood (mean (SD)) | 12.17 (1.71) | 11.59 (1.64) |
| Urea nitrogen [mass/volume] in serum or plasma (mean (SD)) | 16.27 (5.90) | 16.22 (5.99) |
| Platelets [#/volume] in blood (mean (SD)) | 267.45 (82.01) | 245.74 (80.91) |
| Calcium [mass/volume] in serum or plasma (mean (SD)) | 9.23 (0.50) | 9.13 (0.47) |
| Glucose [mass/volume] in serum or plasma (mean (SD)) | 115.18 (24.23) | 116.66 (26.35) |
| Lymphocytes/100 leukocytes in blood (mean (SD)) | 17.73 (6.69) | 16.99 (8.00) |
| Alkaline phosphatase [enzymatic activity/volume] in serum or plasma (mean (SD)) | 95.80 (29.23) | 94.92 (31.44) |
| Protein [mass/volume] in serum or plasma (mean (SD)) | 67.96 (6.07) | 66.58 (5.93) |
| Alanine aminotransferase [enzymatic activity/volume] in serum or plasma (mean (SD)) | 19.27 (10.51) | 22.07 (10.69) |
| Albumin [mass/volume] in serum or plasma (mean (SD)) | 36.79 (4.28) | 36.62 (4.28) |
| Bilirubin.total [mass/volume] in serum or plasma (mean (SD)) | 0.45 (0.18) | 0.43 (0.18) |
| Chloride [moles/volume] in serum or plasma (mean (SD)) | 100.94 (3.31) | 100.94 (3.16) |
| Monocytes [#/volume] in blood (mean (SD)) | 0.63 (0.20) | 0.51 (0.22) |
| Eosinophils/100 leukocytes in blood (mean (SD)) | 2.27 (1.26) | 1.83 (1.02) |
| Lactate dehydrogenase [enzymatic activity/volume] in serum or plasma (mean (SD)) | 205.37 (49.51) | 208.40 (55.50) |
| Heart rate (mean (SD)) | 85.25 (14.27) | 87.47 (13.98) |
| Systolic blood pressure (mean (SD)) | 125.49 (15.98) | 123.19 (14.30) |
| Oxygen saturation in arterial blood by pulse oximetry (mean (SD)) | 96.77 (1.12) | 96.93 (0.96) |
| ECOG (mean (SD)) | 0.93 (0.62) | 0.94 (0.55) |
| NLR (mean (SD)) | 3.79 (1.44) | 3.83 (1.51) |
| BMI (mean (SD)) | 25.62 (5.46) | 26.20 (5.67) |
| Tumor stage at baseline (mean (SD)) | 3.42 (1.01) | 3.47 (0.86) |
| AST/ALT ratio [%] (mean (SD)) | 1.20 (0.42) | 1.13 (0.40) |
| Histology = Squamous cell carcinoma (%) | 244 (28.4) | 288 (33.5) |

Supplementary Table 17: Baseline values for the emulated KEYNOTE-407 clinical trial.

|  | Carboplatin + Paclitaxel | Carboplatin + Paclitaxel+Pembrolizumab |
| --- | --- | --- |
| Number of Patients | 171 | 171 |
| ROPRO (mean (SD)) | 0.18 (0.31) | 0.18 (0.31) |
| Age at baseline [years] (mean (SD)) | 69.70 (7.77) | 69.67 (8.12) |
| Gender = Male (mean (SD)) | 0.60 (0.49) | 0.68 (0.47) |
| History of Smoking = Yes (mean (SD)) | 0.99 (0.11) | 0.97 (0.17) |
| Number of metastatic sites (mean (SD)) | 0.12 (0.38) | 0.51 (0.80) |
| Hemoglobin [mass/volume] in blood (mean (SD)) | 11.48 (1.57) | 11.37 (1.61) |
| Urea nitrogen [mass/volume] in serum or plasma (mean (SD)) | 15.51 (5.61) | 16.02 (5.15) |
| Platelets [#/volume] in blood (mean (SD)) | 234.36 (73.77) | 236.82 (72.80) |
| Calcium [mass/volume] in serum or plasma (mean (SD)) | 9.15 (0.44) | 9.17 (0.47) |
| Glucose [mass/volume] in serum or plasma (mean (SD)) | 117.62 (25.93) | 117.30 (25.08) |
| Lymphocytes/100 leukocytes in blood (mean (SD)) | 18.33 (8.41) | 18.62 (7.65) |
| Alkaline phosphatase [enzymatic activity/volume] in serum or plasma (mean (SD)) | 86.46 (28.34) | 93.54 (28.78) |
| Protein [mass/volume] in serum or plasma (mean (SD)) | 66.91 (5.73) | 67.22 (5.78) |
| Alanine aminotransferase [enzymatic activity/volume] in serum or plasma (mean (SD)) | 19.85 (9.43) | 18.95 (9.60) |
| Albumin [mass/volume] in serum or plasma (mean (SD)) | 36.54 (3.73) | 37.02 (3.98) |
| Bilirubin.total [mass/volume] in serum or plasma (mean (SD)) | 0.44 (0.18) | 0.40 (0.17) |
| Chloride [moles/volume] in serum or plasma (mean (SD)) | 100.59 (3.15) | 101.32 (3.11) |
| Monocytes [#/volume] in blood (mean (SD)) | 0.45 (0.20) | 0.52 (0.20) |
| Eosinophils/100 leukocytes in blood (mean (SD)) | 1.90 (1.05) | 1.73 (0.92) |
| Lactate dehydrogenase [enzymatic activity/volume] in serum or plasma (mean (SD)) | 202.73 (46.72) | 203.94 (38.33) |
| Heart rate (mean (SD)) | 86.24 (13.93) | 86.17 (13.64) |
| Systolic blood pressure (mean (SD)) | 122.25 (13.43) | 123.71 (14.56) |
| Oxygen saturation in arterial blood by pulse oximetry (mean (SD)) | 96.75 (1.02) | 96.92 (1.16) |
| ECOG (mean (SD)) | 0.90 (0.56) | 0.82 (0.63) |
| NLR (mean (SD)) | 3.89 (1.60) | 3.82 (1.58) |
| BMI (mean (SD)) | 26.55 (5.54) | 26.03 (5.23) |
| Tumor stage at baseline (mean (SD)) | 3.35 (0.93) | 3.37 (1.03) |
| AST/ALT ratio [%] (mean (SD)) | 1.18 (0.33) | 1.18 (0.36) |
| Histology = Squamous cell carcinoma (%) | 171 (100.0) | 171 (100.0) |

Supplementary Table 18: Baseline values for the emulated PRONOUNCE clinical trial.

|  | Bevacizumab + Carboplatin+Paclitaxel | Carboplatin + Pemetrexed |
| --- | --- | --- |
| Number of Patients | 119 | 119 |
| ROPRO (mean (SD)) | 0.19 (0.33) | 0.19 (0.33) |
| Age at baseline [years] (mean (SD)) | 65.05 (9.62) | 66.66 (9.13) |
| Gender = Male (mean (SD)) | 0.53 (0.50) | 0.48 (0.50) |
| History of Smoking = Yes (mean (SD)) | 0.88 (0.32) | 0.89 (0.31) |
| Number of metastatic sites (mean (SD)) | 0.25 (0.74) | 0.18 (0.45) |
| Hemoglobin [mass/volume] in blood (mean (SD)) | 11.96 (1.73) | 11.69 (1.58) |
| Urea nitrogen [mass/volume] in serum or plasma (mean (SD)) | 16.20 (5.93) | 16.50 (5.88) |
| Platelets [#/volume] in blood (mean (SD)) | 245.16 (72.56) | 235.62 (86.89) |
| Calcium [mass/volume] in serum or plasma (mean (SD)) | 9.02 (0.45) | 9.17 (0.48) |
| Glucose [mass/volume] in serum or plasma (mean (SD)) | 118.17 (25.27) | 116.92 (25.79) |
| Lymphocytes/100 leukocytes in blood (mean (SD)) | 17.36 (8.16) | 17.02 (7.53) |
| Alkaline phosphatase [enzymatic activity/volume] in serum or plasma (mean (SD)) | 99.37 (33.22) | 98.49 (34.13) |
| Protein [mass/volume] in serum or plasma (mean (SD)) | 66.11 (5.74) | 66.37 (5.61) |
| Alanine aminotransferase [enzymatic activity/volume] in serum or plasma (mean (SD)) | 23.02 (11.07) | 26.19 (11.41) |
| Albumin [mass/volume] in serum or plasma (mean (SD)) | 36.07 (4.17) | 36.83 (4.18) |
| Bilirubin.total [mass/volume] in serum or plasma (mean (SD)) | 0.43 (0.19) | 0.43 (0.16) |
| Chloride [moles/volume] in serum or plasma (mean (SD)) | 100.85 (3.20) | 100.81 (2.64) |
| Monocytes [#/volume] in blood (mean (SD)) | 0.56 (0.24) | 0.59 (0.22) |
| Eosinophils/100 leukocytes in blood (mean (SD)) | 1.69 (0.86) | 1.55 (1.05) |
| Lactate dehydrogenase [enzymatic activity/volume] in serum or plasma (mean (SD)) | 213.11 (53.15) | 210.53 (46.11) |
| Heart rate (mean (SD)) | 87.31 (12.28) | 87.70 (14.09) |
| Systolic blood pressure (mean (SD)) | 125.46 (13.25) | 126.58 (14.46) |
| Oxygen saturation in arterial blood by pulse oximetry (mean (SD)) | 96.86 (1.05) | 96.90 (0.88) |
| ECOG (mean (SD)) | 0.95 (0.44) | 0.96 (0.53) |
| NLR (mean (SD)) | 3.81 (1.57) | 4.01 (1.67) |
| BMI (mean (SD)) | 25.36 (5.32) | 26.71 (5.60) |
| Tumor stage at baseline (mean (SD)) | 3.80 (0.57) | 3.57 (0.85) |
| AST/ALT ratio [%] (mean (SD)) | 1.09 (0.38) | 1.08 (0.39) |
| Histology = Non-squamous cell carcinoma (%) | 119 (100.0) | 119 (100.0) |

Supplementary Table 19: Baseline values for the emulated PointBreak clinical trial.

|  | Bevacizumab + Carboplatin+Paclitaxel | Bevacizumab + Carboplatin+Pemetrexed |
| --- | --- | --- |
| Number of Patients | 60 | 60 |
| ROPRO (mean (SD)) | 0.14 (0.26) | 0.14 (0.26) |
| Age at baseline [years] (mean (SD)) | 64.90 (8.89) | 64.15 (8.66) |
| Gender = Male (mean (SD)) | 0.47 (0.50) | 0.55 (0.50) |
| History of Smoking = Yes (mean (SD)) | 0.89 (0.31) | 0.88 (0.32) |
| Number of metastatic sites (mean (SD)) | 0.08 (0.33) | 0.18 (0.70) |
| Hemoglobin [mass/volume] in blood (mean (SD)) | 12.20 (1.86) | 11.55 (1.76) |
| Urea nitrogen [mass/volume] in serum or plasma (mean (SD)) | 15.71 (5.52) | 16.06 (5.50) |
| Platelets [#/volume] in blood (mean (SD)) | 231.43 (78.45) | 211.76 (90.00) |
| Calcium [mass/volume] in serum or plasma (mean (SD)) | 9.07 (0.49) | 9.20 (0.56) |
| Glucose [mass/volume] in serum or plasma (mean (SD)) | 114.68 (25.58) | 120.18 (27.09) |
| Lymphocytes/100 leukocytes in blood (mean (SD)) | 16.67 (6.94) | 18.74 (8.27) |
| Alkaline phosphatase [enzymatic activity/volume] in serum or plasma (mean (SD)) | 100.13 (24.22) | 104.83 (31.66) |
| Protein [mass/volume] in serum or plasma (mean (SD)) | 66.01 (5.67) | 66.95 (6.03) |
| Alanine aminotransferase [enzymatic activity/volume] in serum or plasma (mean (SD)) | 21.63 (10.37) | 24.60 (10.70) |
| Albumin [mass/volume] in serum or plasma (mean (SD)) | 36.42 (4.24) | 37.34 (4.36) |
| Bilirubin.total [mass/volume] in serum or plasma (mean (SD)) | 0.42 (0.16) | 0.41 (0.18) |
| Chloride [moles/volume] in serum or plasma (mean (SD)) | 101.29 (2.93) | 100.52 (2.90) |
| Monocytes [#/volume] in blood (mean (SD)) | 0.52 (0.25) | 0.56 (0.21) |
| Eosinophils/100 leukocytes in blood (mean (SD)) | 1.67 (0.86) | 1.91 (1.01) |
| Lactate dehydrogenase [enzymatic activity/volume] in serum or plasma (mean (SD)) | 208.11 (52.72) | 203.48 (45.29) |
| Heart rate (mean (SD)) | 85.58 (13.25) | 86.12 (15.04) |
| Systolic blood pressure (mean (SD)) | 125.72 (14.14) | 125.53 (16.35) |
| Oxygen saturation in arterial blood by pulse oximetry (mean (SD)) | 96.87 (0.92) | 96.88 (0.93) |
| ECOG (mean (SD)) | 0.98 (0.57) | 0.94 (0.45) |
| NLR (mean (SD)) | 4.11 (1.84) | 3.87 (1.46) |
| BMI (mean (SD)) | 24.62 (5.37) | 25.50 (4.65) |
| Tumor stage at baseline (mean (SD)) | 3.65 (0.77) | 3.58 (0.86) |
| AST/ALT ratio [%] (mean (SD)) | 1.15 (0.33) | 1.06 (0.34) |
| Histology = Non-squamous cell carcinoma (%) | 60 (100.0) | 60 (100.0) |

Supplementary Table 20: Baseline values for the emulated PROFILE 1014 clinical trial.

|  | Carboplatin-Cisplatin + Pemetrexed | Crizotinib |
| --- | --- | --- |
| Number of Patients | 61 | 61 |
| ROPRO (mean (SD)) | 0.19 (0.33) | 0.19 (0.33) |
| Age at baseline [years] (mean (SD)) | 66.36 (9.49) | 65.13 (11.66) |
| Gender = Male (mean (SD)) | 0.51 (0.50) | 0.52 (0.50) |
| History of Smoking = Yes (mean (SD)) | 0.97 (0.18) | 0.61 (0.49) |
| Number of metastatic sites (mean (SD)) | 0.15 (0.40) | 0.39 (0.74) |
| Hemoglobin [mass/volume] in blood (mean (SD)) | 11.84 (1.59) | 11.77 (2.10) |
| Urea nitrogen [mass/volume] in serum or plasma (mean (SD)) | 16.25 (6.08) | 16.72 (6.11) |
| Platelets [#/volume] in blood (mean (SD)) | 257.50 (73.37) | 262.62 (92.14) |
| Calcium [mass/volume] in serum or plasma (mean (SD)) | 9.23 (0.52) | 8.80 (0.50) |
| Glucose [mass/volume] in serum or plasma (mean (SD)) | 112.44 (20.27) | 114.10 (23.13) |
| Lymphocytes/100 leukocytes in blood (mean (SD)) | 16.83 (7.64) | 17.18 (7.31) |
| Alkaline phosphatase [enzymatic activity/volume] in serum or plasma (mean (SD)) | 96.95 (26.55) | 102.38 (26.61) |
| Protein [mass/volume] in serum or plasma (mean (SD)) | 66.59 (5.78) | 63.90 (6.33) |
| Alanine aminotransferase [enzymatic activity/volume] in serum or plasma (mean (SD)) | 22.81 (9.88) | 30.23 (12.35) |
| Albumin [mass/volume] in serum or plasma (mean (SD)) | 36.20 (4.31) | 33.71 (4.56) |
| Bilirubin.total [mass/volume] in serum or plasma (mean (SD)) | 0.40 (0.19) | 0.39 (0.19) |
| Chloride [moles/volume] in serum or plasma (mean (SD)) | 100.54 (3.03) | 101.78 (3.29) |
| Monocytes [#/volume] in blood (mean (SD)) | 0.57 (0.23) | 0.66 (0.26) |
| Eosinophils/100 leukocytes in blood (mean (SD)) | 1.68 (0.98) | 2.22 (1.30) |
| Lactate dehydrogenase [enzymatic activity/volume] in serum or plasma (mean (SD)) | 202.28 (38.98) | 209.86 (56.61) |
| Heart rate (mean (SD)) | 84.50 (11.75) | 80.91 (12.56) |
| Systolic blood pressure (mean (SD)) | 124.60 (14.72) | 122.48 (15.71) |
| Oxygen saturation in arterial blood by pulse oximetry (mean (SD)) | 97.01 (0.82) | 96.83 (0.93) |
| ECOG (mean (SD)) | 0.95 (0.54) | 1.04 (0.53) |
| NLR (mean (SD)) | 3.90 (1.53) | 3.94 (1.56) |
| BMI (mean (SD)) | 25.12 (5.74) | 26.08 (5.28) |
| Tumor stage at baseline (mean (SD)) | 3.50 (0.88) | 3.87 (0.47) |
| AST/ALT ratio [%] (mean (SD)) | 1.10 (0.40) | 1.04 (0.31) |
| Histology = Non-squamous cell carcinoma (%) | 61 (100.0) | 61 (100.0) |

Supplementary Table 21: Baseline values for the emulated FLAURA clinical trial.

|  | Erlotinib-Gefitinib | Osimertinib |
| --- | --- | --- |
| Number of Patients | 69 | 69 |
| ROPRO (mean (SD)) | -0.02 (0.37) | -0.02 (0.37) |
| Age at baseline [years] (mean (SD)) | 69.99 (9.42) | 69.54 (9.38) |
| Gender = Male (mean (SD)) | 0.32 (0.47) | 0.26 (0.44) |
| History of Smoking = Yes (mean (SD)) | 0.50 (0.50) | 0.49 (0.50) |
| Number of metastatic sites (mean (SD)) | 0.23 (0.55) | 0.61 (0.89) |
| Hemoglobin [mass/volume] in blood (mean (SD)) | 12.81 (1.42) | 12.21 (1.65) |
| Urea nitrogen [mass/volume] in serum or plasma (mean (SD)) | 16.71 (5.63) | 16.58 (5.33) |
| Platelets [#/volume] in blood (mean (SD)) | 258.38 (73.18) | 231.04 (63.64) |
| Calcium [mass/volume] in serum or plasma (mean (SD)) | 9.22 (0.46) | 9.00 (0.49) |
| Glucose [mass/volume] in serum or plasma (mean (SD)) | 118.91 (25.01) | 113.39 (21.84) |
| Lymphocytes/100 leukocytes in blood (mean (SD)) | 17.68 (7.01) | 17.95 (6.64) |
| Alkaline phosphatase [enzymatic activity/volume] in serum or plasma (mean (SD)) | 93.55 (29.60) | 96.82 (32.95) |
| Protein [mass/volume] in serum or plasma (mean (SD)) | 66.43 (6.14) | 66.61 (6.04) |
| Alanine aminotransferase [enzymatic activity/volume] in serum or plasma (mean (SD)) | 21.93 (9.30) | 21.33 (10.11) |
| Albumin [mass/volume] in serum or plasma (mean (SD)) | 37.50 (3.55) | 37.16 (4.30) |
| Bilirubin.total [mass/volume] in serum or plasma (mean (SD)) | 0.57 (0.21) | 0.44 (0.19) |
| Chloride [moles/volume] in serum or plasma (mean (SD)) | 101.92 (2.84) | 101.53 (2.89) |
| Monocytes [#/volume] in blood (mean (SD)) | 0.61 (0.16) | 0.55 (0.16) |
| Eosinophils/100 leukocytes in blood (mean (SD)) | 2.37 (1.43) | 2.08 (1.20) |
| Lactate dehydrogenase [enzymatic activity/volume] in serum or plasma (mean (SD)) | 203.98 (47.11) | 196.64 (19.10) |
| Heart rate (mean (SD)) | 85.17 (13.09) | 81.52 (12.17) |
| Systolic blood pressure (mean (SD)) | 128.54 (15.95) | 126.43 (18.52) |
| Oxygen saturation in arterial blood by pulse oximetry (mean (SD)) | 96.94 (0.93) | 96.86 (1.10) |
| ECOG (mean (SD)) | 0.91 (0.47) | 0.82 (0.66) |
| NLR (mean (SD)) | 3.80 (1.20) | 3.72 (1.33) |
| BMI (mean (SD)) | 25.80 (5.13) | 25.40 (4.64) |
| Tumor stage at baseline (mean (SD)) | 3.44 (0.93) | 3.72 (0.70) |
| AST/ALT ratio [%] (mean (SD)) | 1.10 (0.36) | 1.10 (0.35) |
| Histology = Squamous cell carcinoma (%) | 4 (5.8) | 2 (2.9) |

Supplementary Table 22: Baseline values for the emulated LUX-Lung 3+6 clinical trial.

|  | Afatinib | Platinum-doublet (3 types) |
| --- | --- | --- |
| Number of Patients | 75 | 75 |
| ROPRO (mean (SD)) | 0.07 (0.34) | 0.07 (0.34) |
| Age at baseline [years] (mean (SD)) | 70.57 (10.15) | 66.76 (9.63) |
| Gender = Male (mean (SD)) | 0.35 (0.48) | 0.57 (0.50) |
| History of Smoking = Yes (mean (SD)) | 0.54 (0.50) | 0.95 (0.23) |
| Number of metastatic sites (mean (SD)) | 0.45 (0.84) | 0.17 (0.45) |
| Hemoglobin [mass/volume] in blood (mean (SD)) | 12.18 (1.33) | 12.00 (1.58) |
| Urea nitrogen [mass/volume] in serum or plasma (mean (SD)) | 17.23 (6.37) | 16.74 (6.48) |
| Platelets [#/volume] in blood (mean (SD)) | 247.50 (85.88) | 264.81 (82.84) |
| Calcium [mass/volume] in serum or plasma (mean (SD)) | 9.13 (0.53) | 9.25 (0.53) |
| Glucose [mass/volume] in serum or plasma (mean (SD)) | 113.17 (25.41) | 117.06 (26.36) |
| Lymphocytes/100 leukocytes in blood (mean (SD)) | 17.92 (7.07) | 17.36 (8.53) |
| Alkaline phosphatase [enzymatic activity/volume] in serum or plasma (mean (SD)) | 100.25 (32.91) | 99.91 (32.87) |
| Protein [mass/volume] in serum or plasma (mean (SD)) | 66.23 (5.40) | 67.25 (5.63) |
| Alanine aminotransferase [enzymatic activity/volume] in serum or plasma (mean (SD)) | 20.29 (10.00) | 24.27 (11.11) |
| Albumin [mass/volume] in serum or plasma (mean (SD)) | 37.61 (4.34) | 37.57 (4.17) |
| Bilirubin.total [mass/volume] in serum or plasma (mean (SD)) | 0.44 (0.18) | 0.43 (0.19) |
| Chloride [moles/volume] in serum or plasma (mean (SD)) | 101.23 (3.53) | 101.01 (2.35) |
| Monocytes [#/volume] in blood (mean (SD)) | 0.60 (0.20) | 0.56 (0.22) |
| Eosinophils/100 leukocytes in blood (mean (SD)) | 2.17 (1.18) | 1.95 (1.17) |
| Lactate dehydrogenase [enzymatic activity/volume] in serum or plasma (mean (SD)) | 206.71 (52.40) | 202.71 (37.87) |
| Heart rate (mean (SD)) | 81.10 (12.95) | 86.40 (13.32) |
| Systolic blood pressure (mean (SD)) | 126.77 (15.59) | 123.69 (13.63) |
| Oxygen saturation in arterial blood by pulse oximetry (mean (SD)) | 97.09 (0.91) | 97.01 (0.86) |
| ECOG (mean (SD)) | 1.03 (0.64) | 0.77 (0.58) |
| NLR (mean (SD)) | 4.01 (1.73) | 3.84 (1.50) |
| BMI (mean (SD)) | 24.67 (5.13) | 25.87 (5.09) |
| Tumor stage at baseline (mean (SD)) | 3.77 (0.63) | 3.48 (0.90) |
| AST/ALT ratio [%] (mean (SD)) | 1.16 (0.46) | 0.95 (0.35) |
| Histology = Squamous cell carcinoma (%) | 1 (1.3) | 8 (10.7) |

Supplementary Table 23: Baseline values for the emulated NCT00540514 clinical trial.

|  | Carboplatin + nab-Paclitaxel | Carboplatin + Paclitaxel |
| --- | --- | --- |
| Number of Patients | 395 | 395 |
| ROPRO (mean (SD)) | 0.21 (0.31) | 0.21 (0.31) |
| Age at baseline [years] (mean (SD)) | 69.79 (8.03) | 68.61 (8.42) |
| Gender = Male (mean (SD)) | 0.61 (0.49) | 0.59 (0.49) |
| History of Smoking = Yes (mean (SD)) | 0.94 (0.23) | 0.95 (0.21) |
| Number of metastatic sites (mean (SD)) | 0.19 (0.51) | 0.18 (0.49) |
| Hemoglobin [mass/volume] in blood (mean (SD)) | 11.23 (1.63) | 11.62 (1.67) |
| Urea nitrogen [mass/volume] in serum or plasma (mean (SD)) | 15.74 (5.95) | 16.53 (5.86) |
| Platelets [#/volume] in blood (mean (SD)) | 235.50 (72.83) | 249.06 (76.08) |
| Calcium [mass/volume] in serum or plasma (mean (SD)) | 9.08 (0.47) | 9.16 (0.49) |
| Glucose [mass/volume] in serum or plasma (mean (SD)) | 116.27 (23.98) | 116.49 (25.36) |
| Lymphocytes/100 leukocytes in blood (mean (SD)) | 20.13 (8.58) | 16.91 (7.91) |
| Alkaline phosphatase [enzymatic activity/volume] in serum or plasma (mean (SD)) | 92.67 (29.87) | 92.59 (30.47) |
| Protein [mass/volume] in serum or plasma (mean (SD)) | 66.12 (6.11) | 66.50 (6.07) |
| Alanine aminotransferase [enzymatic activity/volume] in serum or plasma (mean (SD)) | 20.33 (9.89) | 20.58 (9.94) |
| Albumin [mass/volume] in serum or plasma (mean (SD)) | 35.87 (3.86) | 36.60 (4.23) |
| Bilirubin.total [mass/volume] in serum or plasma (mean (SD)) | 0.42 (0.17) | 0.42 (0.16) |
| Chloride [moles/volume] in serum or plasma (mean (SD)) | 100.82 (2.96) | 100.97 (3.04) |
| Monocytes [#/volume] in blood (mean (SD)) | 0.49 (0.19) | 0.49 (0.22) |
| Eosinophils/100 leukocytes in blood (mean (SD)) | 1.93 (0.84) | 1.75 (0.83) |
| Lactate dehydrogenase [enzymatic activity/volume] in serum or plasma (mean (SD)) | 208.58 (60.14) | 202.18 (52.61) |
| Heart rate (mean (SD)) | 86.09 (13.15) | 87.84 (14.27) |
| Systolic blood pressure (mean (SD)) | 122.06 (14.50) | 122.83 (14.48) |
| Oxygen saturation in arterial blood by pulse oximetry (mean (SD)) | 96.86 (0.99) | 96.82 (0.95) |
| ECOG (mean (SD)) | 0.90 (0.54) | 0.98 (0.51) |
| NLR (mean (SD)) | 3.64 (1.38) | 3.93 (1.57) |
| BMI (mean (SD)) | 26.19 (5.68) | 25.68 (5.43) |
| Tumor stage at baseline (mean (SD)) | 3.47 (0.89) | 3.39 (0.84) |
| AST/ALT ratio [%] (mean (SD)) | 1.13 (0.39) | 1.16 (0.38) |
| Histology = Squamous cell carcinoma (%) | 300 (75.9) | 201 (50.9) |

Supplementary Table 24: Baseline values for the emulated AURA3 clinical trial.

|  | Osimertinib | Platinum + Pemetrexed |
| --- | --- | --- |
| Number of Patients | 113 | 113 |
| ROPRO (mean (SD)) | 0.10 (0.33) | 0.10 (0.33) |
| Age at baseline [years] (mean (SD)) | 71.78 (10.48) | 65.68 (8.83) |
| Gender = Male (mean (SD)) | 0.34 (0.47) | 0.44 (0.50) |
| History of Smoking = Yes (mean (SD)) | 0.50 (0.50) | 0.88 (0.32) |
| Number of metastatic sites (mean (SD)) | 0.80 (1.12) | 0.22 (0.55) |
| Hemoglobin [mass/volume] in blood (mean (SD)) | 12.15 (1.52) | 11.72 (1.77) |
| Urea nitrogen [mass/volume] in serum or plasma (mean (SD)) | 16.81 (5.79) | 16.48 (5.86) |
| Platelets [#/volume] in blood (mean (SD)) | 217.39 (66.17) | 235.77 (87.07) |
| Calcium [mass/volume] in serum or plasma (mean (SD)) | 9.11 (0.46) | 9.20 (0.38) |
| Glucose [mass/volume] in serum or plasma (mean (SD)) | 113.49 (23.67) | 114.74 (22.28) |
| Lymphocytes/100 leukocytes in blood (mean (SD)) | 18.12 (7.16) | 16.91 (7.93) |
| Alkaline phosphatase [enzymatic activity/volume] in serum or plasma (mean (SD)) | 95.12 (33.34) | 96.34 (29.28) |
| Protein [mass/volume] in serum or plasma (mean (SD)) | 67.20 (5.40) | 67.29 (5.17) |
| Alanine aminotransferase [enzymatic activity/volume] in serum or plasma (mean (SD)) | 19.73 (9.68) | 25.67 (12.24) |
| Albumin [mass/volume] in serum or plasma (mean (SD)) | 37.35 (3.88) | 37.50 (3.88) |
| Bilirubin.total [mass/volume] in serum or plasma (mean (SD)) | 0.49 (0.19) | 0.40 (0.16) |
| Chloride [moles/volume] in serum or plasma (mean (SD)) | 101.22 (3.36) | 100.93 (2.76) |
| Monocytes [#/volume] in blood (mean (SD)) | 0.57 (0.16) | 0.60 (0.22) |
| Eosinophils/100 leukocytes in blood (mean (SD)) | 2.30 (1.47) | 1.74 (0.87) |
| Lactate dehydrogenase [enzymatic activity/volume] in serum or plasma (mean (SD)) | 200.58 (39.14) | 211.33 (60.42) |
| Heart rate (mean (SD)) | 83.05 (13.41) | 86.74 (12.97) |
| Systolic blood pressure (mean (SD)) | 125.96 (16.51) | 123.88 (12.36) |
| Oxygen saturation in arterial blood by pulse oximetry (mean (SD)) | 96.92 (1.07) | 96.85 (0.91) |
| ECOG (mean (SD)) | 0.95 (0.60) | 0.91 (0.53) |
| NLR (mean (SD)) | 3.91 (1.48) | 3.73 (1.51) |
| BMI (mean (SD)) | 25.42 (5.22) | 26.03 (4.89) |
| Tumor stage at baseline (mean (SD)) | 3.78 (0.72) | 3.52 (0.90) |
| AST/ALT ratio [%] (mean (SD)) | 1.20 (0.39) | 1.04 (0.37) |
| Histology = Squamous cell carcinoma (%) | 1 (0.9) | 3 (2.7) |

Supplementary Table 25: Baseline values for the emulated NCT00520676 clinical trial.

|  | Carboplatin + Docetaxel | Carboplatin + Pemetrexed |
| --- | --- | --- |
| Number of Patients | 55 | 55 |
| ROPRO (mean (SD)) | 0.23 (0.29) | 0.23 (0.29) |
| Age at baseline [years] (mean (SD)) | 65.58 (7.76) | 67.93 (7.98) |
| Gender = Male (mean (SD)) | 0.58 (0.50) | 0.49 (0.50) |
| History of Smoking = Yes (mean (SD)) | 0.90 (0.30) | 0.89 (0.31) |
| Number of metastatic sites (mean (SD)) | 0.07 (0.26) | 0.24 (0.47) |
| Hemoglobin [mass/volume] in blood (mean (SD)) | 11.70 (1.61) | 11.66 (1.74) |
| Urea nitrogen [mass/volume] in serum or plasma (mean (SD)) | 16.29 (5.38) | 16.70 (5.48) |
| Platelets [#/volume] in blood (mean (SD)) | 248.03 (74.82) | 233.56 (74.37) |
| Calcium [mass/volume] in serum or plasma (mean (SD)) | 8.98 (0.42) | 9.04 (0.55) |
| Glucose [mass/volume] in serum or plasma (mean (SD)) | 116.36 (25.15) | 115.33 (22.71) |
| Lymphocytes/100 leukocytes in blood (mean (SD)) | 16.51 (7.95) | 16.43 (7.78) |
| Alkaline phosphatase [enzymatic activity/volume] in serum or plasma (mean (SD)) | 86.55 (23.71) | 96.70 (29.27) |
| Protein [mass/volume] in serum or plasma (mean (SD)) | 64.64 (5.39) | 65.75 (4.73) |
| Alanine aminotransferase [enzymatic activity/volume] in serum or plasma (mean (SD)) | 21.62 (10.36) | 23.70 (10.58) |
| Albumin [mass/volume] in serum or plasma (mean (SD)) | 35.05 (4.10) | 36.58 (3.58) |
| Bilirubin.total [mass/volume] in serum or plasma (mean (SD)) | 0.39 (0.18) | 0.45 (0.17) |
| Chloride [moles/volume] in serum or plasma (mean (SD)) | 100.37 (3.06) | 100.47 (3.27) |
| Monocytes [#/volume] in blood (mean (SD)) | 0.61 (0.23) | 0.62 (0.22) |
| Eosinophils/100 leukocytes in blood (mean (SD)) | 1.59 (1.06) | 1.48 (0.80) |
| Lactate dehydrogenase [enzymatic activity/volume] in serum or plasma (mean (SD)) | 204.82 (57.29) | 202.83 (34.72) |
| Heart rate (mean (SD)) | 89.55 (15.29) | 85.30 (12.36) |
| Systolic blood pressure (mean (SD)) | 122.18 (13.28) | 126.37 (14.97) |
| Oxygen saturation in arterial blood by pulse oximetry (mean (SD)) | 97.02 (0.86) | 96.98 (0.82) |
| ECOG (mean (SD)) | 0.87 (0.51) | 1.01 (0.62) |
| NLR (mean (SD)) | 3.40 (1.18) | 4.06 (1.54) |
| BMI (mean (SD)) | 25.46 (5.01) | 26.33 (5.59) |
| Tumor stage at baseline (mean (SD)) | 3.60 (0.82) | 3.55 (0.93) |
| AST/ALT ratio [%] (mean (SD)) | 0.99 (0.41) | 1.01 (0.29) |
| Histology = Non-squamous cell carcinoma (%) | 55 (100.0) | 55 (100.0) |

#### 1.3. Clinical Trial validation population characteristics

Supplementary Table 50: Baseline values for the IMpower150 clinical trial.

|  | ABCP | BCP |
| --- | --- | --- |
| Number of Patients | 387 | 389 |
| ROPRO (mean (SD)) | -0.06 (0.51) | -0.08 (0.49) |
| Age at baseline [years] (mean (SD)) | 62.95 (9.37) | 63.11 (9.29) |
| Gender = Male (mean (SD)) | 0.61 (0.49) | 0.59 (0.49) |
| History of Smoking = Yes (mean (SD)) | 0.80 (0.40) | 0.81 (0.40) |
| Number of metastatic sites (mean (SD)) | 1.41 (0.64) | 1.39 (0.69) |
| Hemoglobin [mass/volume] in blood (mean (SD)) | 13.02 (1.62) | 13.04 (1.67) |
| Urea nitrogen [mass/volume] in serum or plasma (mean (SD)) | 15.80 (5.00) | 16.19 (5.36) |
| Platelets [#/volume] in blood (mean (SD)) | 301.33 (105.32) | 296.90 (93.38) |
| Calcium [mass/volume] in serum or plasma (mean (SD)) | 9.40 (0.58) | 9.36 (0.55) |
| Glucose [mass/volume] in serum or plasma (mean (SD)) | 114.54 (34.18) | 113.43 (31.61) |
| Lymphocytes/100 leukocytes in blood (mean (SD)) | 23.80 (5.88) | 23.39 (5.82) |
| Alkaline phosphatase [enzymatic activity/volume] in serum or plasma (mean (SD)) | 136.81 (127.41) | 123.61 (79.09) |
| Protein [mass/volume] in serum or plasma (mean (SD)) | 70.45 (5.86) | 70.37 (6.28) |
| Alanine aminotransferase [enzymatic activity/volume] in serum or plasma (mean (SD)) | 22.36 (16.79) | 23.30 (14.07) |
| Albumin [mass/volume] in serum or plasma (mean (SD)) | 38.07 (5.54) | 38.39 (5.54) |
| Bilirubin.total [mass/volume] in serum or plasma (mean (SD)) | 0.49 (0.28) | 0.49 (0.24) |
| Chloride [moles/volume] in serum or plasma (mean (SD)) | 102.00 (3.99) | 101.78 (3.78) |
| Monocytes [#/volume] in blood (mean (SD)) | 0.62 (0.30) | 0.64 (0.27) |
| Eosinophils/100 leukocytes in blood (mean (SD)) | 2.28 (1.22) | 2.34 (1.40) |
| Lactate dehydrogenase [enzymatic activity/volume] in serum or plasma (mean (SD)) | 286.28 (174.30) | 288.31 (199.08) |
| Heart rate (mean (SD)) | 79.86 (11.99) | 81.25 (12.24) |
| Systolic blood pressure (mean (SD)) | 125.10 (14.60) | 128.25 (14.68) |
| Oxygen saturation in arterial blood by pulse oximetry (mean (SD)) | 96.49 (0.00) | 96.49 (0.00) |
| ECOG (mean (SD)) | 0.60 (0.50) | 0.59 (0.52) |
| NLR (mean (SD)) | 4.95 (5.28) | 4.34 (5.05) |
| BMI (mean (SD)) | 25.57 (5.53) | 25.22 (4.60) |
| Tumor stage at baseline (mean (SD)) | 3.67 (0.81) | 3.63 (0.84) |
| AST/ALT ratio [%] (mean (SD)) | 1.19 (0.62) | 1.14 (0.44) |

Supplementary Table 51: Baseline values for the IMvigor211 clinical trial.

|  | Atezolizumab | Chemo |
| --- | --- | --- |
| Number of Patients | 454 | 429 |
| ROPRO (mean (SD)) | -0.07 (0.53) | -0.05 (0.53) |
| Age at baseline [years] (mean (SD)) | 66.04 (9.43) | 66.12 (9.36) |
| Gender = Male (mean (SD)) | 0.76 (0.42) | 0.78 (0.42) |
| History of Smoking = Yes (mean (SD)) | 0.69 (0.46) | 0.73 (0.44) |
| Number of metastatic sites (mean (SD)) | 1.39 (0.76) | 1.38 (0.64) |
| Hemoglobin [mass/volume] in blood (mean (SD)) | 11.84 (1.70) | 11.76 (1.69) |
| Urea nitrogen [mass/volume] in serum or plasma (mean (SD)) | 20.04 (6.49) | 20.61 (7.32) |
| Platelets [#/volume] in blood (mean (SD)) | 259.31 (92.92) | 268.28 (87.20) |
| Calcium [mass/volume] in serum or plasma (mean (SD)) | 9.37 (0.54) | 9.34 (0.53) |
| Glucose [mass/volume] in serum or plasma (mean (SD)) | 111.05 (28.07) | 113.12 (31.47) |
| Lymphocytes/100 leukocytes in blood (mean (SD)) | 22.94 (4.83) | 22.90 (4.73) |
| Alkaline phosphatase [enzymatic activity/volume] in serum or plasma (mean (SD)) | 135.56 (108.15) | 128.67 (98.12) |
| Protein [mass/volume] in serum or plasma (mean (SD)) | 71.52 (6.30) | 71.13 (6.12) |
| Alanine aminotransferase [enzymatic activity/volume] in serum or plasma (mean (SD)) | 22.74 (21.35) | 21.04 (15.05) |
| Albumin [mass/volume] in serum or plasma (mean (SD)) | 38.34 (5.19) | 38.41 (5.33) |
| Bilirubin.total [mass/volume] in serum or plasma (mean (SD)) | 0.45 (0.30) | 0.42 (0.21) |
| Chloride [moles/volume] in serum or plasma (mean (SD)) | 101.51 (4.19) | 101.37 (4.36) |
| Monocytes [#/volume] in blood (mean (SD)) | 0.67 (0.26) | 0.66 (0.29) |
| Eosinophils/100 leukocytes in blood (mean (SD)) | 2.35 (1.14) | 2.29 (0.81) |
| Lactate dehydrogenase [enzymatic activity/volume] in serum or plasma (mean (SD)) | 297.41 (235.56) | 295.96 (264.15) |
| Heart rate (mean (SD)) | 74.97 (11.73) | 78.24 (12.13) |
| Systolic blood pressure (mean (SD)) | 127.78 (15.49) | 131.81 (15.61) |
| Oxygen saturation in arterial blood by pulse oximetry (mean (SD)) | 96.49 (0.00) | 96.49 (0.00) |
| ECOG (mean (SD)) | 0.56 (0.52) | 0.59 (0.51) |
| NLR (mean (SD)) | 4.18 (3.13) | 4.74 (3.99) |
| BMI (mean (SD)) | 26.07 (4.59) | 26.19 (4.38) |
| Tumor stage at baseline (mean (SD)) | 2.16 (1.02) | 2.20 (0.99) |
| AST/ALT ratio [%] (mean (SD)) | 1.29 (0.54) | 1.26 (0.56) |

Supplementary Table 52: Baseline values for the OAK clinical trial.

|  | Atezolizumab | Docetaxel |
| --- | --- | --- |
| Number of Patients | 598 | 564 |
| ROPRO (mean (SD)) | 0.04 (0.51) | 0.05 (0.46) |
| Age at baseline [years] (mean (SD)) | 62.78 (9.78) | 62.84 (9.27) |
| Gender = Male (mean (SD)) | 0.62 (0.49) | 0.62 (0.49) |
| History of Smoking = Yes (mean (SD)) | 0.82 (0.38) | 0.85 (0.36) |
| Number of metastatic sites (mean (SD)) | 1.31 (0.64) | 1.36 (0.65) |
| Hemoglobin [mass/volume] in blood (mean (SD)) | 12.31 (1.72) | 12.15 (1.56) |
| Urea nitrogen [mass/volume] in serum or plasma (mean (SD)) | 15.78 (4.79) | 16.34 (6.05) |
| Platelets [#/volume] in blood (mean (SD)) | 277.90 (96.26) | 285.39 (93.68) |
| Calcium [mass/volume] in serum or plasma (mean (SD)) | 9.38 (0.55) | 9.45 (0.59) |
| Glucose [mass/volume] in serum or plasma (mean (SD)) | 76.61 (32.91) | 84.79 (41.05) |
| Lymphocytes/100 leukocytes in blood (mean (SD)) | 20.09 (6.51) | 20.00 (6.93) |
| Alkaline phosphatase [enzymatic activity/volume] in serum or plasma (mean (SD)) | 121.37 (87.38) | 117.84 (74.20) |
| Protein [mass/volume] in serum or plasma (mean (SD)) | 67.30 (9.26) | 67.50 (9.19) |
| Alanine aminotransferase [enzymatic activity/volume] in serum or plasma (mean (SD)) | 21.45 (13.69) | 21.09 (14.09) |
| Albumin [mass/volume] in serum or plasma (mean (SD)) | 38.22 (5.43) | 39.03 (4.96) |
| Bilirubin.total [mass/volume] in serum or plasma (mean (SD)) | 0.49 (0.63) | 0.45 (0.26) |
| Chloride [moles/volume] in serum or plasma (mean (SD)) | 101.39 (3.94) | 101.27 (3.75) |
| Monocytes [#/volume] in blood (mean (SD)) | 0.64 (0.23) | 0.62 (0.26) |
| Eosinophils/100 leukocytes in blood (mean (SD)) | 2.33 (1.58) | 2.17 (1.78) |
| Lactate dehydrogenase [enzymatic activity/volume] in serum or plasma (mean (SD)) | 306.32 (234.13) | 303.48 (250.38) |
| Heart rate (mean (SD)) | 80.14 (12.87) | 82.04 (13.69) |
| Systolic blood pressure (mean (SD)) | 122.74 (15.47) | 125.35 (15.42) |
| Oxygen saturation in arterial blood by pulse oximetry (mean (SD)) | 96.49 (0.00) | 96.49 (0.00) |
| ECOG (mean (SD)) | 0.64 (0.49) | 0.64 (0.49) |
| NLR (mean (SD)) | 4.44 (3.22) | 6.27 (6.46) |
| BMI (mean (SD)) | 25.38 (5.04) | 25.00 (4.60) |
| Tumor stage at baseline (mean (SD)) | 3.41 (0.90) | 3.45 (0.90) |
| AST/ALT ratio [%] (mean (SD)) | 1.30 (0.58) | 1.29 (0.60) |
