## Supplementary Figures and Tables for "Longitudinal assessment of ROPRO as an early indicator of overall survival in oncology clinical trials: a retrospective analysis": Supplementary Figures.docx

### 1. Supplementary Figures

Supplementary Figure 1: Selection process of emulated clinical trials. The criteria used to exclude clinical trials and number of clinical trials excluded are represented in the rectangles to the right.

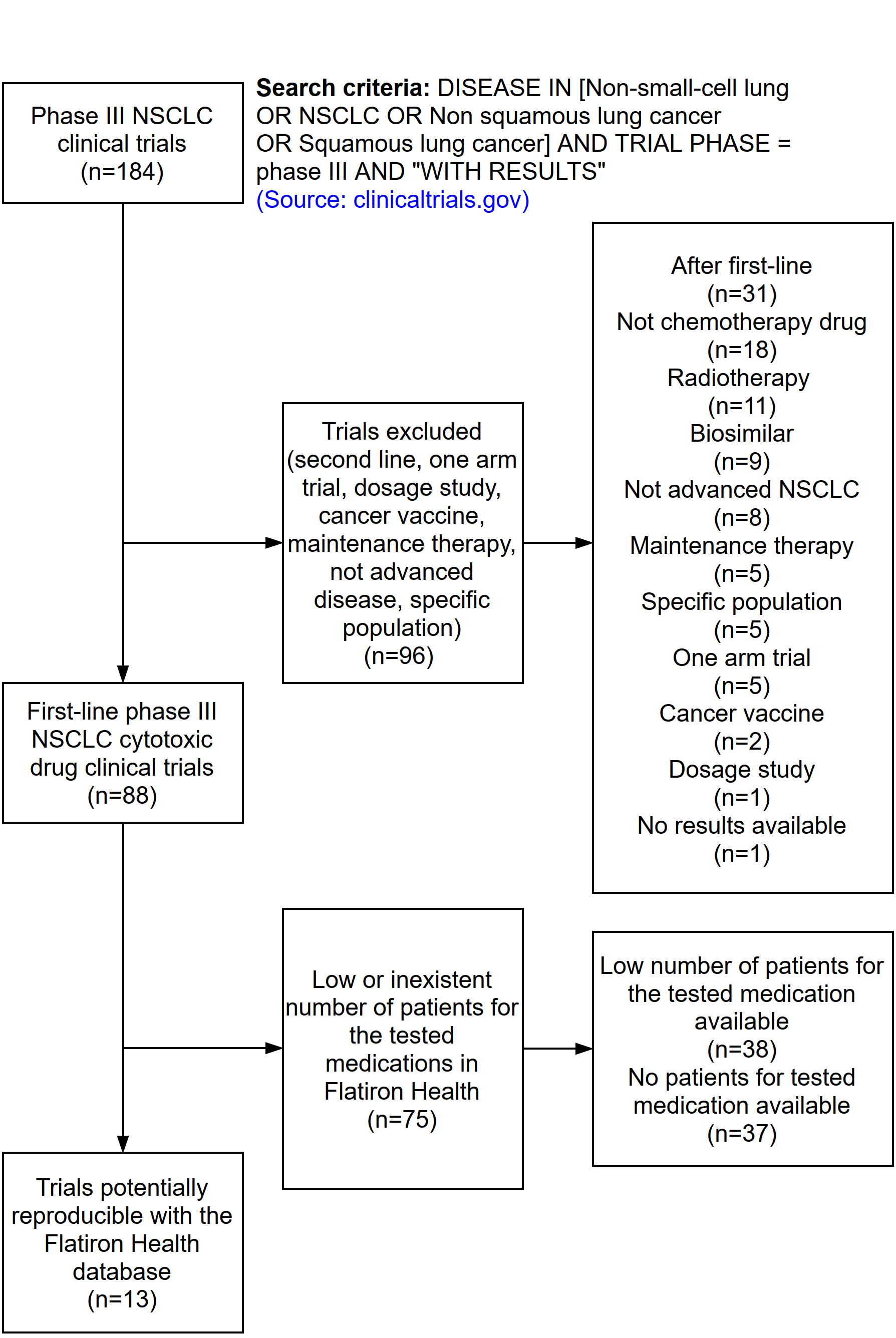

Supplementary Figure 2: Comparison of the Overall Survival (OS) Hazard Ratios (HR) of the original (in red) and emulated (in blue) clinical trials. The panel A and B represent, respectively, the higher caliper (more patients) and lower caliper (fewer patients) analyses.

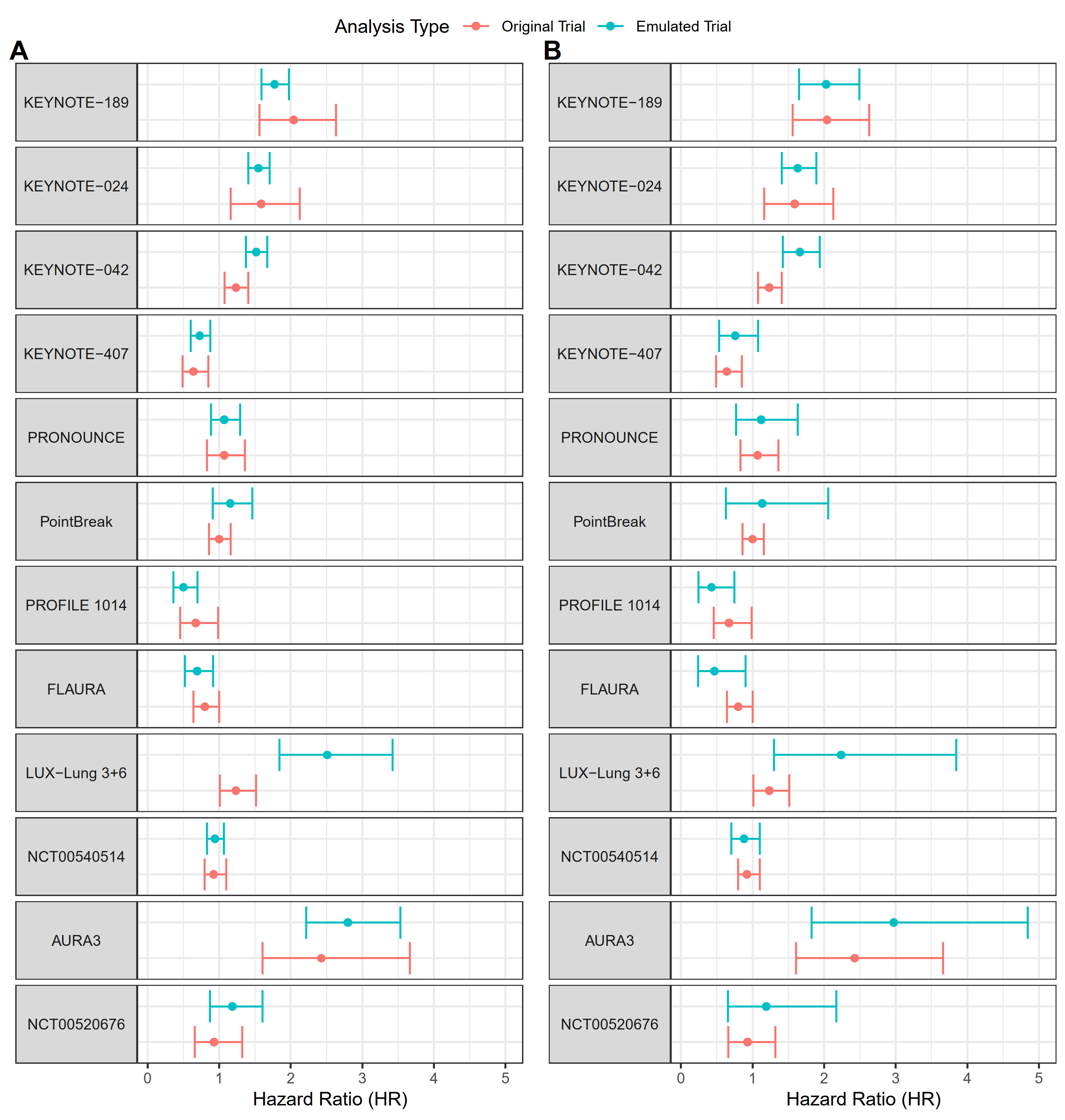

#### 1.1. ΔRisk and Overall Survival plots for each recreated clinical trial (with treatment arm in the hazard function)

This document contains the ΔRisk for ROPRO and DeepROPRO (on top) and Overall Survival (OS) Kaplan Meier (KM) (on the bottom) plots. In the ΔRisk and KM plots, there are black dots on the bottom of each figure. These dots indicate a significant difference between the treatment arms at that time-point. In the ΔROPRO and ΔDeepROPRO analyses, the p-value was derived from the likelihood-ratio test described in the supplementary materials. In the KM plots, the p-value was determined with the log-rank test. Additionally, the KM plots contain the treatment-effect hazard ratio (HR) and corresponding p-value. These values were obtained from an auxiliary Cox model.

In each figure, the hazard ratio (HR) refers to the effect of the second medication in the scale (that is represented in purple).

##### 1.1.1. Higher caliper ΔRisk and Overall Survival plots

Supplementary Figure 3: ΔRisk and Kaplan Meier plots for the KEYNOTE-189 clinical trial. The HR is referent to the treatment effect of Platinum+Pemetrexed.

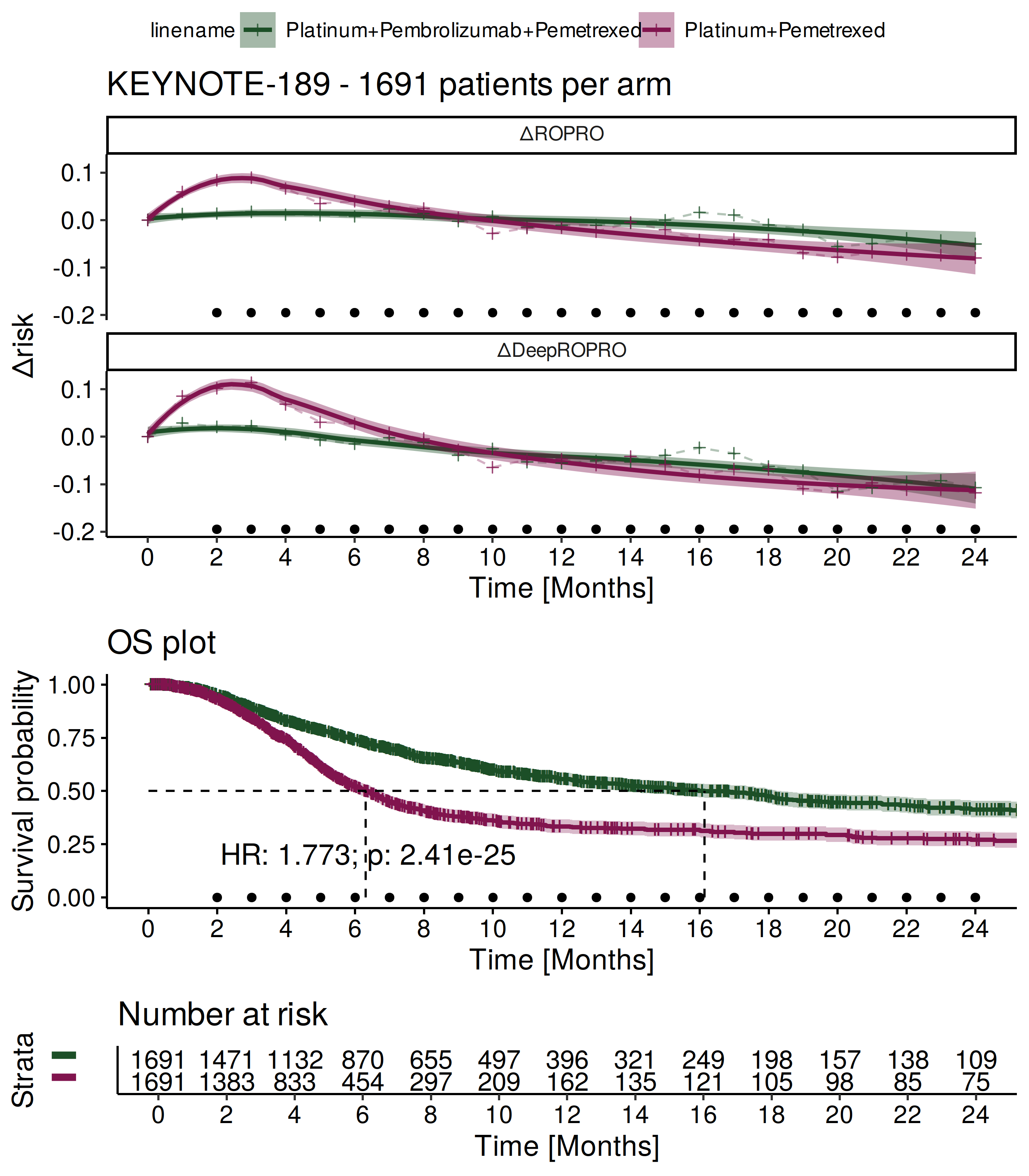

Supplementary Figure 4: ΔRisk and Kaplan Meier plots for the KEYNOTE-024 clinical trial. The HR is referent to the treatment effect of Pembrolizumab.

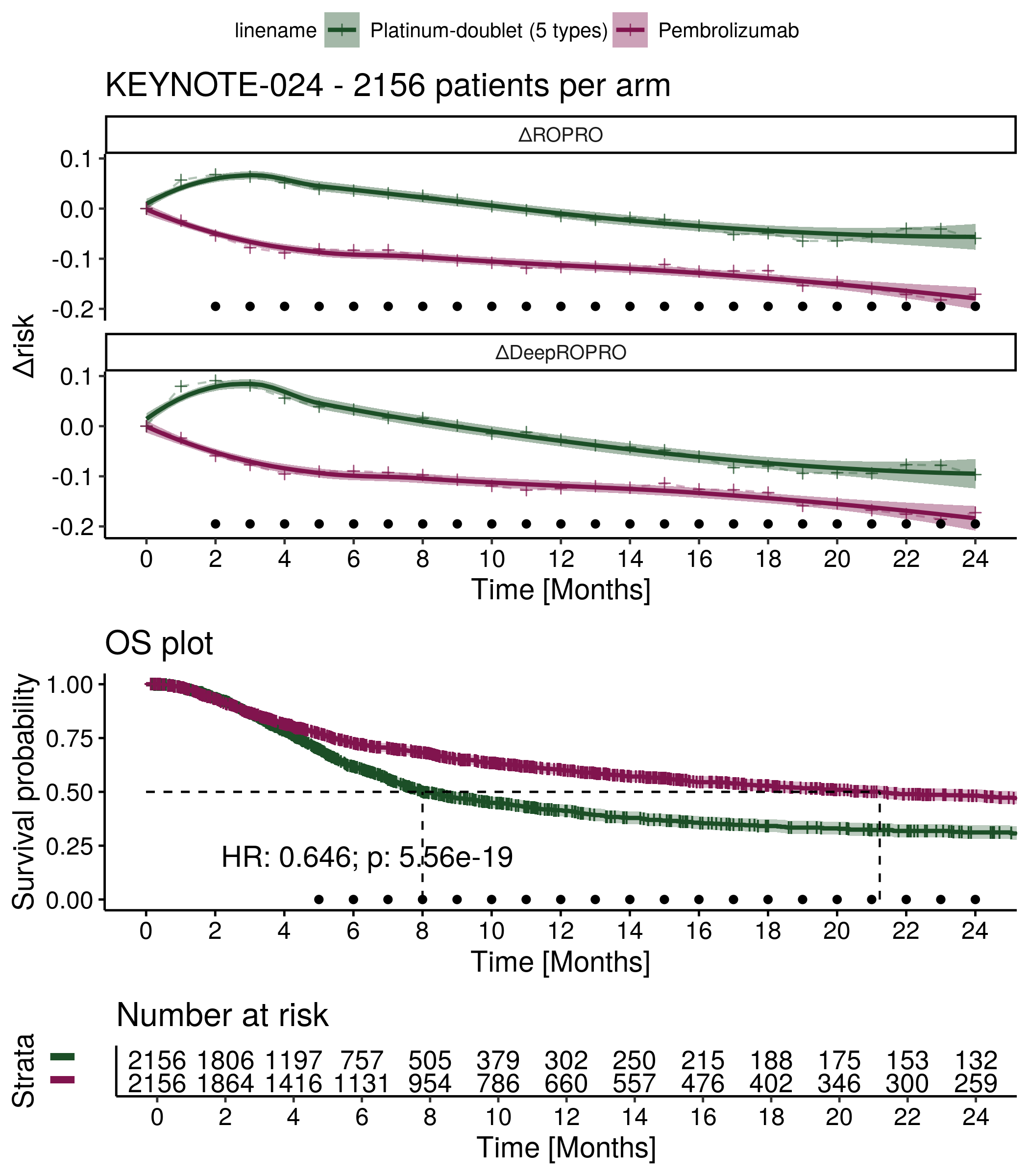

Supplementary Figure 5: ΔRisk and Kaplan Meier plots for the KEYNOTE-042 clinical trial. The HR is referent to the treatment effect of Pembrolizumab.

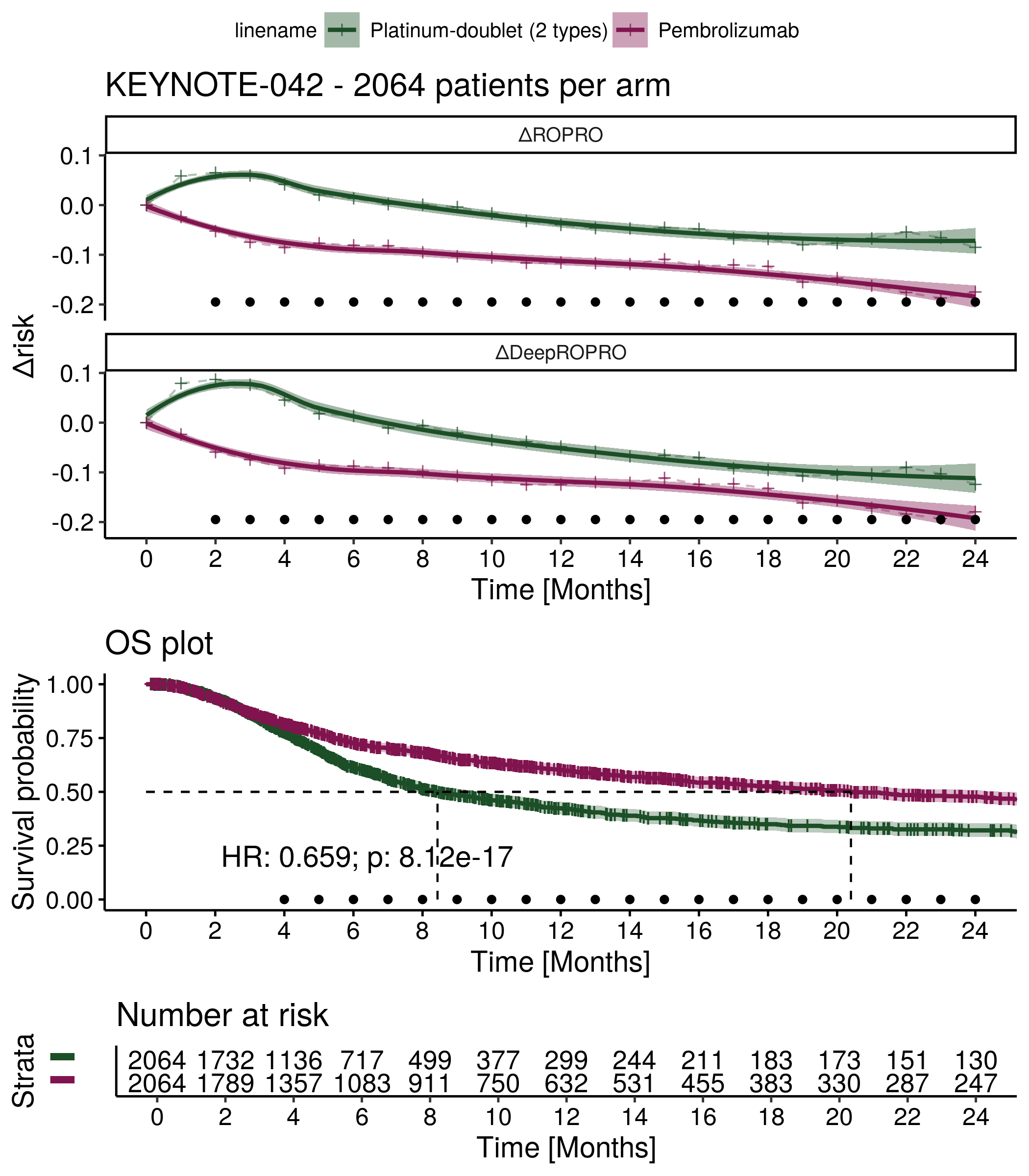

Supplementary Figure 6: ΔRisk and Kaplan Meier plots for the KEYNOTE-407 clinical trial. The HR is referent to the treatment effect of Pembrolizumab-combo.

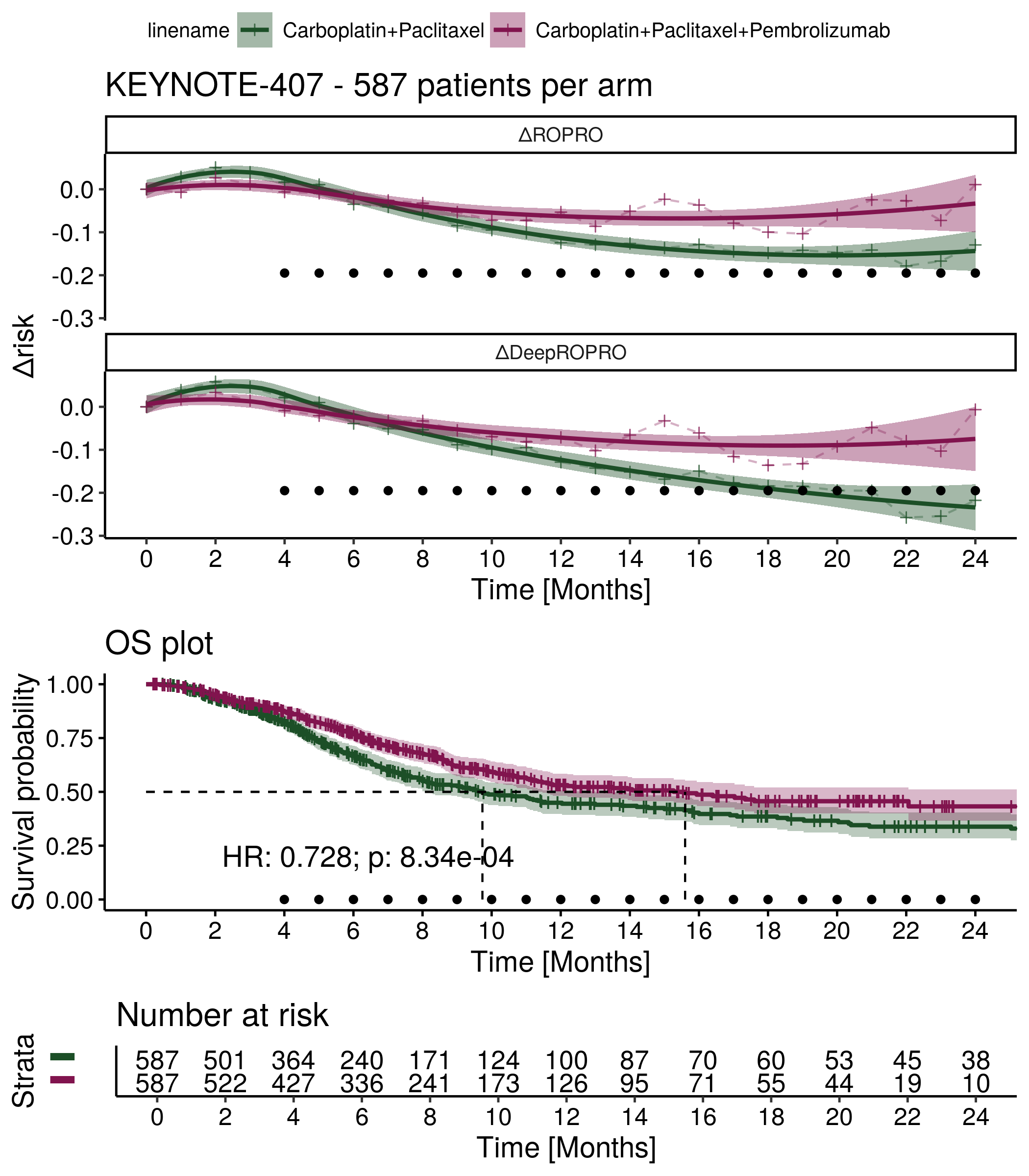

Supplementary Figure 7: ΔRisk and Kaplan Meier plots for the PRONOUNCE clinical trial. The HR is referent to the treatment effect of Bevacizumab+Carboplatin+Paclitaxel.

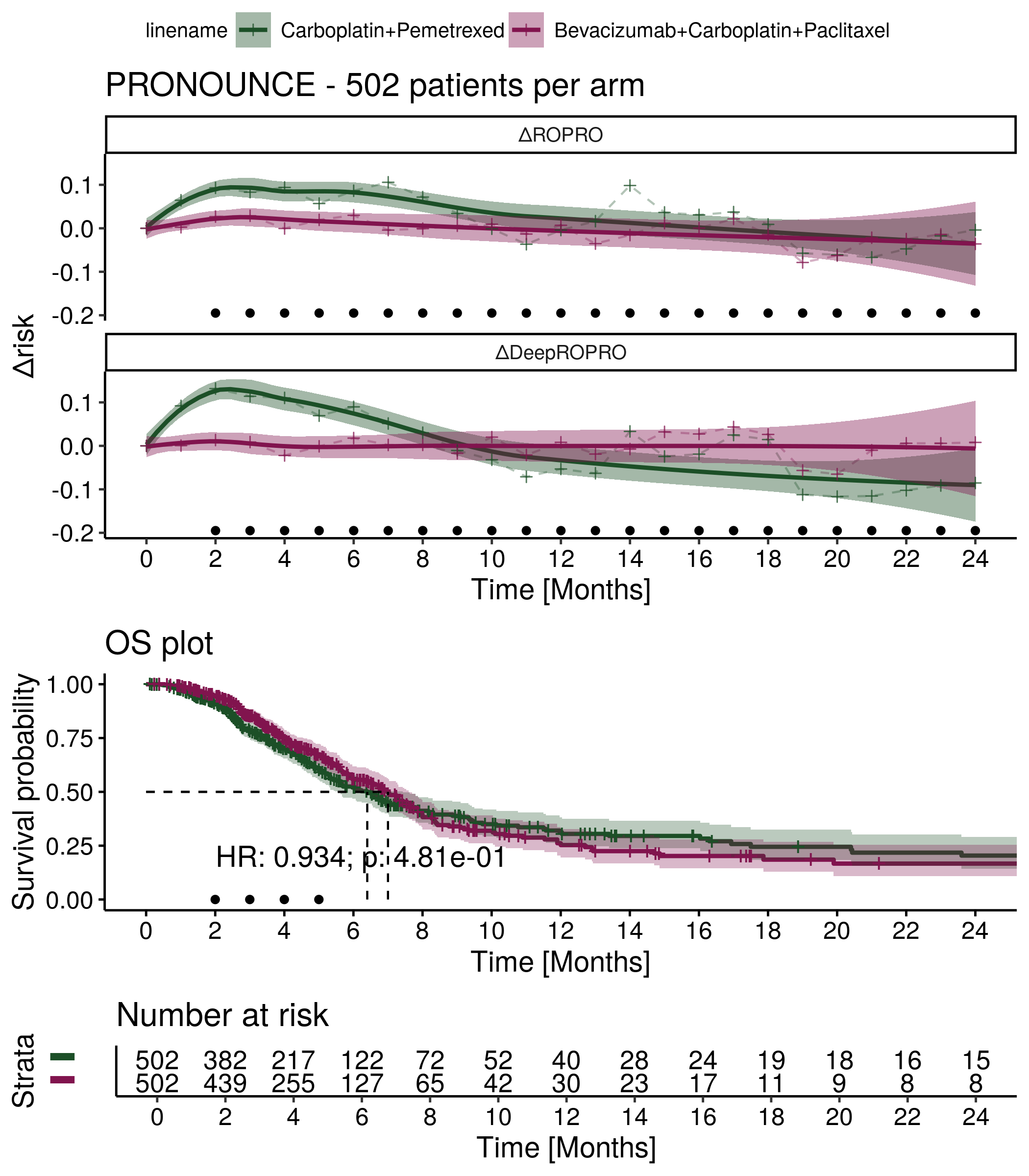

Supplementary Figure 8: ΔRisk and Kaplan Meier plots for the PointBreak clinical trial. The HR is referent to the treatment effect of Bevacizumab+Carboplatin+Paclitaxel.

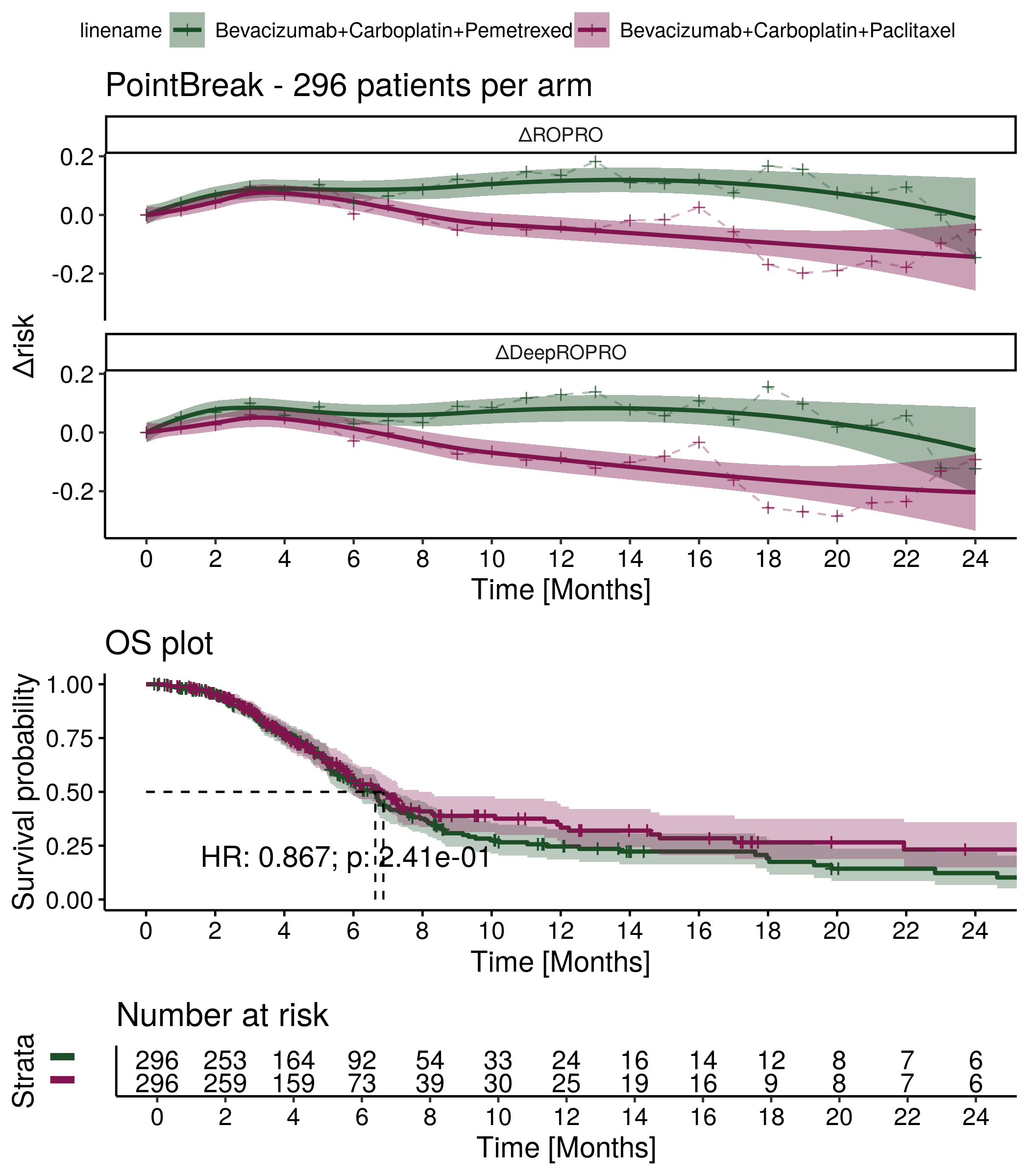

Supplementary Figure 9: ΔRisk and Kaplan Meier plots for the PROFILE 1014 clinical trial. The HR is referent to the treatment effect of Crizotinib.

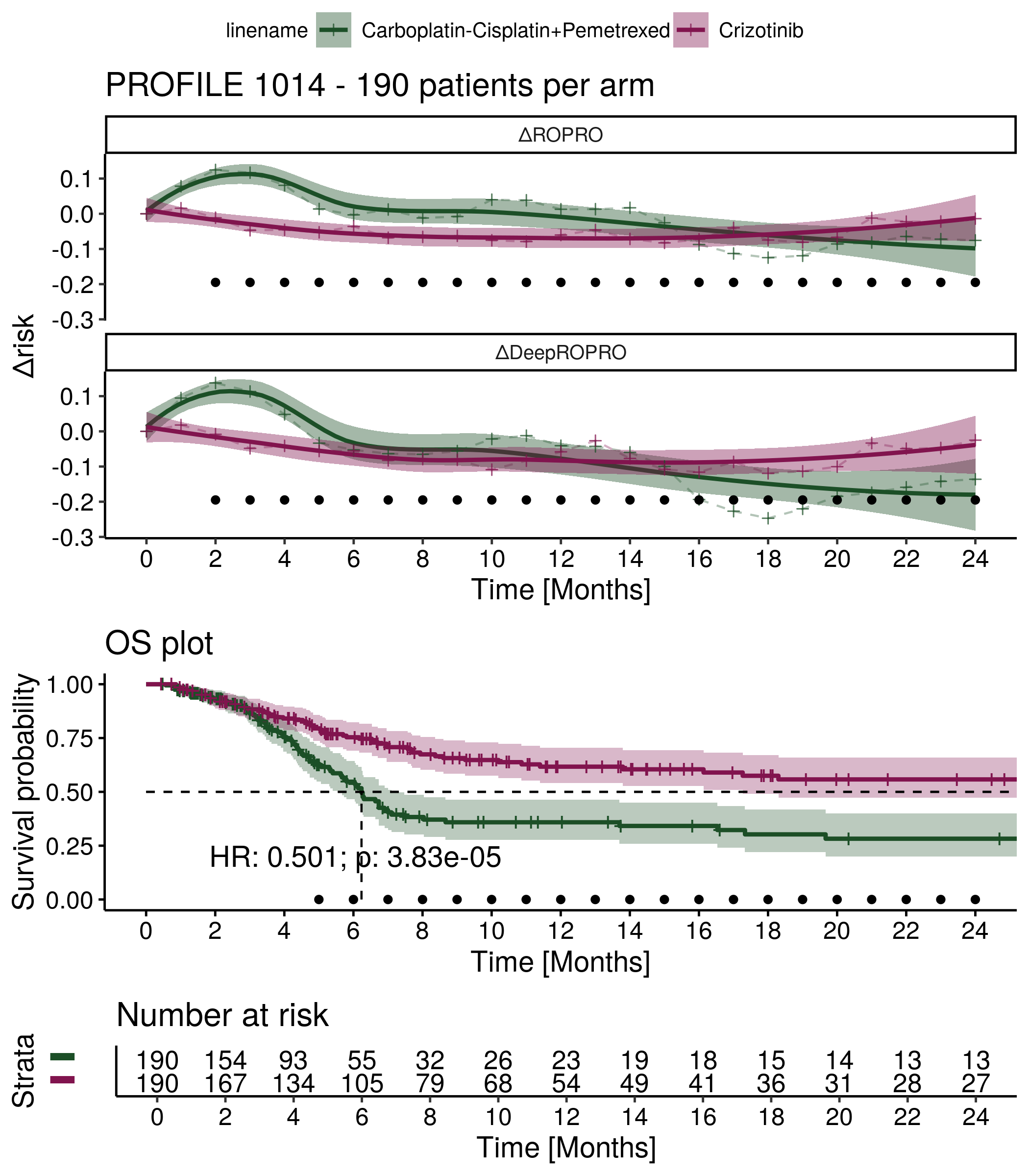

Supplementary Figure 10: ΔRisk and Kaplan Meier plots for the FLAURA clinical trial. The HR is referent to the treatment effect of Osimertinib.

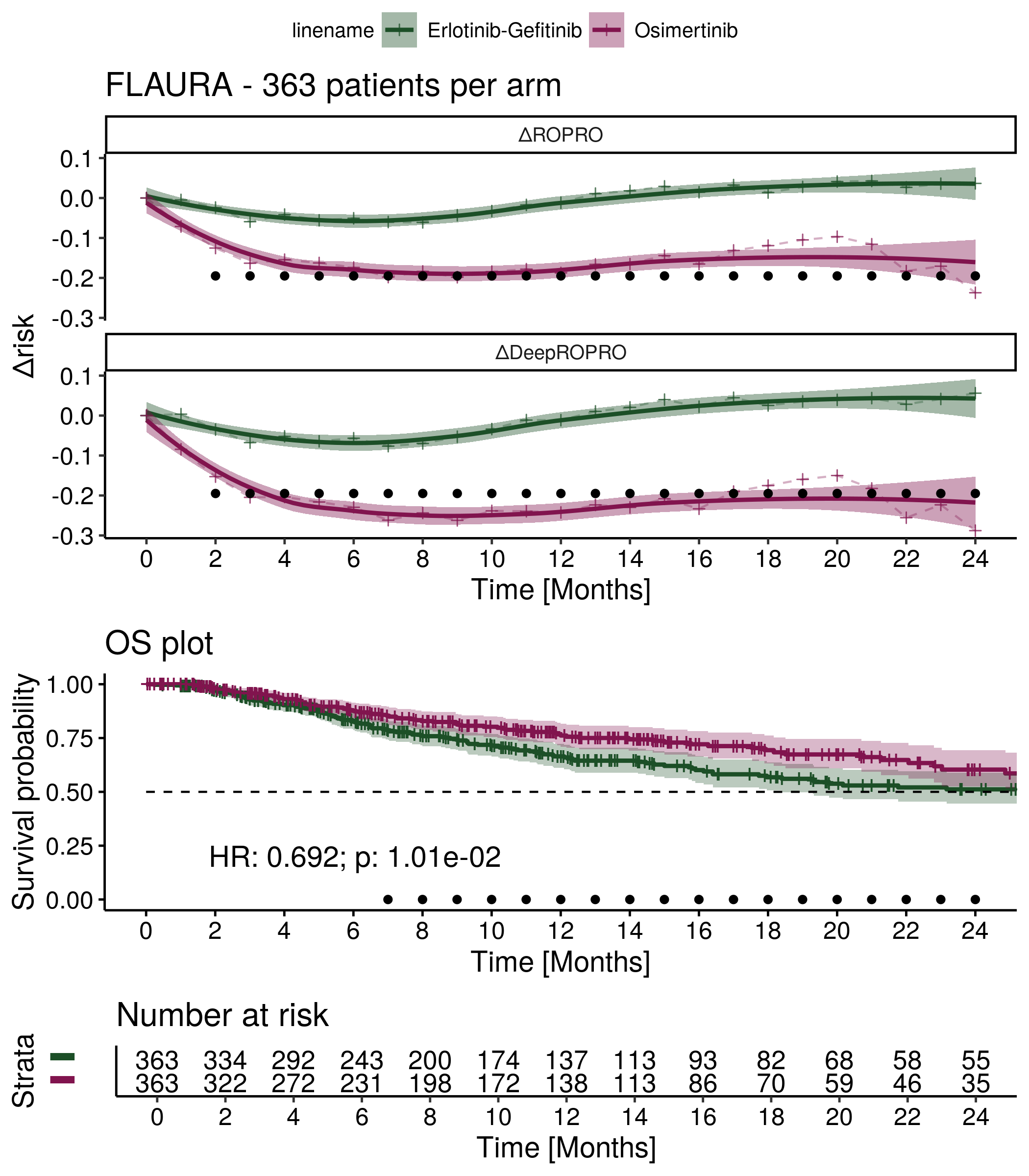

Supplementary Figure 11: ΔRisk and Kaplan Meier plots for the LUX-Lung 3+6 clinical trial. The HR is referent to the treatment effect of Afatinib.

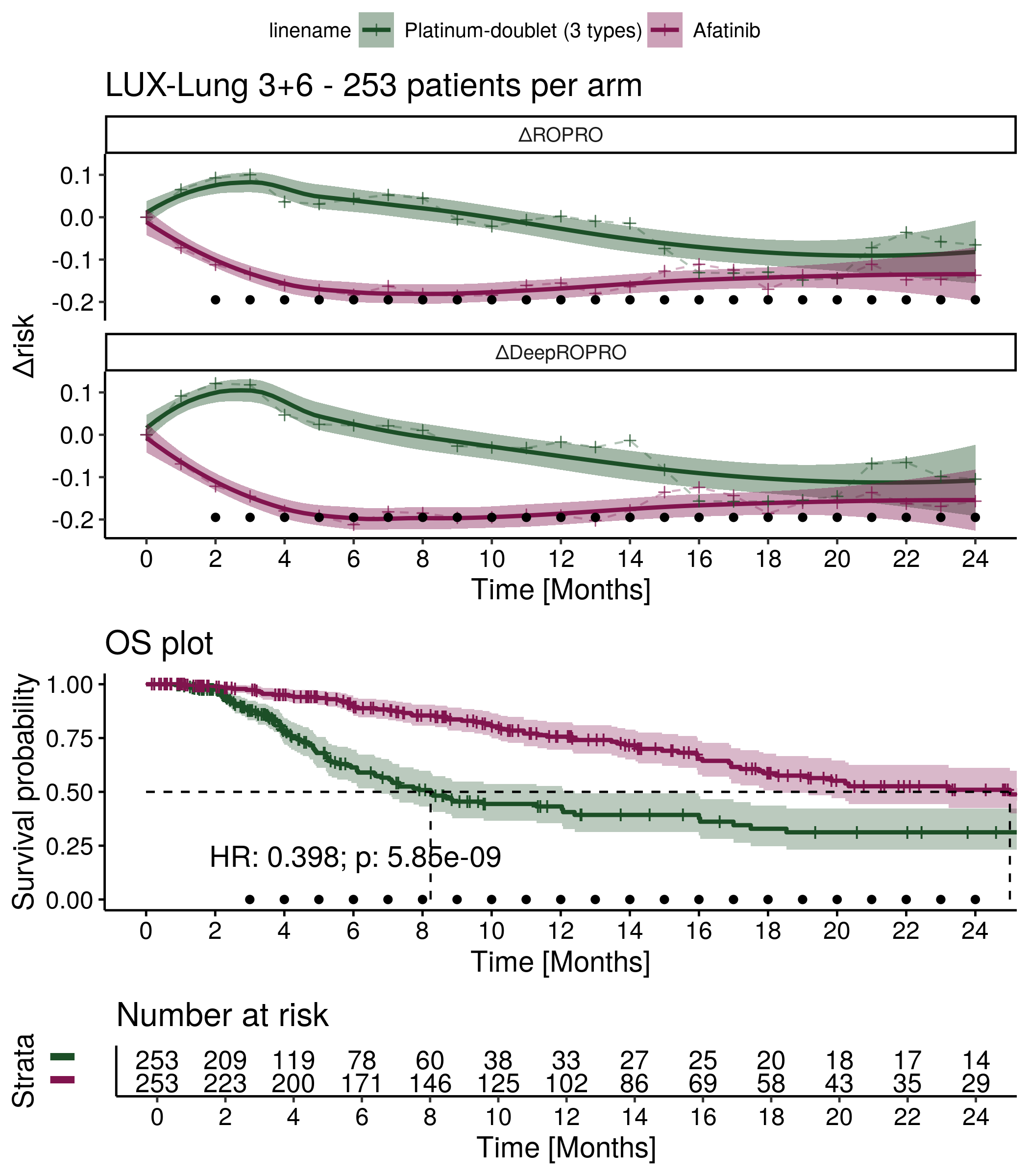

Supplementary Figure 12: ΔRisk and Kaplan Meier plots for the NCT00540514 clinical trial. The HR is referent to the treatment effect of Carboplatin+nab-Paclitaxel.

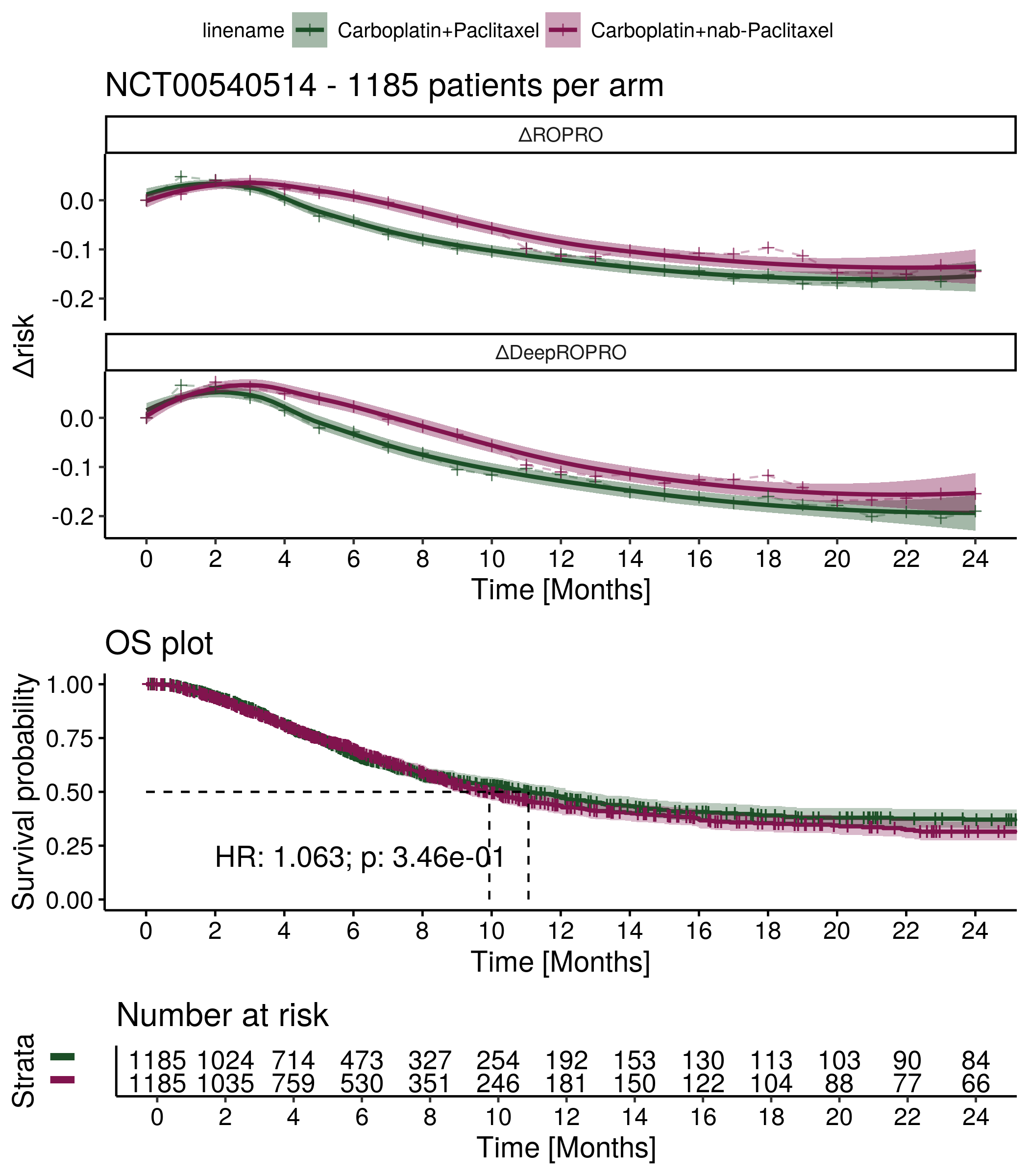

Supplementary Figure 13: ΔRisk and Kaplan Meier plots for the AURA3 clinical trial. The HR is referent to the treatment effect of Osimertinib.

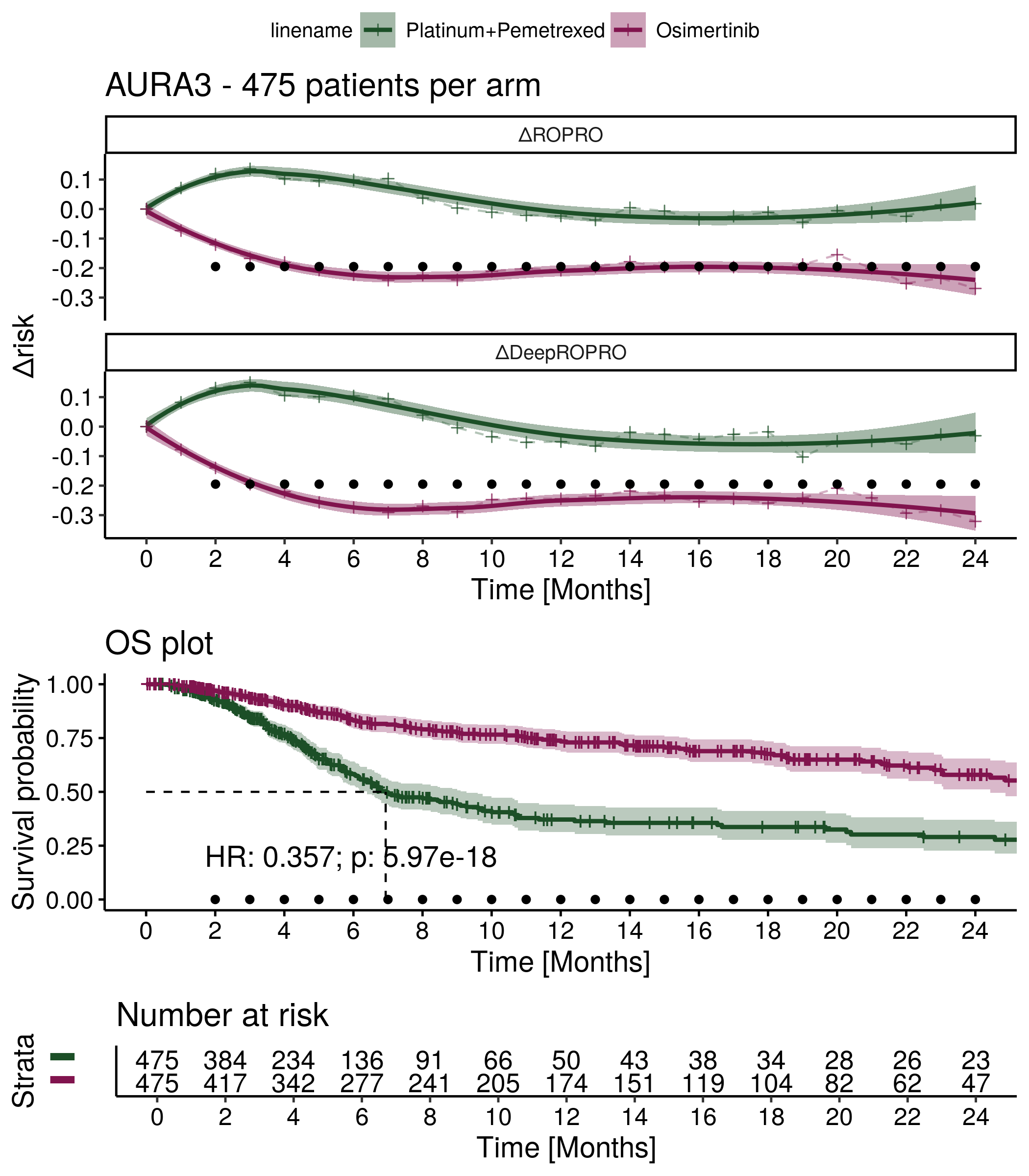

Supplementary Figure 14: ΔRisk and Kaplan Meier plots for the NCT00520676 clinical trial. The HR is referent to the treatment effect of Carboplatin+Docetaxel.

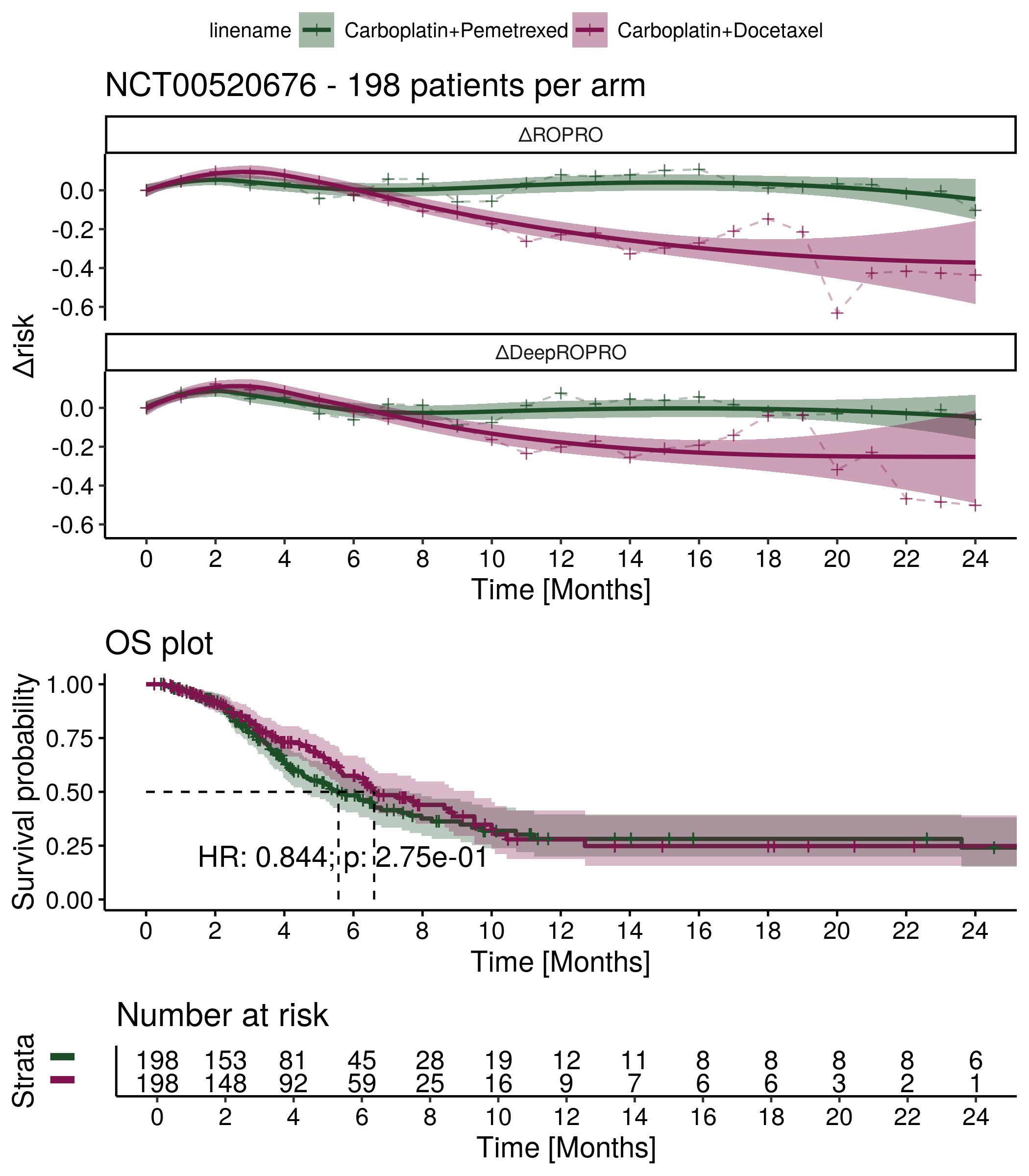

##### 1.1.2. Lower caliper ΔRisk and Overall Survival plots

Supplementary Figure 15: ΔRisk and Kaplan Meier plots for the KEYNOTE-189 clinical trial. The HR is referent to the treatment effect of Platinum+Pemetrexed.

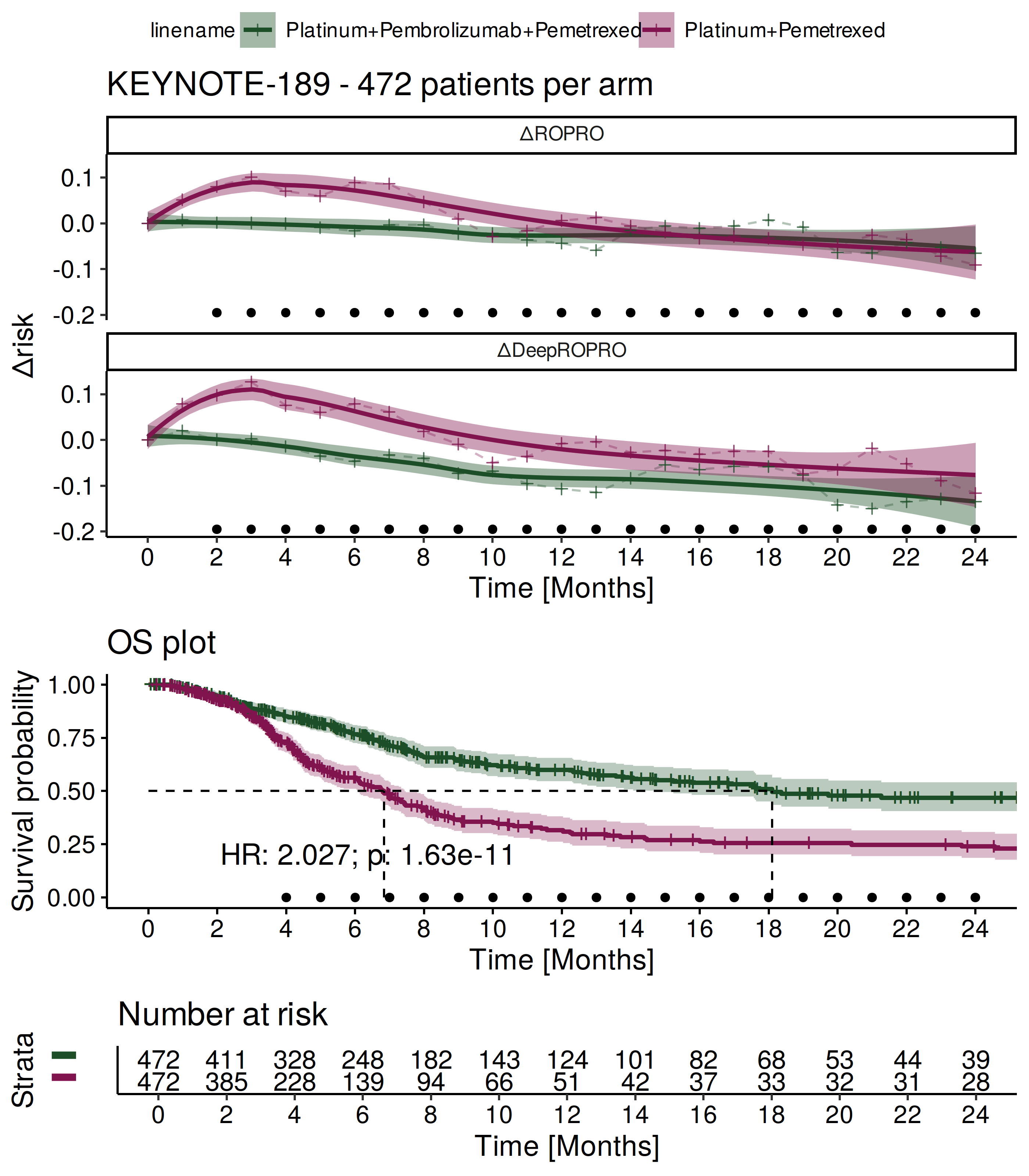

Supplementary Figure 16: ΔRisk and Kaplan Meier plots for the KEYNOTE-024 clinical trial. The HR is referent to the treatment effect of Pembrolizumab.

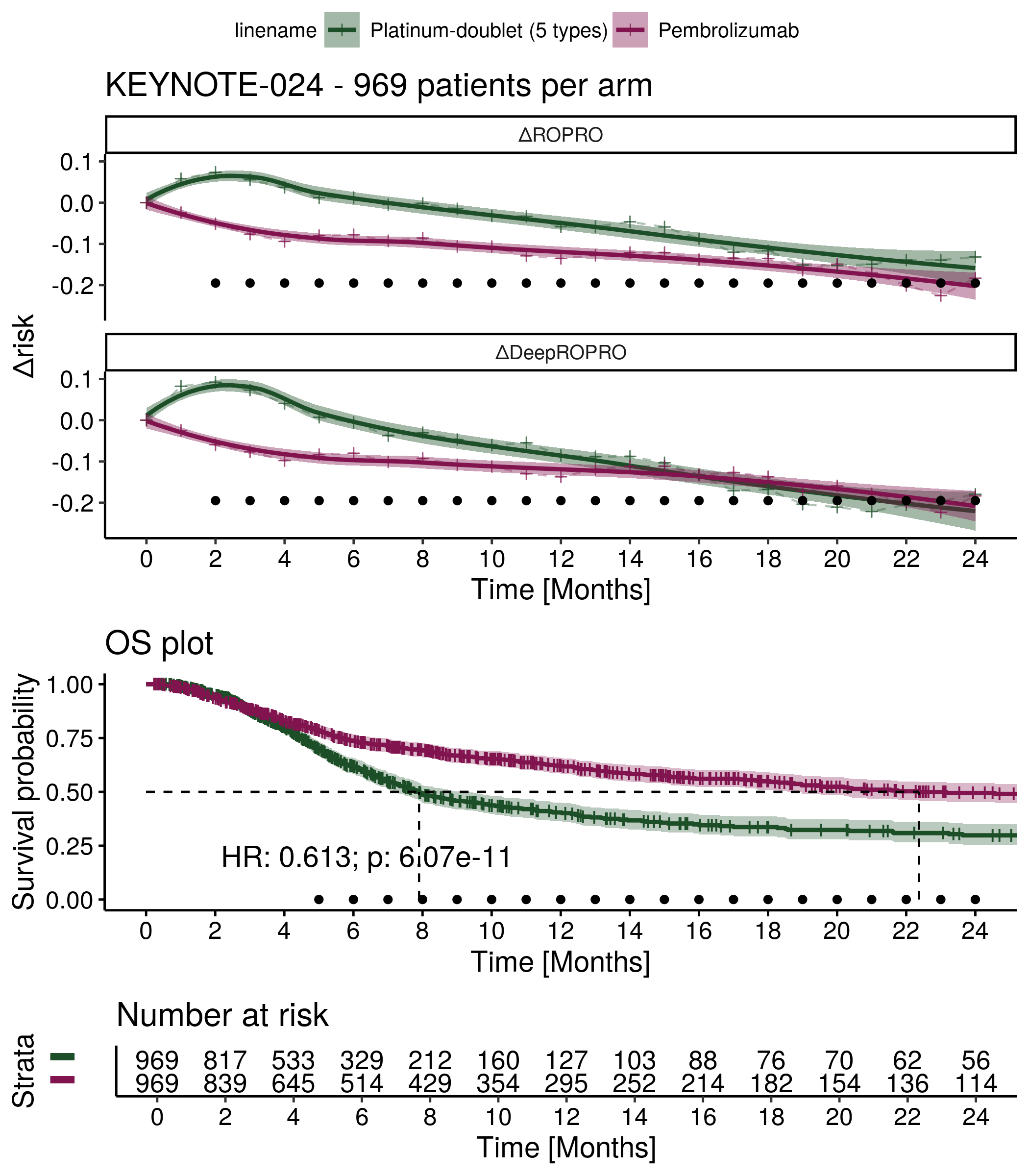

Supplementary Figure 17: ΔRisk and Kaplan Meier plots for the KEYNOTE-042 clinical trial. The HR is referent to the treatment effect of Pembrolizumab.

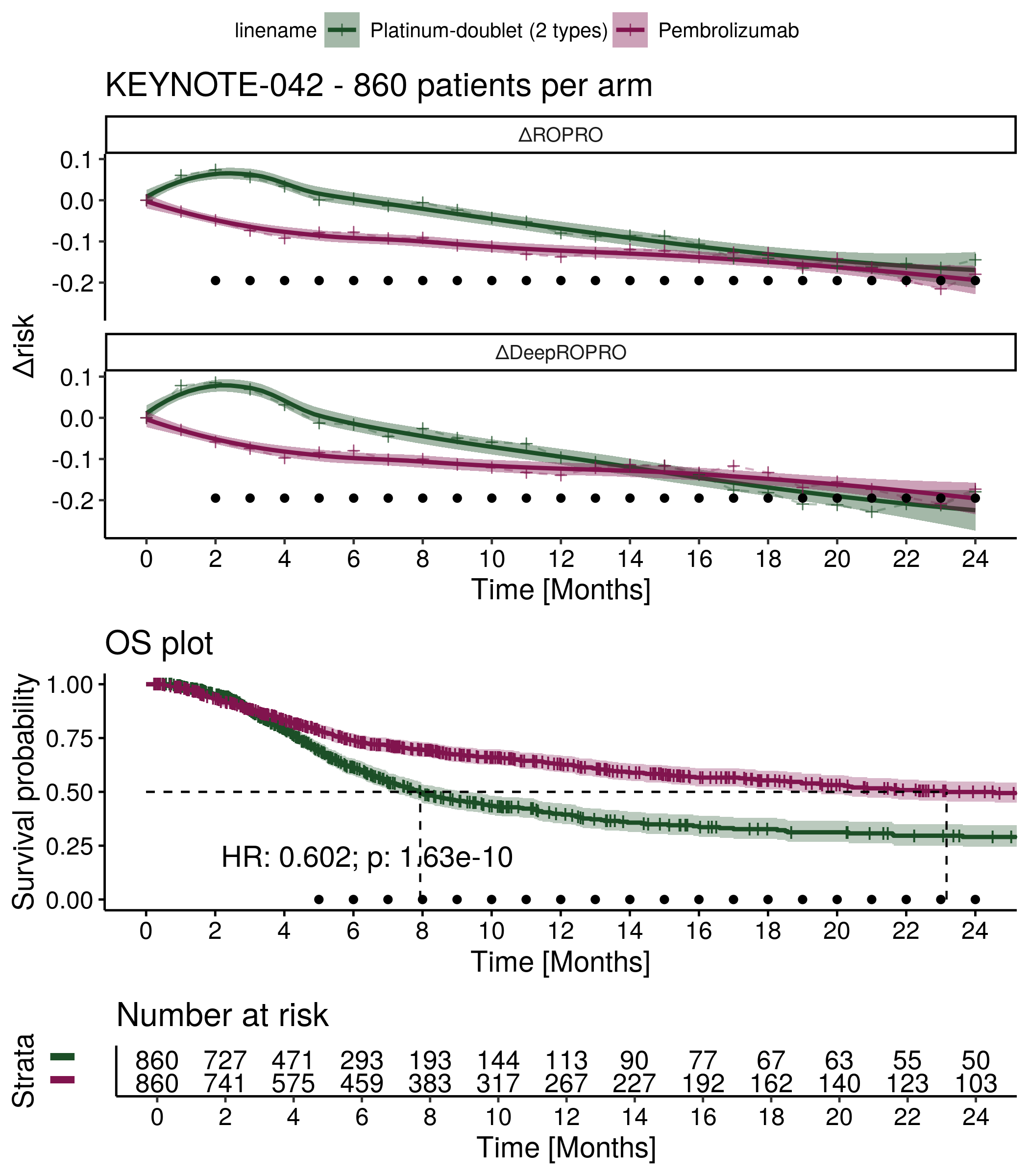

Supplementary Figure 18: ΔRisk and Kaplan Meier plots for the KEYNOTE-407 clinical trial. The HR is referent to the treatment effect of Pembrolizumab-combo.

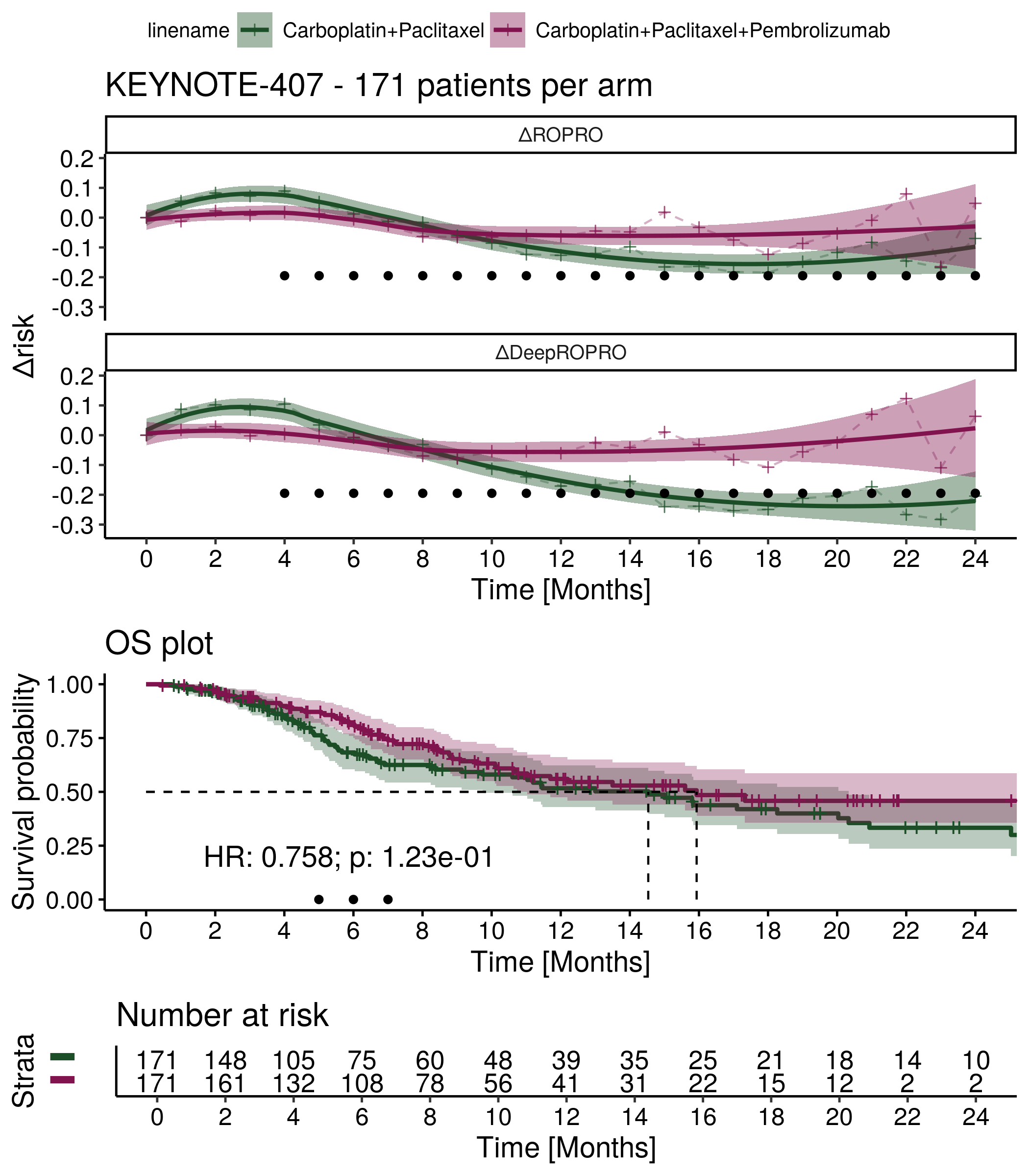

Supplementary Figure 19: ΔRisk and Kaplan Meier plots for the PRONOUNCE clinical trial. The HR is referent to the treatment effect of Bevacizumab+Carboplatin+Paclitaxel.

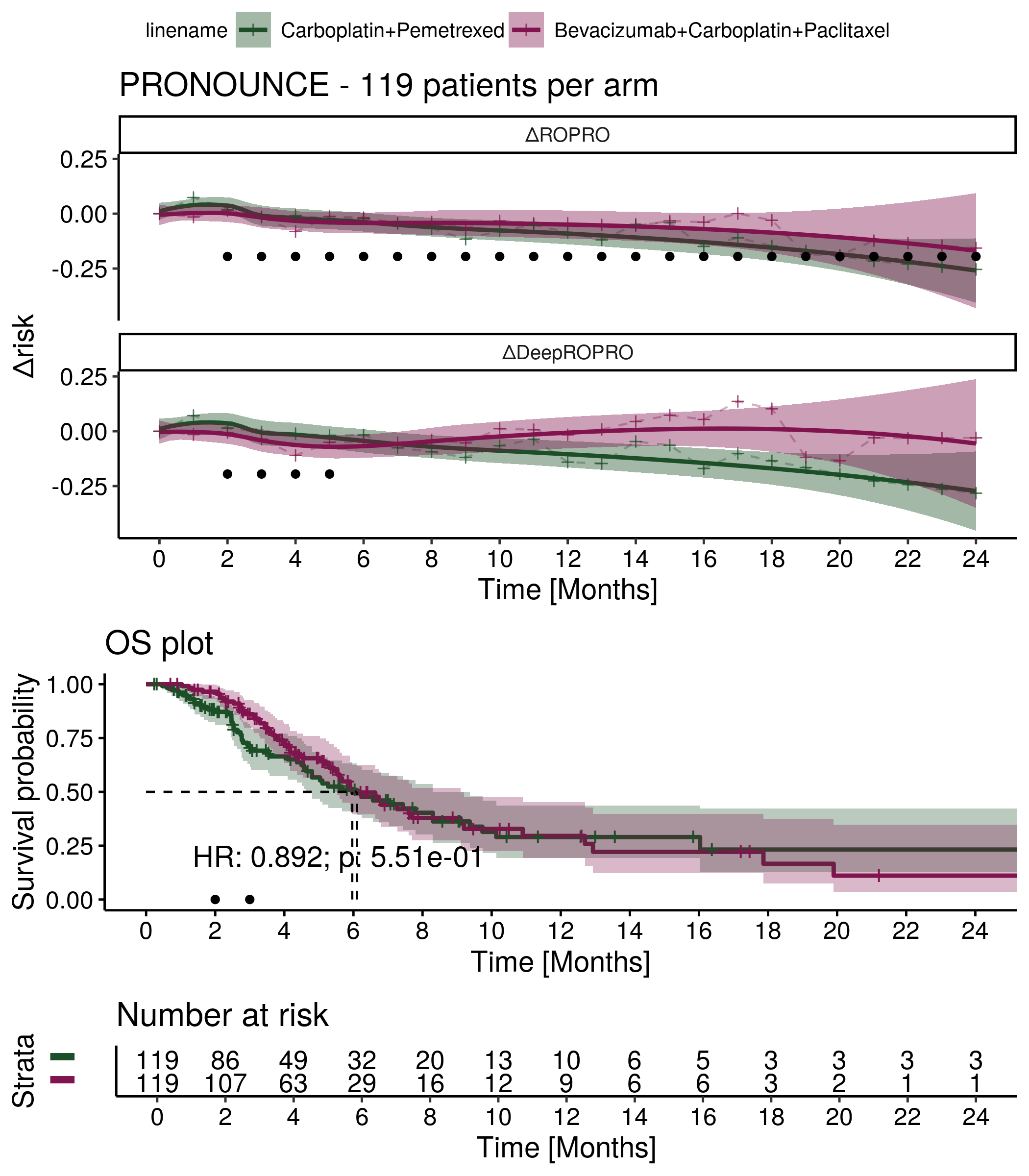

Supplementary Figure 20: ΔRisk and Kaplan Meier plots for the PointBreak clinical trial. The HR is referent to the treatment effect of Bevacizumab+Carboplatin+Paclitaxel.

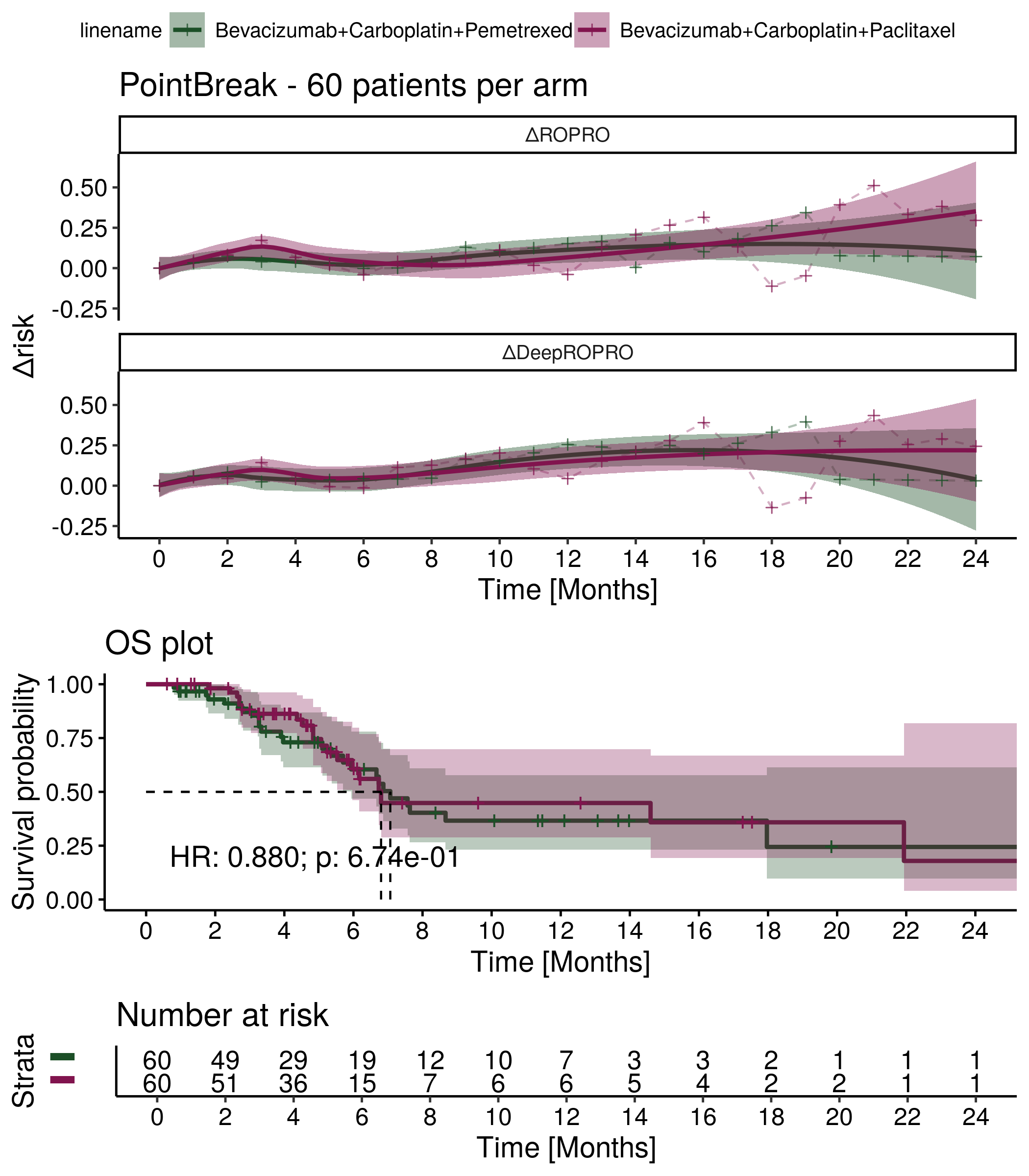

Supplementary Figure 21: ΔRisk and Kaplan Meier plots for the PROFILE 1014 clinical trial. The HR is referent to the treatment effect of Crizotinib.

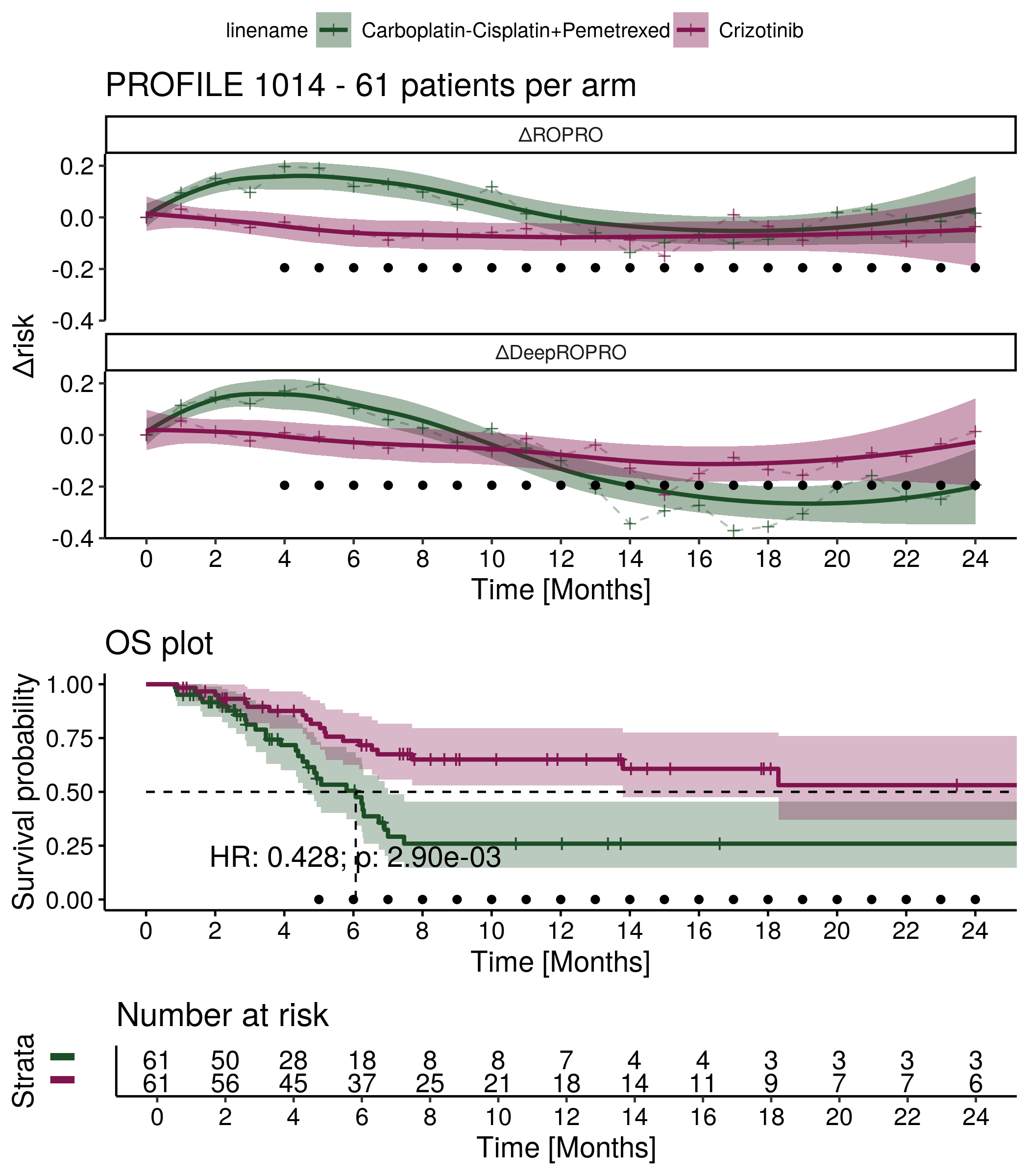

Supplementary Figure 22: ΔRisk and Kaplan Meier plots for the FLAURA clinical trial. The HR is referent to the treatment effect of Osimertinib.

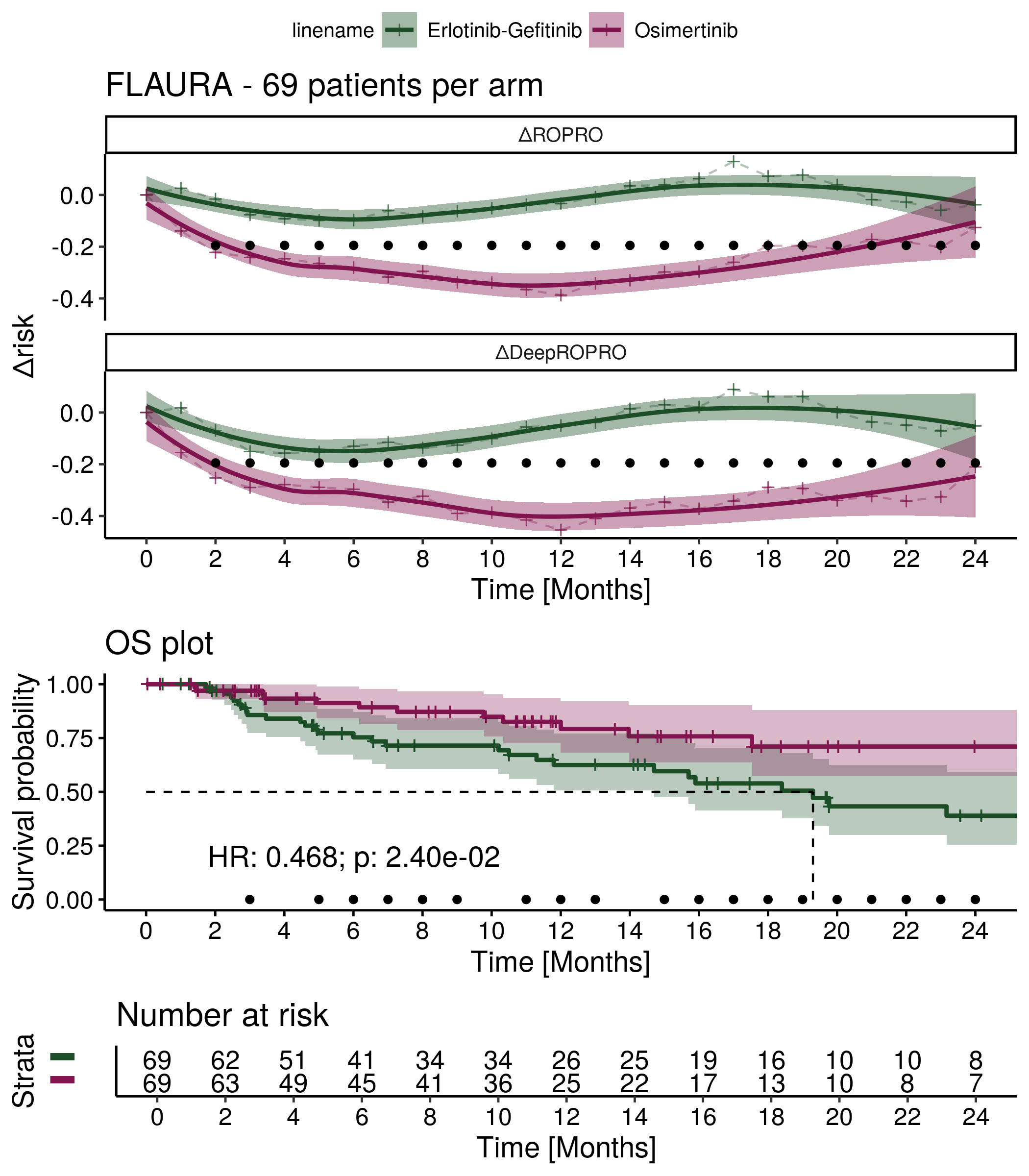

Supplementary Figure 23: ΔRisk and Kaplan Meier plots for the LUX-Lung 3+6 clinical trial. The HR is referent to the treatment effect of Afatinib.

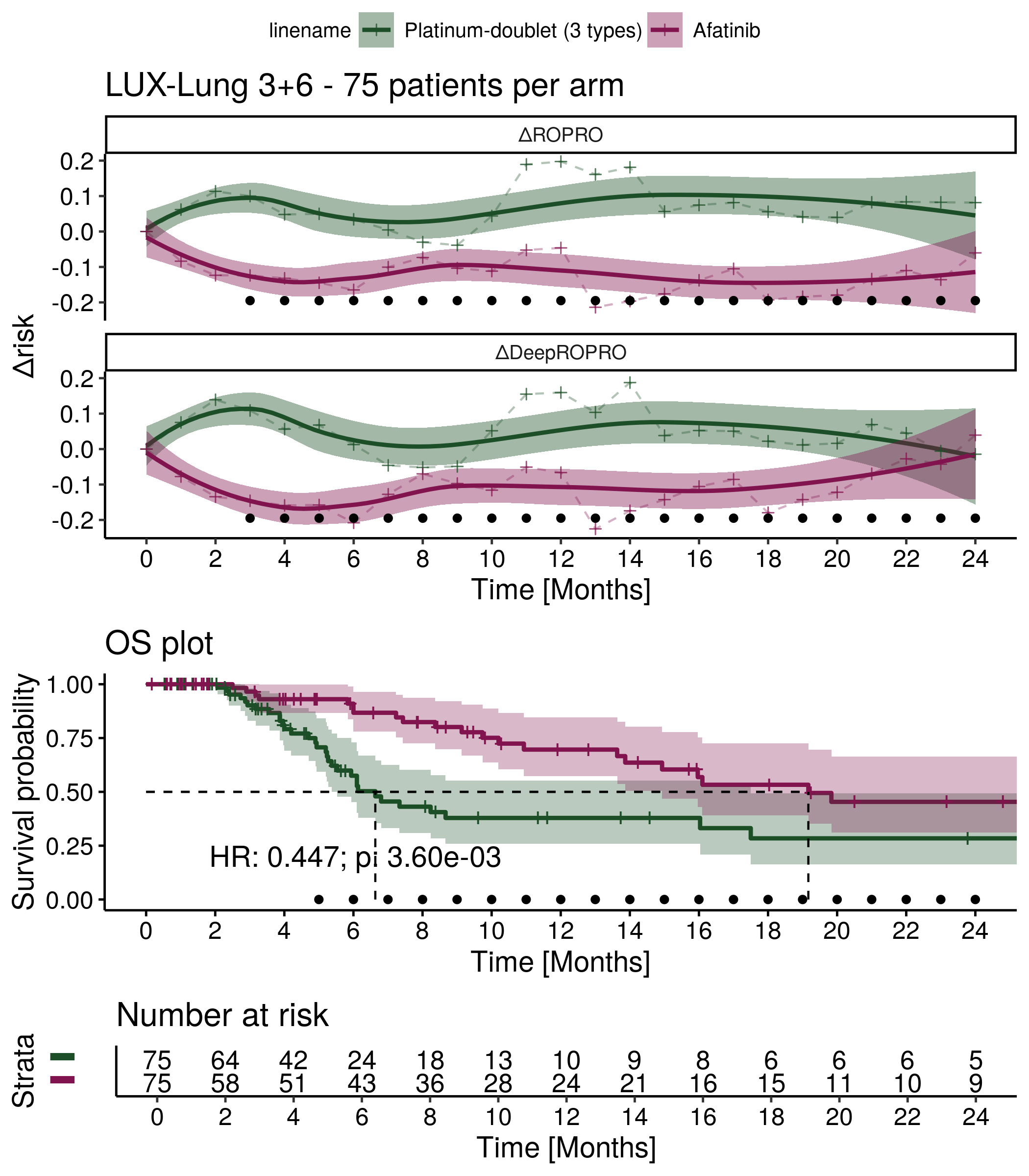

Supplementary Figure 24: ΔRisk and Kaplan Meier plots for the NCT00540514 clinical trial. The HR is referent to the treatment effect of Carboplatin+nab-Paclitaxel.

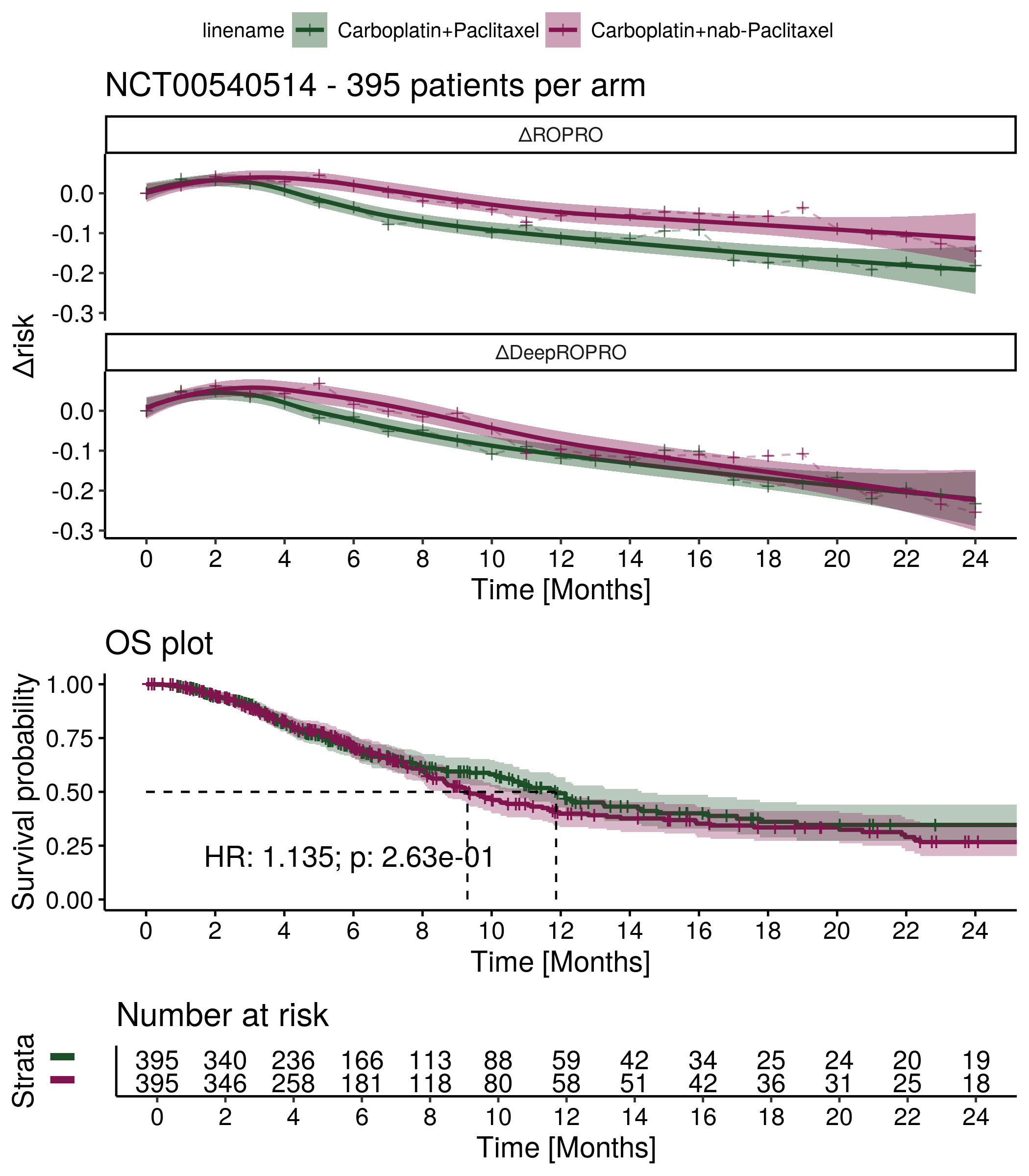

Supplementary Figure 25: ΔRisk and Kaplan Meier plots for the AURA3 clinical trial. The HR is referent to the treatment effect of Osimertinib.

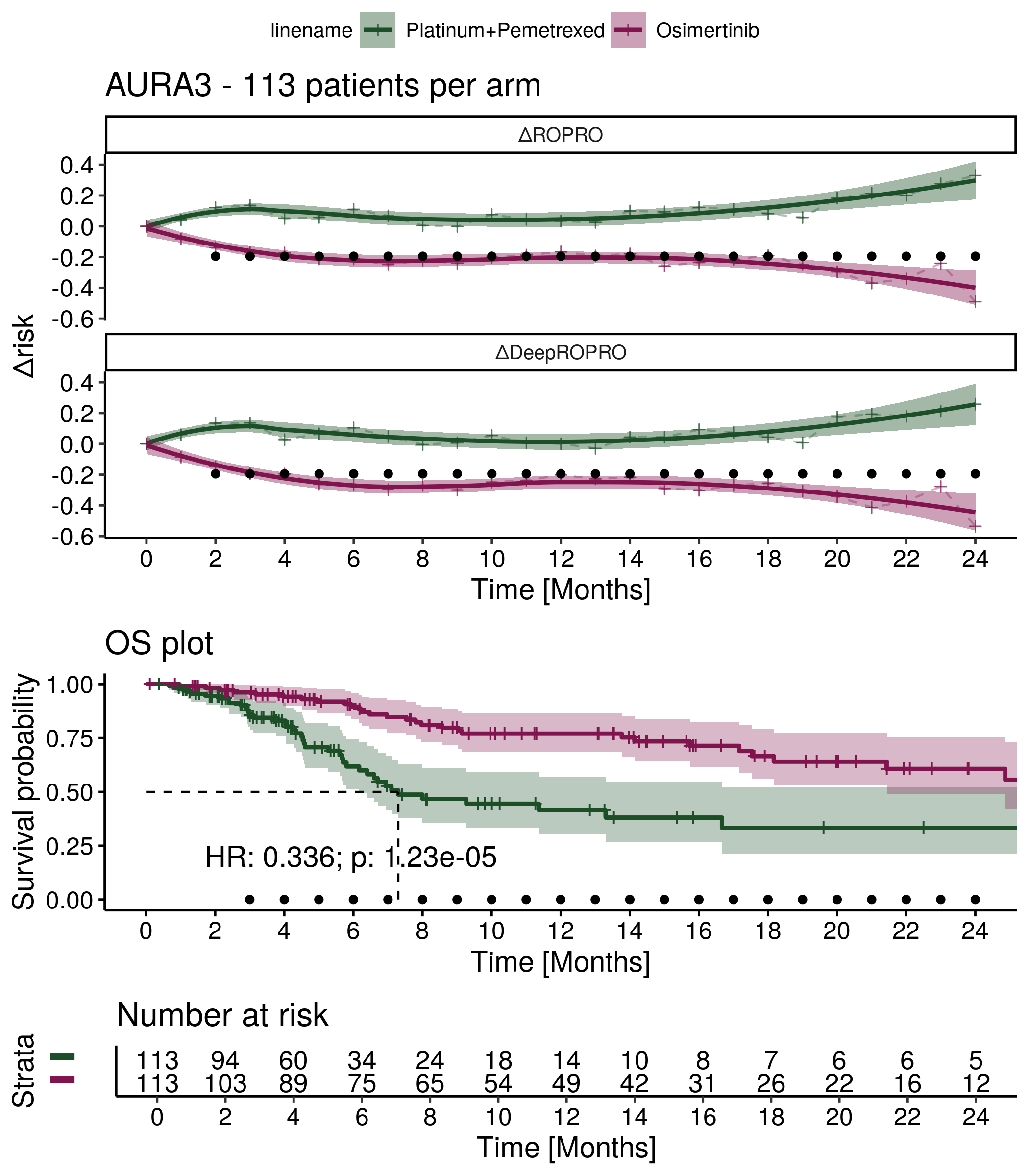

Supplementary Figure 26: ΔRisk and Kaplan Meier plots for the NCT00520676 clinical trial. The HR is referent to the treatment effect of Carboplatin+Docetaxel.

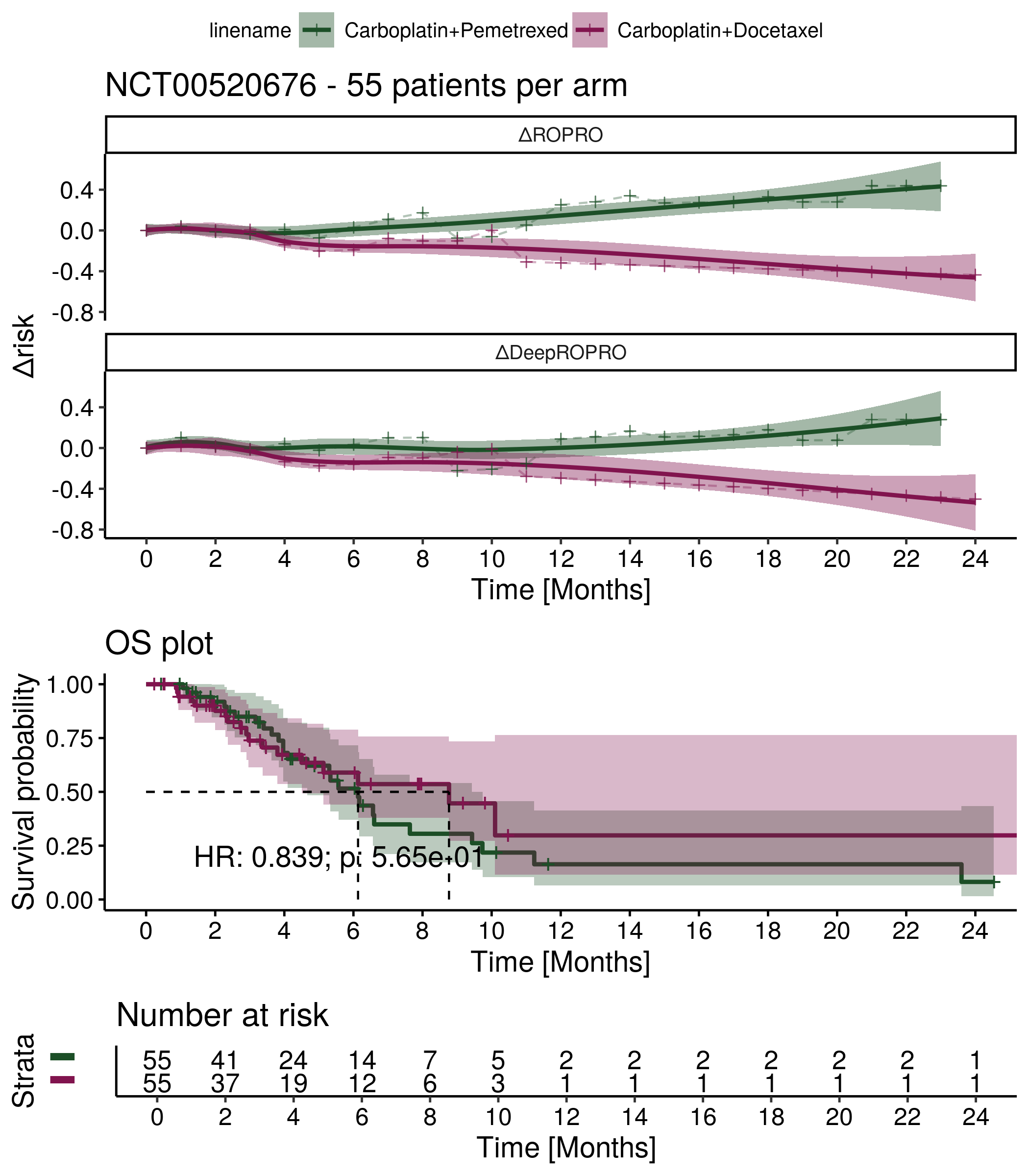

#### 1.2. ΔRisk and Overall Survival plots for each Atezolizumab (Roche) clinical trials (with treatment arm in the hazard function)

##### 1.2.1. ΔRisk, Overall Survival and Progression-free survival plots for the three Roche clinical trials

Supplementary Figure 27: ΔRisk and Kaplan Meier plots for the IMpower150 clinical trial. The HR is referent to the treatment effect of ABCP.

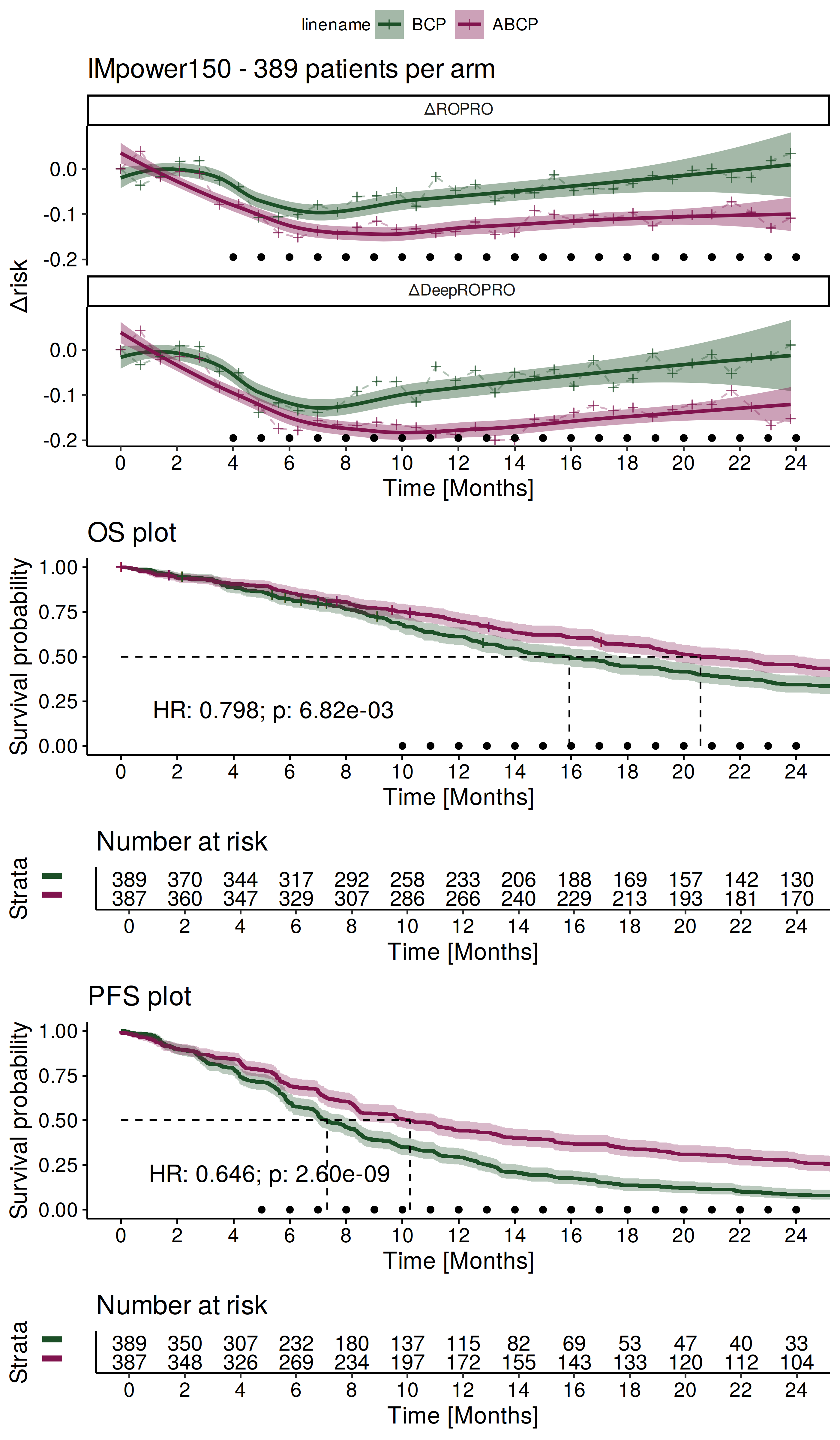

Supplementary Figure 28: ΔRisk and Kaplan Meier plots for the IMvigor211 clinical trial. The HR is referent to the treatment effect of cytotoxic chemotherapy.

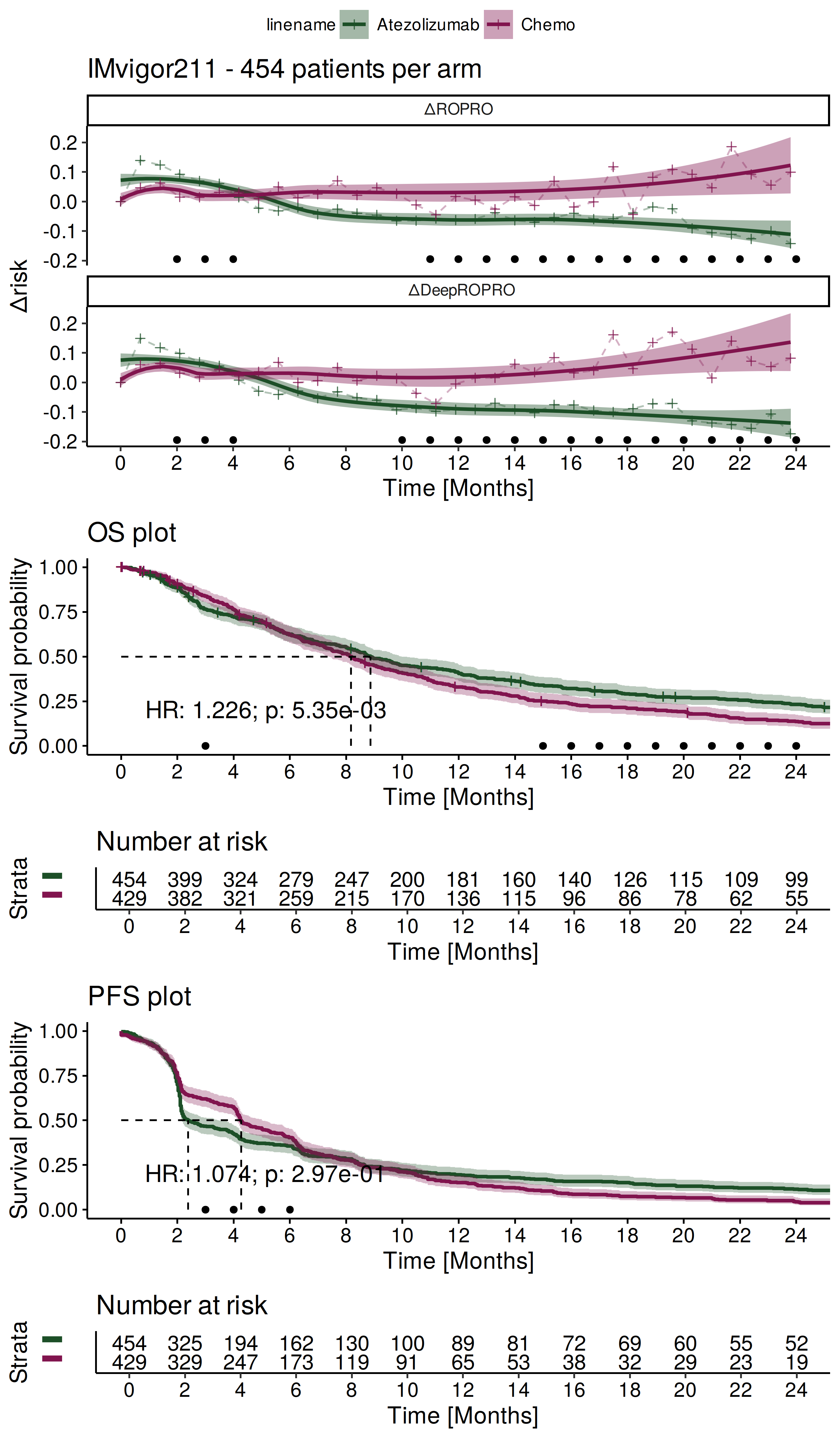

Supplementary Figure 29: ΔRisk and Kaplan Meier plots for the OAK clinical trial. The HR is referent to the treatment effect of Docetaxel.

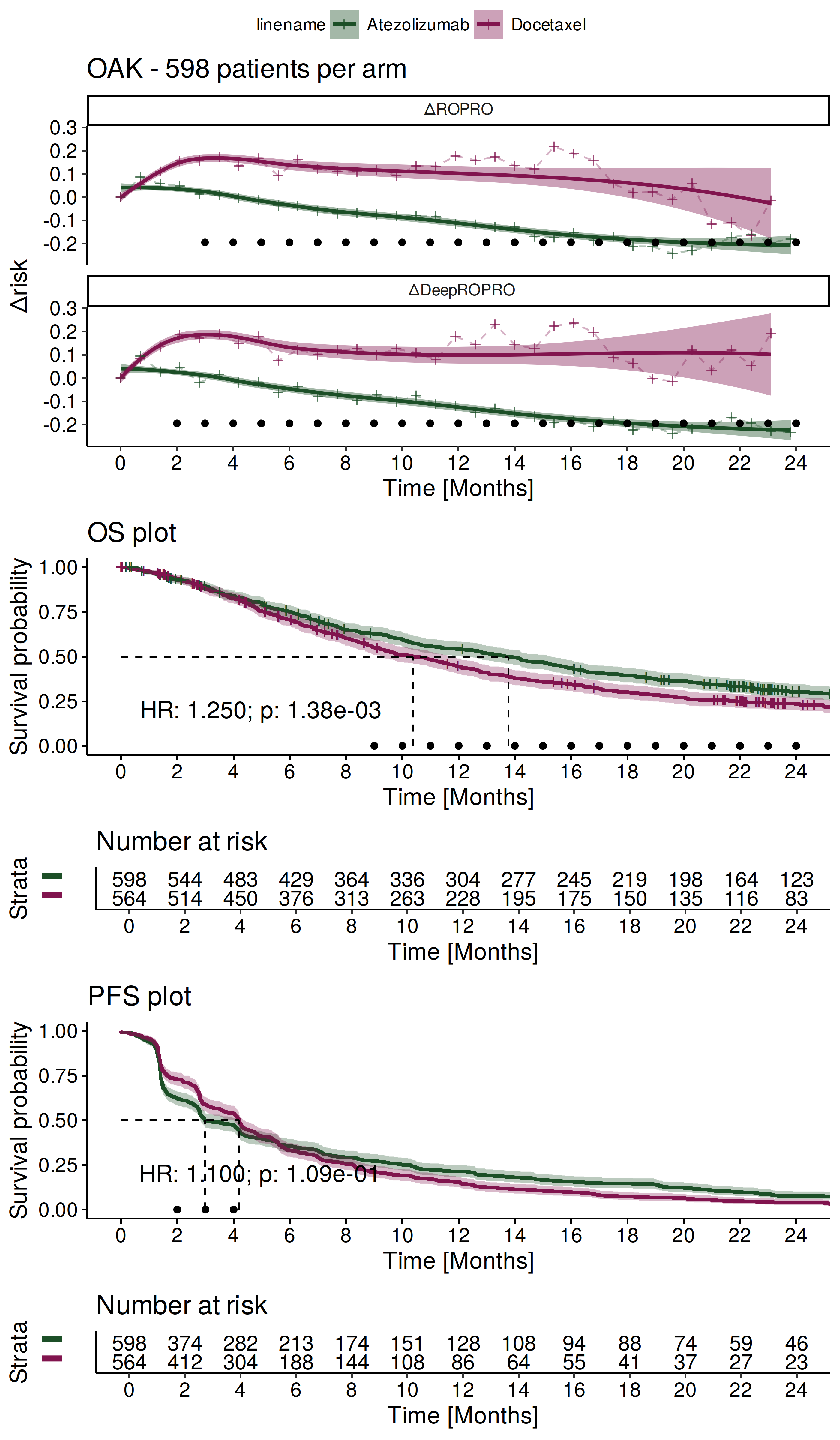

#### 1.3. ΔRisk, Overall Survival and adverse event plots for the PRONOUNCE clinical trial

##### 1.3.1. Higher caliper ΔRisk and Overall Survival plots

Supplementary Figure 30: ΔRisk, Kaplan Meier, and adverse events plots for the PRONOUNCE clinical trial.

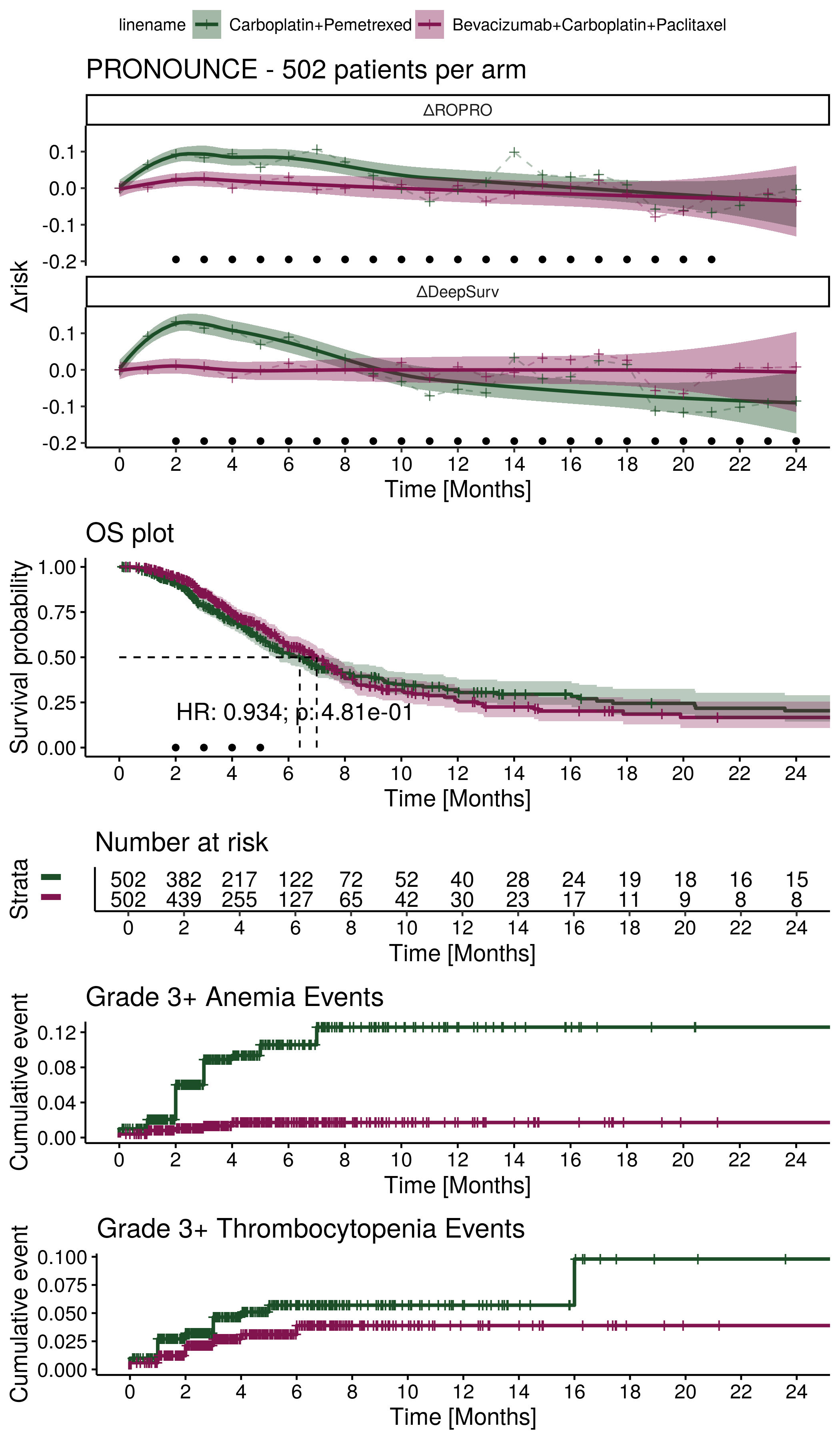

##### 1.3.2. Lower caliper ΔRisk and Overall Survival plots

Supplementary Figure 31: ΔRisk, Kaplan Meier, and adverse events plots for the PRONOUNCE clinical trial.

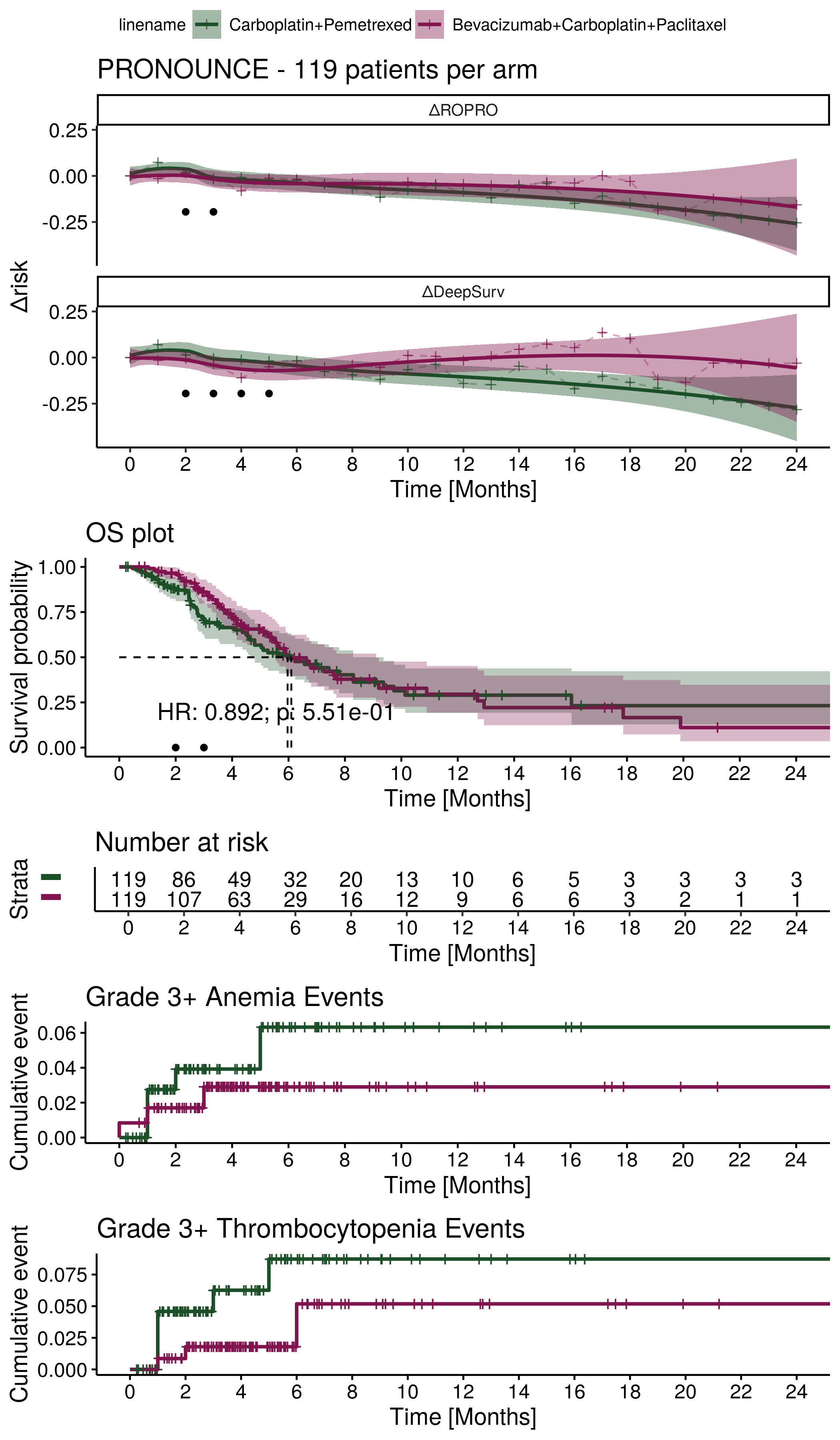

#### 1.4. Other supplementary figures

Supplementary Figure 32: ΔRisk and OS Kaplan Meier curves for the FLAURA clinical trial. The ΔRisk plots include both the smoothed (average) ΔRisk curve and the individual patient ΔRisk curves. The HR is referent to the treatment effect of Osimertinib.

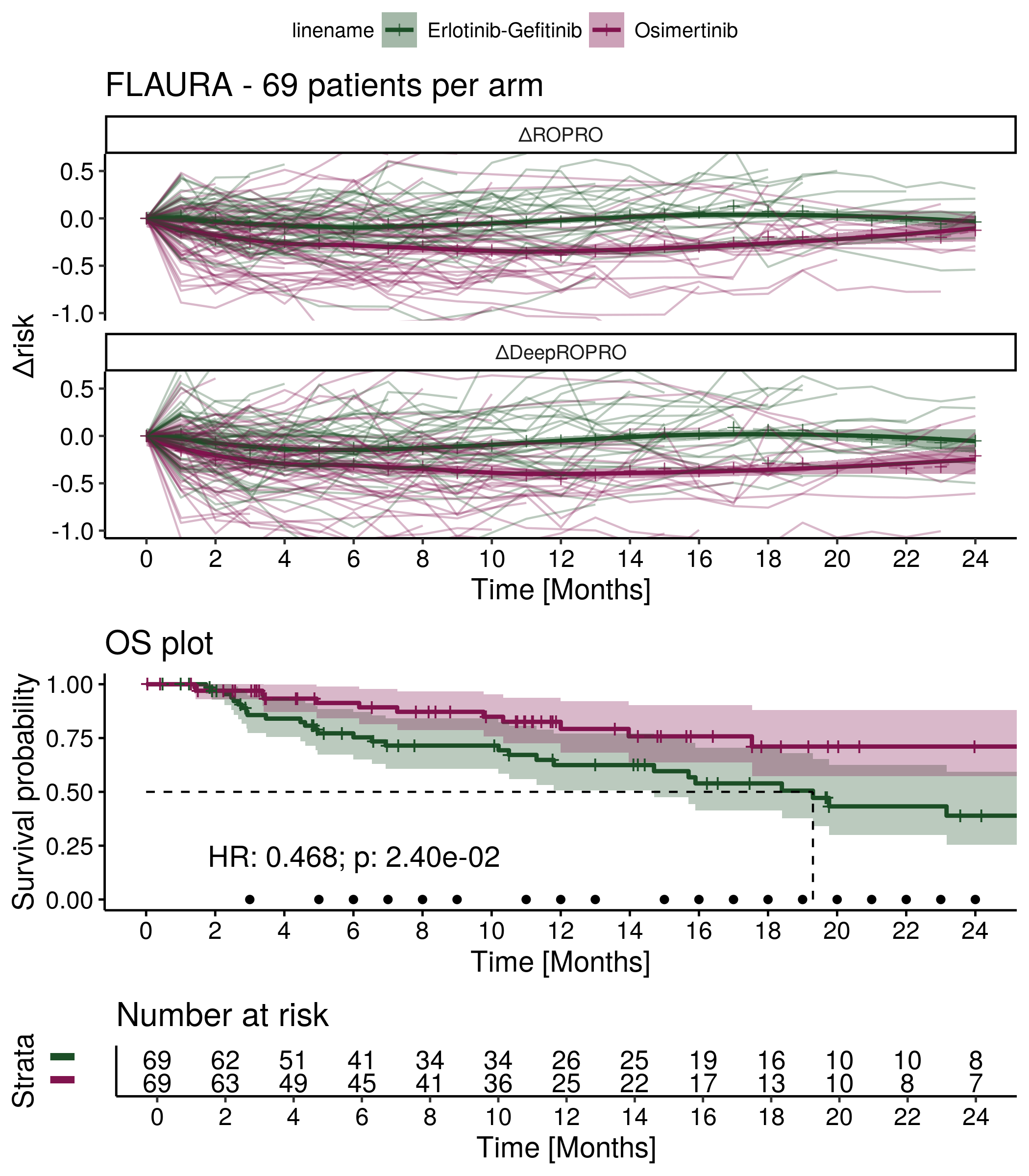

Supplementary Figure 33: OS hazard ratios at 24 months versus the ΔROPRO coefficients at 3 months for the lower caliper analysis. The included regression line considers that $\boldsymbol{\gamma=0}$.

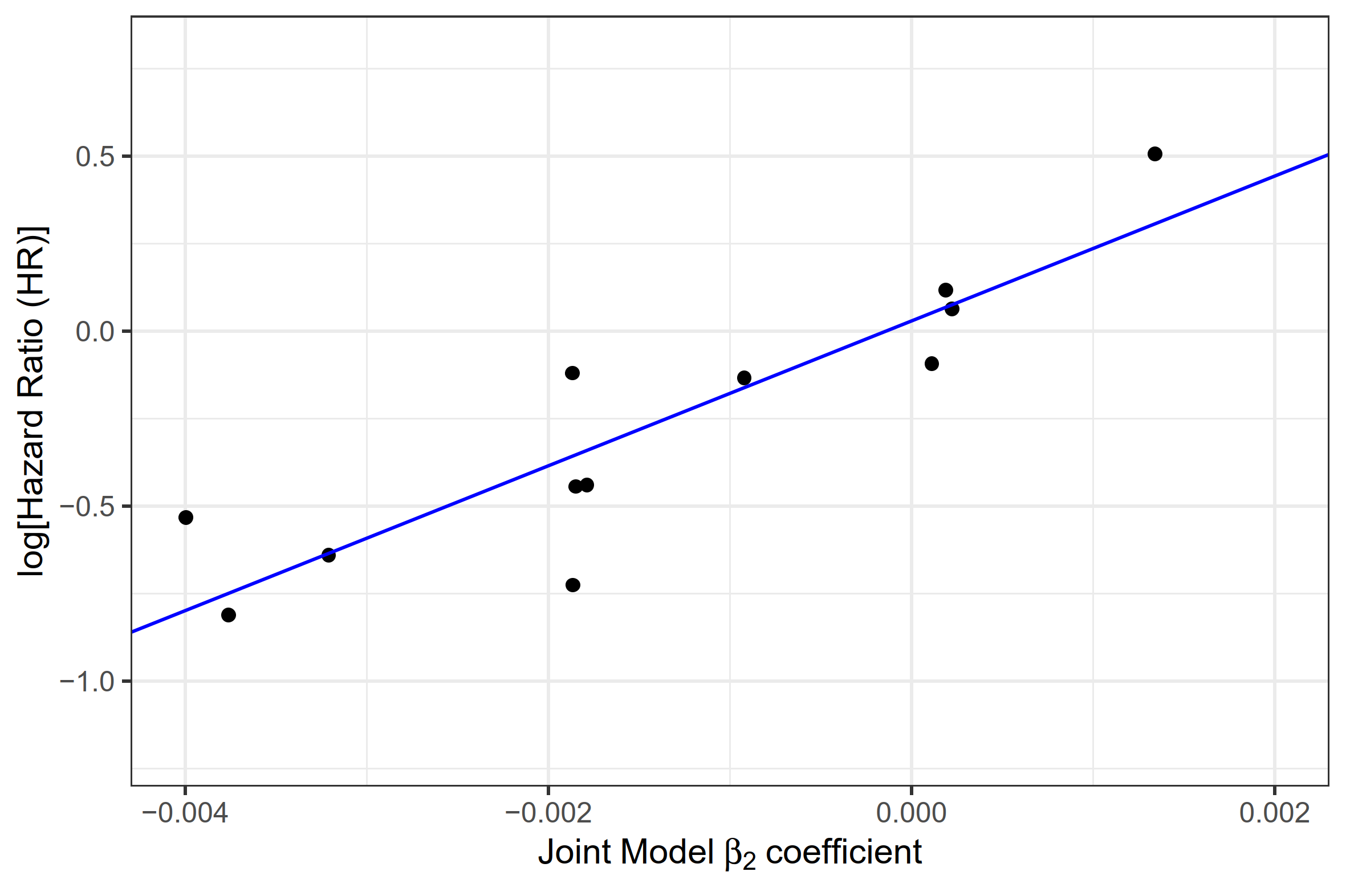

Supplementary Figure 34: OS hazard ratios at 48 months versus the ΔROPRO coefficients at 6 months for the higher caliper analysis. The included regression line considers that $\boldsymbol{\gamma=0}$.

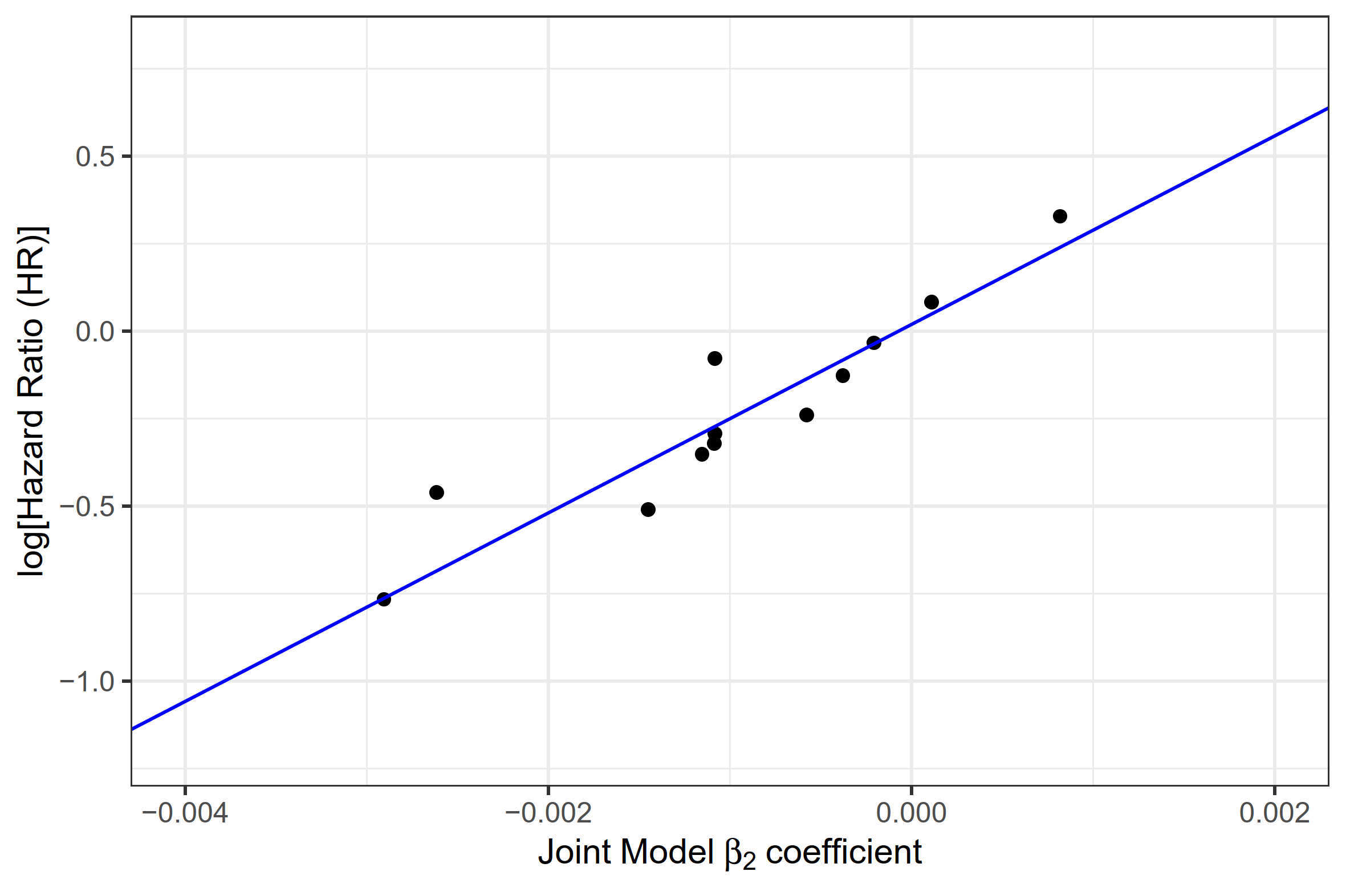
