## Supplementary Information for "Longitudinal assessment of ROPRO as an early indicator of overall survival in oncology clinical trials: a retrospective analysis"

#### ΔRisk visualization

We proposed to represent the average ΔRisk curves for each treatment arm using the LOESS smoother [1]. Since patients will drop-out at different time-points (due to death or censoring), the number of patients that are included in the ΔRisk average curves will decrease over time. As a consequence, the right censoring might induce an artificial bias in the ΔRisk curves. Particularly, when there is uneven right censoring between the two groups in analysis, (e.g., one group has a higher censoring/mortality rate than the other) then the ΔRisk curve of the group with the highest censoring/mortality might be artificially underestimated (have a negative bias). This bias could be caused by the greater number of high-risk patients that are dropped in the higher censoring/mortality group and are no longer included in the ΔRisk curve estimation. Therefore, direct interpretation of the ΔRisk curves should be done with care to avoid misinterpretation of the real effect.

The proposed ΔRisk visualization, that displays the average ΔRisk curves for each treatment arm, does not fully represent the variability present in the patient’s ΔRisk curves. Still, considering the example in Supplementary Figure 33, the smoothed ΔRisk curves captured the main difference between the treatment arms. Specifically, the ΔRisk curves of the patients that took Erlotinib or Gefinitib (green curves) are mainly clustered at high ΔRisk values, while the patients that took Osimertinib (purple curves) are clustered at lower ΔRisk values. The smoothed (average) ΔRisk curves show the same effect. The Erlotinib-Gefinitib smoothed ΔRisk curve has higher values than the Osimertinib curve over time.

#### ΔRisk modeling and statistical test

Based on our previous benchmarking study of prognostic scores in oncology [2], we selected two distinct prognostic scores to use in the ΔRisk framework. ROPRO [3], is a model based on the Cox model [4], and DeepROPRO [2] a deep learning based prognostic score based on DeepSurv [5]. These models, although of different algorithmic complexity, use the same set of 27 covariates and were developed exclusively using baseline data (before the start of treatment).

The ΔRisk value at time $t$ for patient $i$ is given by the difference between the risk at time $t$ and the baseline risk ($\Delta Risk_{i}(t)=risk_{i}(t)-risk_{i}(t=0)$). The risk values at all time-points (time $t$ and 0) were calculated by the prognostic scores (in this analysis, ROPRO and DeepROPRO, that yielded, the ΔROPRO and ΔDeepROPRO, respectively).

We used the Joint Models for Longitudinal and Time-to-event data (JM) [6] as the base of our ΔRisk statistical test. These models can jointly model the ΔRisk and the censoring process, avoiding bias due to uneven censoring of the arms. To model the ΔRisk and survival jointly we fit JMs defined as: $h_{a}(t)=h_{0}(t)exp\{\gamma\cdot\boldsymbol{treatment}+\alpha\cdot\Delta Risk(t)\}, \Delta Risk(t)=\beta_{0}+{(b}_{1}+\beta_{1})\cdot t+\beta_{2}t\cdot treatment$. Where $h_{a}(t)$ is the hazard function, $\gamma$ is the direct effect of the treatment on the hazard, $\beta_{0}$ and $\beta_{1}$ are the intercept and slope coefficients, $\beta_{2}$ is an additional slope coefficient that depends on the arm of treatment and $b_{1}$ is a slope random effect. For this specific modeling, the drug/treatment arm has an effect directly on the hazard function ($\gamma$ coefficient) and on the ΔRisk modeling ($\beta_{2}$ coefficient). We chose to not include a random intercept term ($b_{0}$) because there should be no difference in intercept between the patients since all have intercept 0 ($\Delta Risk(t=0)=0$). Nonetheless, we kept $\beta_{0}$ since it made the regression more stable. To reach this model structure we performed a sequential analysis of the different coefficients (Investigate ΔRisk and treatment effects in the Joint Model).

#### There were convergence issues in the JM regression (usually non-positive definite Hessian matrix) in the OAK and IMvigor211 clinical trials. For these two clinical trials, we changed the random effects structure, removing the $b_{1}$ slope random effect but adding a random intercept $b_{0}$.

At each time-point from 2 to 24 months we modeled the ΔRisk using JM, and ran a log-rank test, so that we could determine at which time-points there was a significant difference between the treatment arms. At each time-point we considered only biomarker measurements and death-events that occurred before the time-point currently being modeled. This procedure tries to simulate the incomplete nature of the data during clinical trial interim analysis.

#### The statistical test to detect whether there is a difference between the arms of treatment is based on the likelihood ratio. It requires that two models be fit: (i) the model that includes the treatment coefficients (defined above), and (ii) a model without the treatment terms: $h_{a}(t)=h_{0}(t)exp\{\alpha\cdot\Delta Risk(t)\}, \Delta Risk(t)=\beta_{0}+{(b}_{1}+\beta_{1})\cdot t$. In the likelihood ratio test the null hypothesis is $H_{0}:\gamma=\beta_{2}=0$ (the models (i) and (ii) are equal). Therefore, if we reject the null hypothesis there should be a difference between the treatment arms. In the cases where the random effects were changed ($b_{1}$ was replaced by $b_{0}$), both models, with and without the arm effects were changed, so that the only difference between the models was the $\gamma$ and $\beta_{2}$ terms.

##### Validate the statistical test

To evaluate the validity of the JM statistical test, in particular for small sample size, we performed simulations under the null hypothesis where there was no difference between the treatment arms. To obtain treatment arms without difference between them, we sampled patients at random from all treatment arms and randomly assigned a novel treatment arm label. We performed this random sampling analysis separately for the RWD and clinical trial data. The analysis can be summarized by the following algorithm:

#### 1. Repeat these steps (in our case 100 or 1000 times):

#### a. Sample patients from the whole dataset at random and assign the patients to one of two arms of treatment;

#### b. Apply the JM statistical test to the sampled arms and obtain the p-value;

#### c. Following the fixed significance level (in this case $\alpha=0.05$) map the p-values from the statistical test to “success” (if $p-value<\alpha)$ or “no success” (if $p-value\geq\alpha)$;

#### d. Save the “success” or “no success” value as the result of the run;

#### 2. Use the binomial test on the vector of “success” and “no success” to obtain the empirical alpha-level as (number of success)/(number of repetitions).

#### 3. By construction, the empirical alpha-level should be close to 0.05, and any deviation from 0.05 would indicate conceptual flaws in the framework, inaccuracies induced by convergence issues or inadequate assumptions on the asymptotic behavior of the test statistic of the JM test. To formalize this, the binomial test was used to check if the empirical alpha-level was different from the nominal alpha-level of 0.05 (the null hypothesis and alternative hypotheses are $H_{0}:p_{success}=0.05;H_{1}:p_{success}\neq0.05$).

#### We applied the algorithm to the RWD (Supplementary Table 56) and clinical data (Supplementary Table 57), varying the number of patients and the month that was tested. For all experiments, empirical alpha levels were around 0.05 and in none of the cases the binomial test was significant. We conclude that the proposed statistical test keeps the alpha-level throughout and can safely be applied also to smaller data sets.

#### Real-world data and clinical trial analyses

In the RWD analysis, we identified a total of 13 clinical trials that were conformant with the inclusion and exclusion criteria (Supplementary Figure 1). The list includes the LUX-Lung 6 clinical trial that would have been removed based on the exclusion criteria. Instead, we combined it with Lux-Lung 3, following the original analysis by Yang *et al.* [7], thus reducing the number of emulated clinical trials to 12. Additionally, the AURA3 clinical trial [8] did not show a difference in OS between the treatment arms. The study team hypothesized that the high patient crossover rate could have led to the statistical significance. Conversely, a previous study [9] using previous clinical trial data had shown that the Osimertinib treatment arm had a significant improvement in OS. In this analysis, we followed the results in the Mann et al. study [9] since they do not suffer from the crossover issues.

In the RWD analysis, we grouped the patient covariate measurements in monthly bins to make the sampling rate homogeneous across patients. We chose one month as the sampling frequency, instead of the usual three weeks of a chemotherapy cycle, because it was common for covariate measurements to have a higher spacing than 21 days. Conversely, in the clinical trial datasets, the covariates were measured every three weeks (21 days). Therefore, we maintained their original measurement dates. Afterwards, we dealt with missing values in two steps: (1) median imputation in the baseline variables, and (2) simple linear interpolation to impute the time-varying covariates.

##### Propensity scores

We used both RWD and clinical trial data in our analysis. Although both types of data are clinical in nature, they have fundamental differences. In clinical trials, the study team takes several measures (eligibility criteria, randomization, …) to reduce the impact of confounders between the study arms. Conversely, in RWD, we constructed the study arms from the original pool of patients. Therefore, in this case, there was no prior control for confounders. To overcome this issue, we used propensity score matching [10] on the baseline ROPRO value to minimize major risk differences between the populations. Specifically, this step matches (on a one to one basis) patients between the treatment and control arms that have similar baseline ROPRO values. Ultimately, the propensity score matching will ensure that at baseline none of the treatment arms has patients considerably more sick (higher risk) than the control arms.

To emulate the clinical trials we followed these steps: first, for each clinical trial, we selected a cohort of patients from FH that took the same medications as in the original clinical trial. Then, we applied propensity score matching on the baseline (before treatment) ROPRO value to generate two treatment arms (treatment and control) with lower confounding. For each identified clinical trial we produced two RWD emulated clinical trial datasets, differing on the maximum difference between baseline ROPRO values allowed (the caliper parameter of the propensity score). The dataset produced with the higher caliper value allowed for a larger difference between matched patients, hence more patients were included. The second dataset used a narrower caliper, and thus included less patients.

#### Correlation between early ΔRisk coefficients and final OS hazard ratios

To calculate the correlation between early ΔRisk coefficients and the final OS hazard ratios (HR) we followed the methodology in Sargent et al. [11]. They used two different methods to determine the correlation between disease-free survival (DFS) and OS. The first method is based on the coefficient of determination ($R^{2}$) of a linear regression between the early DFS and late OS HRs. The second method uses the copula $R^{2}$[12] that contrasts the patient-level DFS and OS times. In this work, we intended to use these methods by replacing DFS with the ΔRisk coefficients. However, only the first method could be applied to measure the correlation between ΔRisk coefficients and the final OS HR since the ΔRisk coefficients do not change at a patient level.

In the linear regression we expressed the logarithm of the final OS HR as a linear combination of the ΔRisk coefficients: $log(OS\_HR)=\theta_{0}+\theta_{1}\beta_{2}+\theta_{2}\gamma+\epsilon$. In the expression the $\theta$ coefficients correspond to the regression coefficients, $\beta_{2}$ to the ΔRisk trend, $\gamma$ to the treatment effect on the mortality risk (from the JM), and $\epsilon$ to the error. After the regression, we calculated the coefficient of determination ($R^{2}$). We selected the early ΔROPRO/ΔDeepROPRO trends ($\beta_{2}$) and treatment effect ($\gamma$) at 3, 6, 9 and 12 months, while the final OS HRs were calculated at 24, 36 and 48 months. The Supplementary Tables 56-58 show a high correlation between the early ΔROPRO/ΔDeepROPRO coefficients and final OS HRs for all time-points. In particular, the $R^{2}$ values were always above 0.85 and 0.75 in the higher and lower caliper analyses, respectively. Considering the analysis with the highest difference in time between the ΔROPRO/ΔDeepROPRO and OS cut-off points (3, and 48 months, respectively), the ΔROPRO (ROPRO $R^{2}$ high caliper: 0.89, higher caliper and validation trials: 0.75, lower caliper: 0.76) and ΔDeepROPRO at 3 months (DeepROPRO $R^{2}$ high caliper: 0.85, higher caliper and validation trials: 0.77, lower caliper: 0.75) had a high correlation with OS at 48 months (almost four years later). Note that the lower values in the higher caliper and validation trials had consistently lower values at 3 months, likely due to the cross-over in IMvigor211.

Additionally, we performed a second correlation analysis without the treatment effect on the mortality risk ($\gamma$). In this second analysis, we calculated the correlation the ΔROPRO/ΔDeepROPRO trends with OS, resulting in a modified the regression expression: $log(OS\_HR)=\theta_{0}+\theta_{1}\beta_{2}+\epsilon$. The results show a high correlation between the early trends and OS even without the $\gamma$ coefficient (refer to Supplementary Tables 61-63 for the full correlation results). There was a slight decrease in correlation especially at higher cut-off points of the ΔRisk coefficients (especially 12 months). We hypothesize that this effect is due to a higher prediction power of the treatment effect $\gamma$ at higher cut-off points, since the treatment effect is equivalent to the early OS.

#### Investigate ΔRisk and treatment effects in the Joint Model

We conducted three analyses: first, we investigated whether introducing a correlation effect between the ΔRisk and OS (the $\alpha$ parameter) contributed to the estimation of OS. In this case, the hypothesis tested was $H_{0}:\alpha=0, H_{1}:\alpha\neq0$. Second, we analyzed whether a treatment-dependent ΔROPRO/ΔDeepROPRO slope (the $\beta_{2}$ coefficient) informed the estimation of OS, i.e. $H_{0}:\beta_{2}=0, H_{1}:\beta_{2}\neq0$, with $\alpha\neq0$ in both hypotheses. Finally, we investigated whether the early OS treatment effect (the $\gamma$ parameter) carried extra prognostic information. The $\gamma$ parameter models the difference in OS between the treatment arms until the censoring month imposed by us in each model fit. In this third experiment, we evaluated $H_{0}:\gamma=0, H_{1}:\gamma\neq0$, with $\alpha\neq0, \beta_{2}\neq0$. We evaluated these hypotheses with the Wald (first hypothesis), and likelihood ratio tests (second and third hypotheses).

##### ΔRisk prognostic information ($\alpha$ parameter)

The mortality risk values used in the ΔRisk were derived from the ROPRO and DeepROPRO prognostic scores. These prognostic scores are correlated with the date-of-death such that a higher prognostic score value indicates a lower time-to-event. Therefore, since the ΔRisk is calculated with these prognostic score values, it should be correlated with OS. We tested this association with the statistical test described above.

Our analysis (Supplementary Table 64; additional tables available in the Github repository) demonstrated that the ΔRisk contributed to the estimation of OS. The null hypothesis was rejected for all clinical trials tested, typically with overwhelming evidence. For the majority of clinical trials, the null hypothesis was rejected for all time-points. Therefore, we conclude that the correlation between ΔRisk and OS contributed to the estimation of OS.

##### Treatment-dependent slope contribution for OS estimation ($\beta_{2}$)

The $\beta_{2}$ coefficient introduces an additional slope term, allowing each treatment arm to have a different ΔRisk slope. For this analysis, we evaluated the concordance between the ΔRisk slope ($\beta_{1}+\beta_{2}$) and OS treatment benefit. Additionally, we analyzed whether the statistical test described above (only with $\beta_{2}$, without $\gamma$) could detect the treatment benefit before the log-rank test.

The ΔRisk slope was concordant with the final OS benefit for 11 (ROPRO) and 12 (DeepROPRO) out of 15 clinical trials (Supplementary Table 65; additional tables available in the Github repository). Additionally, the ΔRisk framework identified the difference between the arms 1.75, 1.86, and over 6 months before the log-rank test for the higher caliper, lower caliper, and validation with clinical trials, respectively.

##### Treatment effect contribution to OS estimation ($\gamma$)

The previous analysis demonstrated that the ΔRisk slope identified the treatment arm with OS benefit. Next we investigated whether the treatment effect in the hazard function ($\gamma$) further improved the OS estimation.

Our results (Supplementary Table 66; additional tables available in the Github repository) demonstrate that the treatment effect did not improve OS estimation in the clinical trials for which there was no OS benefit (PRONOUNCE, PointBreak, NCT00540514, and NCT00520676). For the remaining trials (where one medication had OS benefit) the treatment effect contributed to OS estimation in almost all clinical trials but FLAURA (higher caliper), LUX-Lung 3+6 (lower caliper), AURA3 (lower caliper), and OAK. The treatment effect was especially important for later time-points (after 5/6 months). Given this result, we decided to include the $\gamma$ coefficient into the combined ΔRisk/early OS statistical test we described in the manuscript, since it further improved the performance. We note, however, that in many cases the ROPRO time course alone would have enabled the right decision.

#### Implementation

All the analyses were performed using R 4.0.4 [13] and Python 3.6. The ROPRO was calculated as specified in the original publication [3]. The DeepROPRO model had been previously fit in our previous analysis [2]. The propensity score matching, DeepSurv, JM were implemented in the MatchIt [14], DeepSurv [5] and JM [6] packages, respectively. The analysis code and additional supplementary tables/figures are available in <https://github.com/loureirh/deltaRisk>.

### Supplementary Tables

**Supplementary Table 1: Clinical trials selected for real-world data analysis.**

| **Medication Type** | **Clinical Trial Acronym** | **Treatment arm 1** | **Treatment arm 2** | **Treatment arm with highest overall survival** |
| --- | --- | --- | --- | --- |
| Immunotherapy | KEYNOTE-189 [15] | Carboplatin/Cisplatin + Pembrolizumab + Pemetrexed | Carboplatin/Cisplatin + Pemetrexed | Treatment arm 1 |
|  | KEYNOTE-024 [16] | Pembrolizumab | Platinum-doublet (carboplatin + pemetrexed, cisplatin + pemetrexed, carboplatin + gemcitabine, cisplatin + gemcitabine, or carboplatin + paclitaxel) | Treatment arm 1 |
|  | KEYNOTE-042 [17] | Pembrolizumab | Platinum-doublet (carboplatin + pemetrexed, carboplatin + paclitaxel) | Treatment arm 1 |
|  | KEYNOTE-407 [18] | Carboplatin + Pembrolizumab + Paclitaxel/nab-paclitaxel | Carboplatin + Paclitaxel/nab-paclitaxel | Treatment arm 1 |
| Anti-VEGF monoclonal antibodies | PRONOUNCE [19] | Carboplatin + Pemetrexed | Bevacizumab + Carboplatin + Paclitaxel | - |
|  | PointBreak [20] | Bevacizumab + Carboplatin + Pemetrexed | Bevacizumab + Carboplatin + Paclitaxel | - |
| TK-inhibitors | PROFILE 1014 [21] | Crizotinib | Carboplatin/Cisplatin + Pemetrexed | Treatment arm 1 |
|  | FLAURA [22] | Osimertinib | Gefitinib / Erlotinib | Treatment arm 1 |
|  | LUX-Lung 3 and 6 [7] | Afatinib | Cisplatin + Pemetrexed, or Cisplatin + Gemcitabine | Treatment arm 1 |
|  | AURA3 [8], [9] | Osimertinib | Carboplatin/Cisplatin + Pemetrexed | Treatment arm 1^[1]^ |
| Platinum-doublet chemotherapy | NCT00540514 [23] | Carboplatin + Nab-Paclitaxel | Carboplatin + Paclitaxel | - |
|  | NCT00520676 [24] | Carboplatin + Pemetrexed | Carboplatin + Docetaxel | - |

**Supplementary table 56: Association between the early ΔROPRO and ΔDeepROPRO** $\beta_{\boldsymbol{2}}$ **and** $\gamma$ **coefficients and the final Overall Survival (OS) Hazard Ratio (HR) at several readout dates for the higher caliper dataset. The confidence intervals were calculated with bootstrap (1000 resamplings).**

| **JM model [months]** | **Emulated dataset OS readout [months]** | **ΔROPRO** $\boldsymbol{R}^{\boldsymbol{2}}$ **(Confidence interval)** | **ΔDeepROPRO** $\boldsymbol{R}^{\boldsymbol{2}}$ **(Confidence interval)** |
| --- | --- | --- | --- |
| 3 | 24 | 0.91 (0.73, 0.98) | 0.89 (0.67, 0.98) |
| 6 | 24 | 0.94 (0.8, 0.99) | 0.92 (0.72, 0.99) |
| 9 | 24 | 0.96 (0.88, 0.99) | 0.95 (0.84, 1) |
| 12 | 24 | 0.96 (0.9, 0.99) | 0.96 (0.88, 1) |
| 3 | 36 | 0.88 (0.66, 0.97) | 0.85 (0.58, 0.97) |
| 6 | 36 | 0.88 (0.66, 0.99) | 0.85 (0.58, 0.98) |
| 9 | 36 | 0.91 (0.76, 1) | 0.89 (0.73, 1) |
| 12 | 36 | 0.92 (0.81, 0.99) | 0.91 (0.77, 1) |
| 3 | 48 | 0.87 (0.66, 0.97) | 0.85 (0.58, 0.97) |
| 6 | 48 | 0.86 (0.63, 0.99) | 0.83 (0.53, 0.98) |
| 9 | 48 | 0.88 (0.69, 0.99) | 0.86 (0.65, 0.99) |
| 12 | 48 | 0.89 (0.73, 0.99) | 0.88 (0.68, 0.99) |

**Supplementary table 57: Association between the early ΔROPRO and ΔDeepROPRO** $\beta_{\boldsymbol{2}}$ **and** $\gamma$ **coefficient and the final Overall Survival (OS) Hazard Ratio (HR) at several readout dates for the higher caliper datasets and validation clinical trials (IMpower150, IMvigor211, OAK). The confidence intervals were calculated with bootstrap (1000 resamplings).**

| **JM model [months]** | **Emulated dataset OS readout [months]** | **ΔROPRO** $\boldsymbol{R}^{\boldsymbol{2}}$ **(Confidence interval)** | **ΔDeepROPRO** $\boldsymbol{R}^{\boldsymbol{2}}$ **(Confidence interval)** |
| --- | --- | --- | --- |
| 3 | 24 | 0.77 (0.42, 0.95) | 0.78 (0.54, 0.95) |
| 6 | 24 | 0.91 (0.78, 0.97) | 0.9 (0.77, 0.97) |
| 9 | 24 | 0.94 (0.87, 0.98) | 0.94 (0.86, 0.98) |
| 12 | 24 | 0.96 (0.93, 0.99) | 0.96 (0.91, 0.99) |
| 3 | 36 | 0.75 (0.4, 0.94) | 0.77 (0.52, 0.94) |
| 6 | 36 | 0.86 (0.73, 0.96) | 0.85 (0.68, 0.97) |
| 9 | 36 | 0.91 (0.82, 0.98) | 0.89 (0.78, 0.98) |
| 12 | 36 | 0.93 (0.84, 0.99) | 0.92 (0.82, 0.99) |
| 3 | 48 | 0.75 (0.38, 0.94) | 0.77 (0.51, 0.94) |
| 6 | 48 | 0.84 (0.68, 0.96) | 0.83 (0.63, 0.96) |
| 9 | 48 | 0.88 (0.73, 0.98) | 0.87 (0.74, 0.98) |
| 12 | 48 | 0.9 (0.79, 0.98) | 0.9 (0.77, 0.99) |

**Supplementary table 58: Association between the early ΔROPRO and ΔDeepROPRO** $\beta_{\boldsymbol{2}}$ **and** $\gamma$ **coefficient and the final Overall Survival (OS) Hazard Ratio (HR) at several readout dates for the lower caliper datasets. The confidence intervals were calculated with bootstrap (1000 resamplings).**

| **JM model [months]** | **Emulated dataset OS readout [months]** | **ΔROPRO** $\boldsymbol{R}^{\boldsymbol{2}}$ **(Confidence interval)** | **ΔDeepROPRO** $\boldsymbol{R}^{\boldsymbol{2}}$ **(Confidence interval)** |
| --- | --- | --- | --- |
| 3 | 24 | 0.79 (0.55, 0.97) | 0.79 (0.5, 0.97) |
| 6 | 24 | 0.88 (0.69, 0.99) | 0.91 (0.73, 0.99) |
| 9 | 24 | 0.87 (0.71, 0.99) | 0.91 (0.77, 0.99) |
| 12 | 24 | 0.91 (0.79, 0.99) | 0.94 (0.84, 0.99) |
| 3 | 36 | 0.78 (0.51, 0.98) | 0.76 (0.43, 0.98) |
| 6 | 36 | 0.84 (0.57, 0.98) | 0.88 (0.66, 0.99) |
| 9 | 36 | 0.8 (0.54, 0.98) | 0.83 (0.59, 0.99) |
| 12 | 36 | 0.83 (0.58, 0.98) | 0.86 (0.66, 0.99) |
| 3 | 48 | 0.76 (0.44, 0.98) | 0.75 (0.37, 0.99) |
| 6 | 48 | 0.82 (0.55, 0.98) | 0.86 (0.61, 0.99) |
| 9 | 48 | 0.77 (0.5, 0.99) | 0.8 (0.51, 0.99) |
| 12 | 48 | 0.79 (0.5, 0.98) | 0.83 (0.57, 0.99) |

**Supplementary table 59: Simulation results of the validation of the ΔRisk statistical test under the null hypothesis, based on FH data.**

| **Seed** | **Month tested** | **Number of Simulations** | **Number of Patients** | **Empirical α (confidence interval)** | $\boldsymbol{p}$ **binomial test** |
| --- | --- | --- | --- | --- | --- |
| 1000 | 6 | 1000 | 100 | 0.058 (0.044, 0.074) | 0.245 |
| 1000 | 6 | 1000 | 1000 | 0.04 (0.028, 0.054 | 0.167 |
| 1000 | 12 | 1000 | 100 | 0.059 (0.045, 0.075) | 0.191 |
| 1000 | 12 | 1000 | 1000 | 0.05 (0.039, 0.068) | 0.771 |

**Supplementary table 60: Simulation results of the validation of the ΔRisk statistical test on the clinical trial data.**

| **Seed** | **Month tested** | **Number of Simulations** | **Number of Patients** | **Empirical α (confidence interval)** | $\boldsymbol{p}$ **binomial test** |
| --- | --- | --- | --- | --- | --- |
| 456 | 12 | 100 | 1000 | 0.06 (0.022, 0.126) | 0.642 |
| 456 | 12 | 100 | 100 | 0.08 (0.035, 0.152) | 0.165 |
| 456 | 12 | 1000 | 100 | 0.049 (0.036, 0.064) | 0.942 |

**Supplementary table 61: Association between the early ΔROPRO and ΔDeepROPRO** $\beta_{\boldsymbol{2}}$ **and the Overall Survival (OS) Hazard Ratio (HR) at several readout dates for the higher caliper dataset. The confidence intervals were calculated with bootstrap (1000 resamplings).**

| **JM model [months]** | **Emulated dataset OS readout [months]** | **ΔROPRO** $\boldsymbol{R}^{\boldsymbol{2}}$ **(Confidence interval)** | **ΔDeepROPRO** $\boldsymbol{R}^{\boldsymbol{2}}$ **(Confidence interval)** |
| --- | --- | --- | --- |
| 3 | 24 | 0.91 (0.68, 0.98) | 0.89 (0.56, 0.98) |
| 6 | 24 | 0.94 (0.74, 0.99) | 0.91 (0.66, 0.99) |
| 9 | 24 | 0.93 (0.73, 0.99) | 0.91 (0.6, 0.99) |
| 12 | 24 | 0.92 (0.72, 0.99) | 0.89 (0.61, 0.99) |
| 3 | 36 | 0.85 (0.59, 0.96) | 0.82 (0.49, 0.96) |
| 6 | 36 | 0.88 (0.64, 0.98) | 0.84 (0.51, 0.98) |
| 9 | 36 | 0.88 (0.62, 0.98) | 0.85 (0.45, 0.98) |
| 12 | 36 | 0.87 (0.54, 0.98) | 0.83 (0.45, 0.97) |
| 3 | 48 | 0.84 (0.56, 0.96) | 0.81 (0.43, 0.96) |
| 6 | 48 | 0.86 (0.6, 0.98) | 0.83 (0.53, 0.97) |
| 9 | 48 | 0.87 (0.58, 0.99) | 0.84 (0.48, 0.98) |
| 12 | 48 | 0.86 (0.56, 0.99) | 0.83 (0.46, 0.98) |

**Supplementary table 62: Association between the early ΔROPRO and ΔDeepROPRO** $\beta_{\boldsymbol{2}}$ **and the Overall Survival (OS) Hazard Ratio (HR) at several readout dates for the higher caliper datasets and validation clinical trials (IMpower150, IMvigor211, OAK). The confidence intervals were calculated with bootstrap (1000 resamplings).**

| **JM model [months]** | **Emulated dataset OS readout [months]** | **ΔROPRO** $\boldsymbol{R}^{\boldsymbol{2}}$ **(Confidence interval)** | **ΔDeepROPRO** $\boldsymbol{R}^{\boldsymbol{2}}$ **(Confidence interval)** |
| --- | --- | --- | --- |
| 3 | 24 | 0.77 (0.36, 0.95) | 0.78 (0.46, 0.95) |
| 6 | 24 | 0.88 (0.72, 0.96) | 0.88 (0.69, 0.97) |
| 9 | 24 | 0.89 (0.7, 0.97) | 0.88 (0.68, 0.97) |
| 12 | 24 | 0.88 (0.68, 0.97) | 0.87 (0.62, 0.98) |
| 3 | 36 | 0.74 (0.39, 0.94) | 0.75 (0.41, 0.93) |
| 6 | 36 | 0.85 (0.64, 0.95) | 0.84 (0.6, 0.95) |
| 9 | 36 | 0.86 (0.69, 0.96) | 0.85 (0.57, 0.95) |
| 12 | 36 | 0.86 (0.62, 0.96) | 0.84 (0.56, 0.96) |
| 3 | 48 | 0.72 (0.36, 0.93) | 0.73 (0.39, 0.91) |
| 6 | 48 | 0.84 (0.64, 0.95) | 0.83 (0.63, 0.94) |
| 9 | 48 | 0.85 (0.68, 0.96) | 0.84 (0.58, 0.96) |
| 12 | 48 | 0.85 (0.61, 0.96) | 0.83 (0.56, 0.96) |

**Supplementary table 63: Association between the early ΔROPRO and ΔDeepROPRO** $\beta_{\boldsymbol{2}}$ **and the Overall Survival (OS) Hazard Ratio (HR) at several readout dates for the lower caliper datasets. The confidence intervals were calculated with bootstrap (1000 resamplings).**

| **JM model [months]** | **Emulated dataset OS readout [months]** | **ΔROPRO** $\boldsymbol{R}^{\boldsymbol{2}}$ **(Confidence interval)** | **ΔDeepROPRO** $\boldsymbol{R}^{\boldsymbol{2}}$ **(Confidence interval)** |
| --- | --- | --- | --- |
| 3 | 24 | 0.79 (0.51, 0.94) | 0.79 (0.46, 0.97) |
| 6 | 24 | 0.78 (0.44, 0.95) | 0.75 (0.39, 0.94) |
| 9 | 24 | 0.79 (0.49, 0.95) | 0.77 (0.44, 0.94) |
| 12 | 24 | 0.79 (0.49, 0.95) | 0.78 (0.47, 0.94) |
| 3 | 36 | 0.78 (0.45, 0.98) | 0.76 (0.37, 0.98) |
| 6 | 36 | 0.73 (0.35, 0.96) | 0.7 (0.28, 0.95) |
| 9 | 36 | 0.75 (0.37, 0.95) | 0.72 (0.32, 0.94) |
| 12 | 36 | 0.75 (0.37, 0.96) | 0.73 (0.33, 0.94) |
| 3 | 48 | 0.76 (0.42, 0.98) | 0.74 (0.34, 0.99) |
| 6 | 48 | 0.72 (0.26, 0.97) | 0.69 (0.28, 0.97) |
| 9 | 48 | 0.73 (0.34, 0.96) | 0.72 (0.28, 0.96) |
| 12 | 48 | 0.73 (0.33, 0.96) | 0.72 (0.29, 0.95) |

**Supplementary table 64: Contribution of the ΔRisk to the OS estimation. This analysis did not include a drug effects (**$\gamma$ **and** $\beta_{\boldsymbol{2}}$**).**

|  | **Trial Name** | **ΔROPRO significant contribution to the OS estimation [months]** | **ΔDeepROPRO significant contribution to the OS estimation [months]** |
| --- | --- | --- | --- |
| Higher caliper | KEYNOTE-189 | 2-24 | 2-24 |
|  | KEYNOTE-024 | 2-24 | 2-24 |
|  | KEYNOTE-042 | 2-24 | 2-24 |
|  | KEYNOTE-407 | 2-24 | 2-24 |
|  | PRONOUNCE | 2-24 | 2-24 |
|  | PointBreak | 2-24 | 2-24 |
|  | PROFILE 1014 | 2-24 | 2-24 |
|  | FLAURA | 2-24 | 2-24 |
|  | LUX-Lung 3+6 | 2-24 | 3-24 |
|  | NCT00540514 | 2-24 | 2-24 |
|  | AURA3 | 2-24 | 2-24 |
|  | NCT00520676 | 2-24 | 2-24 |
| Lower caliper | KEYNOTE-189 | 2-24 | 2-24 |
|  | KEYNOTE-024 | 2-24 | 2-24 |
|  | KEYNOTE-042 | 2-24 | 2-24 |
|  | KEYNOTE-407 | 4-24 | 4-24 |
|  | PRONOUNCE | 2-24 | 2-24 |
|  | PointBreak | 2,4-24 | 2-24 |
|  | PROFILE 1014 | 2,4,6-24 | 2-24 |
|  | FLAURA | 2-13,15-23 | 2-13,15,17-19 |
|  | LUX-Lung 3+6 | 3-24 | 3-24 |
|  | NCT00540514 | 2-24 | 2-24 |
|  | AURA3 | 4-24 | 4-24 |
|  | NCT00520676 | 3-24 | 2-24 |
| Validation with Clinical Trials | IMpower150 | 3-14,16-18,21-24 | 2-20,21-24 |
|  | IMvigor211 | 2-24 | 2-24 |
|  | OAK | 2-24 | 2-24 |

**Supplementary table 65: ΔRisk analysis of the** $\gamma$ **parameter. Treatment arms with overall survival (OS) and ΔRisk benefit for the higher caliper, lower caliper and validation with clinical trial analyses without treatment arm effect on the hazard function.**

|  | **Trial Name** | **OS treatment benefit** | **OS difference significant [months]^[2]^** | **ΔRisk treatment benefit** | **ΔRisk difference significant [months]** | **ΔRisk detects OS benefit? [months]^[3]^** |
| --- | --- | --- | --- | --- | --- | --- |
| Higher caliper | KEYNOTE-189 | Platinum + Pembrolizumab + Pemetrexed | 2-24 | Platinum + Pembrolizumab + Pemetrexed | 2-24 | Yes [0 months] |
|  | KEYNOTE-024 | Pembrolizumab | 5-24 | Pembrolizumab | 2-24 | Yes [3 months] |
|  | KEYNOTE-042 | Pembrolizumab | 4-24 | Pembrolizumab | 2-24 | Yes [2 months] |
|  | KEYNOTE-407 | Carboplatin + Pembrolizumab + Paclitaxel / nab-Paclitaxel | 4-24 | Carboplatin + Pembrolizumab + Paclitaxel / nab-Paclitaxel | 4-24 | Yes [0 months] |
|  | PRONOUNCE | - | 2-5^[4]^ | Bevacizumab + Carboplatin + Paclitaxel | 2-24 | No |
|  | PointBreak | - | - | Bevacizumab + Carboplatin + Paclitaxel | 9-24 (ROPRO), 7-24 (DeepROPRO) | No |
|  | PROFILE 1014 | Crizotinib | 5-24 | Crizotinib | 2-24 | Yes [3 months] |
|  | FLAURA | Osimertinib | 7-24 | Osimertinib | 2-24 | Yes [7 months] |
|  | LUX-Lung 3+6 | Afatinib | 3-24 | Afatinib | 2-24 | Yes [1 month] |
|  | NCT00540514 | - | - | - | - | Yes |
|  | AURA3 | Osimertinib | 2-24 | Osimertinib | 2-24 | Yes [0 months] |
|  | NCT00520676 | - | - | - | - | Yes |
| Lower caliper | KEYNOTE-189 | Platinum + Pembrolizumab + Pemetrexed | 4-24 | Platinum + Pembrolizumab + Pemetrexed | 2-24 | Yes [2 months] |
|  | KEYNOTE-024 | Pembrolizumab | 5-24 | Pembrolizumab | 2-24 | Yes [3 months] |
|  | KEYNOTE-042 | Pembrolizumab | 5-24 | Pembrolizumab | 2-24 | Yes [3 months] |
|  | KEYNOTE-407 | -^[5]^ | 5-7 | Carboplatin + Pembrolizumab + Paclitaxel / nab-paclitaxel | 3-24 | -^[6]^ |
|  | PRONOUNCE | - | 2-3^3^ | Bevacizumab + Carboplatin + Paclitaxel | 2-24 | No |
|  | PointBreak | - | - | - | - | Yes |
|  | PROFILE 1014 | Crizotinib | 5-24 | Crizotinib | 4-24 | Yes [1 month] |
|  | FLAURA | Osimertinib | 3,5-9,11-13,15-24 | Osimertinib | 2-24 | Yes [1 month] |
|  | LUX-Lung 3+6 | Afatinib | 5-24 | Afatinib | 3-24 | Yes [2 months] |
|  | NCT00540514 | - | - | - | - | Yes |
|  | AURA3 | Osimertinib | 3-24 | Osimertinib | 2-24 | Yes [1 month] |
|  | NCT00520676 | - | - | - | - | Yes |
| Validation with Clinical Trials | IMpower150 | ABCP | 10-24 | ABCP | 3-16 (ROPRO), 3-18 (DeepROPRO) | No |
|  | IMvigor211^[7]^ | Atezolizumab | 3,15-24^[8]^ | Atezolizumab | 2-5,11-16 (ROPRO),2-5,8-24 (DeepROPRO) | No (ROPRO)  Yes [7 months] DeepROPRO |
|  | OAK | Atezolizumab | 9-24 | Atezolizumab | 2-24 | Yes [7 months] |

**Supplementary table 66: Contribution of the** $\gamma$ **parameter to the OS estimation.**

|  | **Trial Name** | **OS treatment benefit** | $\gamma$ **significant contribution to the OS estimation in ΔROPRO model [months]** | $\gamma$ **significant contribution to the OS estimation in ΔDeepROPRO model [months]** |
| --- | --- | --- | --- | --- |
| Higher caliper | KEYNOTE-189 | Platinum + Pembrolizumab + Pemetrexed | 5-24 | 5-24 |
|  | KEYNOTE-024 | Pembrolizumab | 2-3,7-24 | 2-3,7-24 |
|  | KEYNOTE-042 | Pembrolizumab | 2-3,7-24 | 2-4,7-24 |
|  | KEYNOTE-407 | Carboplatin + Pembrolizumab + Paclitaxel / nab-Paclitaxel | 5-24 | 6-24 |
|  | PRONOUNCE | - | - | - |
|  | PointBreak | - | - | - |
|  | PROFILE 1014 | Crizotinib | 7-24 | 7-24 |
|  | FLAURA | Osimertinib | - | - |
|  | LUX-Lung 3+6 | Afatinib | 3-24 | 3-24 |
|  | NCT00540514 | - | - | - |
|  | AURA3 | Osimertinib | 5-24 | 5-24 |
|  | NCT00520676 | - | - | - |
| Lower caliper | KEYNOTE-189 | Platinum + Pembrolizumab + Pemetrexed | 7-24 | 7-24 |
|  | KEYNOTE-024 | Pembrolizumab | 2,8-24 | 2-4,8-24 |
|  | KEYNOTE-042 | Pembrolizumab | 2-3,8-24 | 2-4,8-24 |
|  | KEYNOTE-407 | -^[9]^ | - | - |
|  | PRONOUNCE | - | - | - |
|  | PointBreak | - | - | - |
|  | PROFILE 1014 | Crizotinib | 5-24 | 5,7-24 |
|  | FLAURA | Osimertinib | 2,20-24 | 20-21 |
|  | LUX-Lung 3+6 | Afatinib | 9-11 | - |
|  | NCT00540514 | - | - | - |
|  | AURA3 | Osimertinib | - | - |
|  | NCT00520676 | - | - | - |
| Validation with Clinical Trials | IMpower150 | ABCP | 10-24 | 10-24 |
|  | IMvigor211 | Atezolizumab | 14-24 | 15-24 |
|  | OAK | Atezolizumab | - | - |

1. Based on the results from Mann et al. [9].
2. The time-points (in months) for which the log-rank test identified a difference between the treatment arms. We performed the log-rank test (and ΔRisk statistical test) for all months between 2 and 24.
3. Whether the ΔRisk detects the same treatment benefit as OS. In square brackets, the how many months earlier the ΔRisk detects the treatment benefit (only available if a difference between the treatment arms was identified in both tests).
4. The OS benefit for the Bevacizumab + Carboplatin + Paclitaxel treatment arm was only identified between two to five months in the higher caliper analysis, and two to three months on the lower caliper analysis.
5. The Pembrolizumab-combo treatment arm had OS benefit only between five and seven months.
6. Since no OS difference was identified, we cannot calculate the month difference.
7. IMvigor211 was the only urothelial carcinoma clinical trial. All remaining trials focused on advanced non-small-cell lung cancer.
8. The Chemotherapy treatment arm had OS (three months) and ΔRisk (two to four months) before the crossover in the kaplan meier curves.
9. The Pembrolizumab-combo treatment arm had OS benefit only between five and seven months.
